# Physical exercise is a risk factor for amyotrophic lateral sclerosis: Convergent evidence from mendelian randomisation, transcriptomics and risk genotypes

**DOI:** 10.1101/2020.11.24.20238063

**Authors:** Thomas H Julian, Nicholas Glascow, A Dylan Fisher Barry, Tobias Moll, Calum Harvey, Yann C Klimentidis, Michelle Newell, Sai Zhang, Michael P Snyder, Johnathan Cooper-Knock, Pamela J Shaw

**Author notes:** Correspondence should be addressed to PJS, Professor Pamela J Shaw, Sheffield Institute for Translational Neuroscience., 385 Glossop Road, Sheffield S10 2HQ, United Kingdom.

## Abstract

**Background:** Amyotrophic lateral sclerosis (ALS) is a universally fatal neurodegenerative disease. ALS is determined by gene-environment interactions and improved understanding of these interactions may lead to effective personalised medicine. The role of physical exercise in the development of ALS is currently controversial.

**Methods:** We dissected the exercise-ALS relationship in a series of two-sample Mendelian randomisation (MR) experiments. We then we tested for enrichment of ALS genetic risk within exercise-associated transcriptome changes. Finally, we applied a validated physical activity (PA) questionnaire in a small cohort of genetically selected ALS patients.

**Findings:** We present MR evidence supporting a causal relationship between genetic liability to strenuous leisure-time exercise and ALS (multiplicative random effects IVW, p=0.01). Transcriptomic analysis revealed that genes with altered expression in response to acute exercise are enriched with known ALS risk genes (permutation test, p=0.013) including *C9ORF72*, and with ALS-associated rare variants of uncertain significance. Questionnaire evidence revealed that age of onset is inversely proportional to historical PA for *C9ORF72*-ALS (linear regression, t=- 2.28, p=0.036) but not for non*-C9ORF72*-ALS. Moreover, compared to non-*C9ORF72*-ALS patients and neurologically normal controls, *C9ORF72*-ALS cases reported the highest minimum average PA (20.9kJ/kg/day) consistent with an exercise threshold for penetrance.

**Interpretation:** Our MR approach suggests a positive causal relationship between ALS and physical exercise. Exercise is likely to cause motor neuron injury only in patients with a risk- genotype. Consistent with this we have shown that ALS risk genes are activated in response to exercise. In particular, we propose that G4C2-repeat expansion of *C9ORF72* predisposes to exercise-induced ALS.

**Funding:** We acknowledge support from the Wellcome Trust (JCK, 216596/Z/19/Z), NIHR (PJS, NF-SI-0617-10077; IS-BRC-1215-20017) and NIH (MPS, CEGS 5P50HG00773504, 1P50HL083800, 1R01HL101388, 1R01-HL122939, S10OD025212, and P30DK116074, UM1HG009442).

**RESEARCH IN CONTEXT:** *Evidence before this study:* The role of physical activity (PA) as a risk factor for ALS was evaluated in a systematic review of 26 studies performed by Lacorte *et al*. in 2016. The authors concluded that there was insufficient evidence to draw a firm conclusion. The authors highlighted limitations of previous studies relating to heterogeneous classification of PA and ALS. They noted that none of the published literature achieved the highest quality rating in the Newcastle Ottawa Scale, which they attribute to methodological challenges posed by the rarity and severity of the disease. Failure to address genetic subtypes of ALS was proposed as a shortcoming in the studies surveyed. To identify more recent publications, we conducted a literature search using the PubMed database for articles published between 01/01/2015 - 11/11/2020. The search terms used were (“Amyotrophic lateral sclerosis”[Title/Abstract] OR “motor neuron disease”[Title/Abstract] OR MND[Title/Abstract] OR ALS[Title/Abstract]) AND (PA[Title/Abstract] OR exercise[Title/Abstract] OR “physical activity”[Title/Abstract] OR sport[Title/Abstract]). This search strategy yielded 182 results and we filtered for original, observational, human-subject studies but we excluded case series with <10 participants and case reports. This process identified 12 further relevant publications which report opposite conclusions without significantly addressing the methodological issues highlighted above. A single recent study used linkage disequilibrium score regression and mendelian randomisation to test for a causal relationship between ALS and a number of UK biobank questionnaire items including participation in light DIY, walking for pleasure and moderate activity duration, but this study did not address the relationship between ALS and strenuous, frequent physical exercise.

*Added value of this study:* In the present study, we have exploited the methodological advantages of mendelian randomisation (MR) to counter bias, together with a tailored approach to PA exposure aimed at isolating strenuous, frequent physical exercise. We achieved this by selecting and combining UK biobank questionnaire items. In contrast to previous studies, we have addressed the gene-environment interaction by measuring the effect of exercise on expression of ALS risk genes. Furthermore, we have considered in detail the relationship between PA and the most frequent genetic risk factor for ALS: hexanucleotide (G4C2) repeat expansion of *C9ORF72*. Our data suggests that genetic liability to leisure time physical activity is a risk factor for ALS and *C9ORF72*-ALS in particular. In addition, we offer evidence that a number of known ALS-associated genetic variants are functionally linked to the physiological response to exercise.

*Implications of all the available evidence:* Our results indicate that participation in leisure time physical activity is a risk factor for ALS particularly in the context of certain risk genotypes. This could explain some of the controversy in previous studies which have largely neglected genetic heterogeneity within ALS patients. Our results form a platform for future research to explore the interaction between specific genotypes and exercise-induced ALS in a prospective manner with larger numbers, and in selected pedigrees. Ultimately this could lead to the design of personalised medicine including lifestyle advice regarding physical activity, to patients with ALS and their family members.

## INTRODUCTION

Amyotrophic lateral sclerosis (ALS) is a devastating, rapidly progressive and relatively common neurodegenerative disease affecting ∼1/400 individuals.^1^ ALS is thought to result from interplay between the environment and risk-genotypes.^2^ Like other complex diseases, the risk of ALS has a significant heritable component, but penetrance of specific genetic variants is variable which is consistent with environmental modifiers. ALS is notable for the late age of onset, even in rare monogenic forms, and this has been interpreted as a ‘multiple-hit’ process involving sequential genetic and environmental insults.^3,4,5,6^ Identification of specific gene-environment interactions may open the door to personalised medicine and even disease prevention.

To date, identification of environmental risk factors for ALS has been limited. ALS has a higher incidence and lower age of onset in professional sportspeople which led to the proposal that exercise is a risk factor for ALS.^7^ However, epidemiological studies attempting to quantify exercise-history in ALS patients have produced conflicting results.^8–11^ These studies have largely relied on questionnaire-based quantification of exercise in an unselected cohort of ALS patients. Inherent in these approaches is selection bias, recall bias, and confounding due to the effect of exercise on other causes of mortality. In particular, failure to consider genetic heterogeneity within ALS cohorts may have masked specific gene-environment interactions. Of the conflicting results that have been published, it is potentially significant that a positive relationship between ALS risk and exercise has often been reported in populations with high incidence of the G4C2 repeat expansion within *C9ORF72*.^*9,12,13*^ *C9ORF72* expansion is the most common genetic risk factor for ALS, but shows marked phenotypic variability including incomplete penetrance.^14^ This variability has led to the proposal of various genetic and environmental modifiers, but to date no specific modifier has been conclusively demonstrated.

Exercise itself is a heterogeneous activity. Muscle fibres and motor neurons are sub-specialised for aerobic and anaerobic conditions. Skeletal muscle fibres are categorised as fast twitch (type IIa, IIb and IIx) or slow-twitch (type I) according to functional (e.g. contractile speed) and metabolic properties.^15^ In ALS it is the motor neurons supplying type IIb muscle fibres responsible for anaerobic burst activity which are most vulnerable to the disease process.^16,17^ On this basis we propose that ALS is associated specifically with vigorous exercise and indeed this hypothesis is consistent with a number of previous studies. ^9,18^ In designing the present study we therefore focused on intense, anaerobic, burst activity often undertaken in leisure, training and competition physical exercise activities.^19^

Two-sample mendelian randomisation (MR) allows evaluation of the relationship between an exposure and an outcome through upstream genetic correlates in independent cohorts. This ameliorates much of the bias which has confounded previous studies of historical-exercise in ALS patients. We set out to use MR to test whether physical exercise is a risk factor for the development of ALS. We hypothesised that some of the inconsistency between previous studies may reflect heterogeneity in exercise measures and we therefore focused on genome-wide association studies (GWAS) measuring strenuous leisure-time activity. To address genetic heterogeneity we evaluated the genetic association between exercise-associated gene expression changes and ALS risk, and we have evaluated historical physical activity (PA) in a small cohort of genetically homogeneous ALS patients. Our study approach is summarised in **Figure 1**.

**Figure 1:**
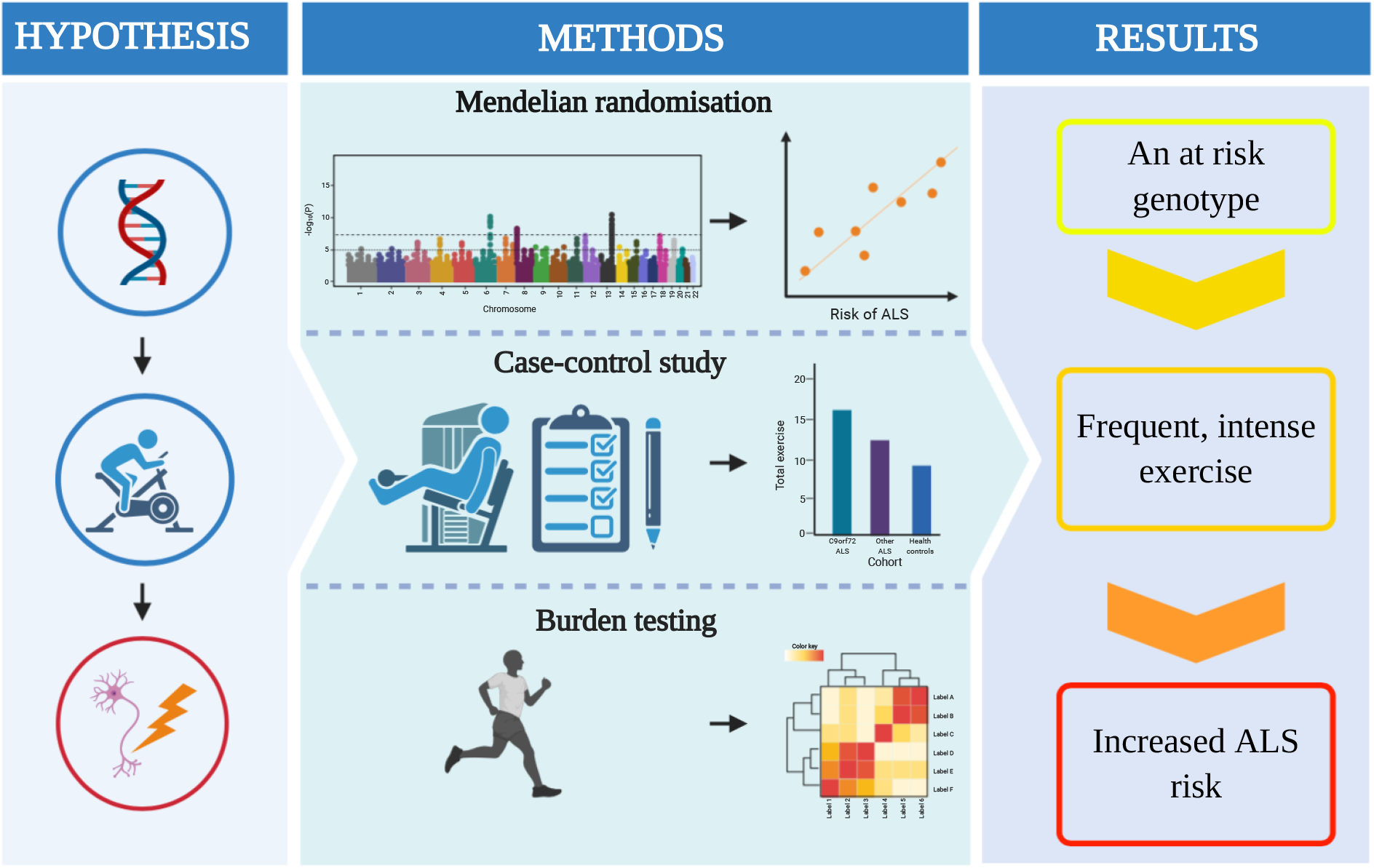
Exercise is a risk factor for amyotrophic lateral sclerosis: Convergent evidence from mendelian randomisation, transcriptomics and risk genotypes. A graphical abstract describing the fundamental elements of the study and core results. ALS = amyotrophic lateral sclerosis; PA = physical activity; SNP = single nucleotide polymorphism; GWAS = genome wide association study.

## RESULTS

### Genetic liability to frequent, strenuous, leisure-time exercise is a risk factor for ALS

We hypothesised that genetic liability to exercise may be associated with ALS. Exercise is not a single exposure, but is heterogeneous both in terms of the activity performed and the metabolic, neuromuscular and cardiovascular consequences. Our previous work has indicated that leisure-time strenuous activity may be linked to risk of ALS; this is captured by a ‘strenuous sport and other exercise’ (SSOE) GWAS (**Methods**).^9,20^ Interestingly, occupational physical activity is negatively associated with the SSOE measure suggesting that this is a relatively specific measure of frequent and intense leisure-time activity.^20^ As a control we have employed a GWAS utilising an accelerometer study of total movement over a limited period.

We discovered that genetic liability to SSOE is positively associated with ALS (IVW p=0.01, beta=0.21, **Table 1, Figure 2, Supplementary Table 1**). There was no statistically significant heterogeneity or directional pleiotropy and the F-statistic indicates adequate instrument strength. MR-PRESSO global test and leave-one-out analysis did not demonstrate any SNP outliers (**Table 2**). Robust tests consistently reached or trended towards significance. This result is consistent with a causative relationship between exercise and ALS which is not confounded by selection or recall bias. This result suggests that the association is driven primarily by a deleterious effect of exercise on motor neuron health and not by horizontal pleiotropy, i.e. our data are not consistent with a common genotype which independently influences both exercise and ALS (**Methods**).

**Table 1:** Two-sample mendelian randomisation demonstrates that strenuous sport and other exercise (SSOE) is a risk factor for ALS.

| **Mendelian randomisation method** | **Beta** | **Standard error** | **p value** |
| --- | --- | --- | --- |
| Inverse variance weighted (fixed effects) | 0.21 | 0.08 | 0.007 |
| Inverse variance weighted (multiplicative random effects) | 0.21 | 0.08 | 0.01 |
| MR Egger | 0.75 | 0.41 | 0.07 |
| Weighted median | 0.22 | 0.11 | 0.05 |
| Weighted mode | 0.52 | 0.25 | 0.04 |

**Table 2:**
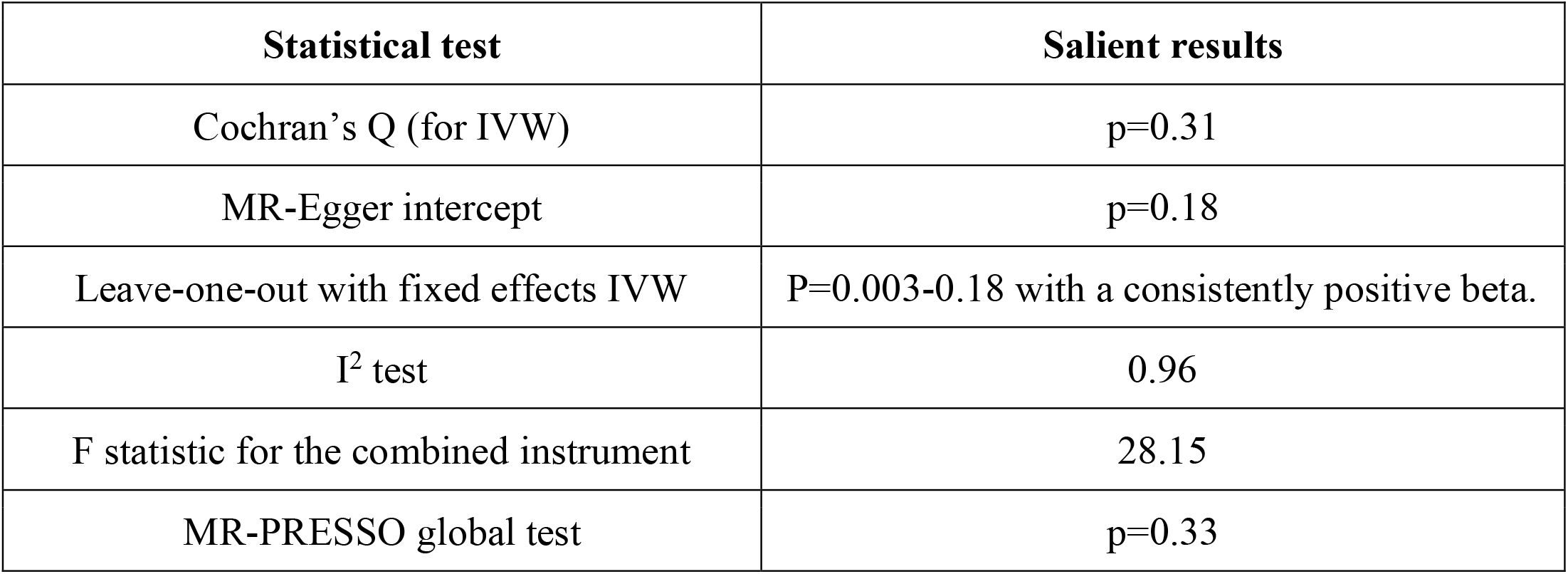
Two-sample mendelian randomisation for the effect of strenuous sport and other exercise (SSOE) on risk of ALS consists of robust instrumental variables.

| **Statistical test** | **Salient results** |
| --- | --- |
| Cochran's Q (for IVW) | p=0.31 |
| MR-Egger intercept | p=0.18 |
| Leave-one-out with fixed effects IVW | P=0.003-0.18 with a consistently positive beta. |
| I<sup>2</sup> test | 0.96 |
| F statistic for the combined instrument | 28.15 |
| MR-PRESSO global test | p=0.33 |

**Figure 2:**
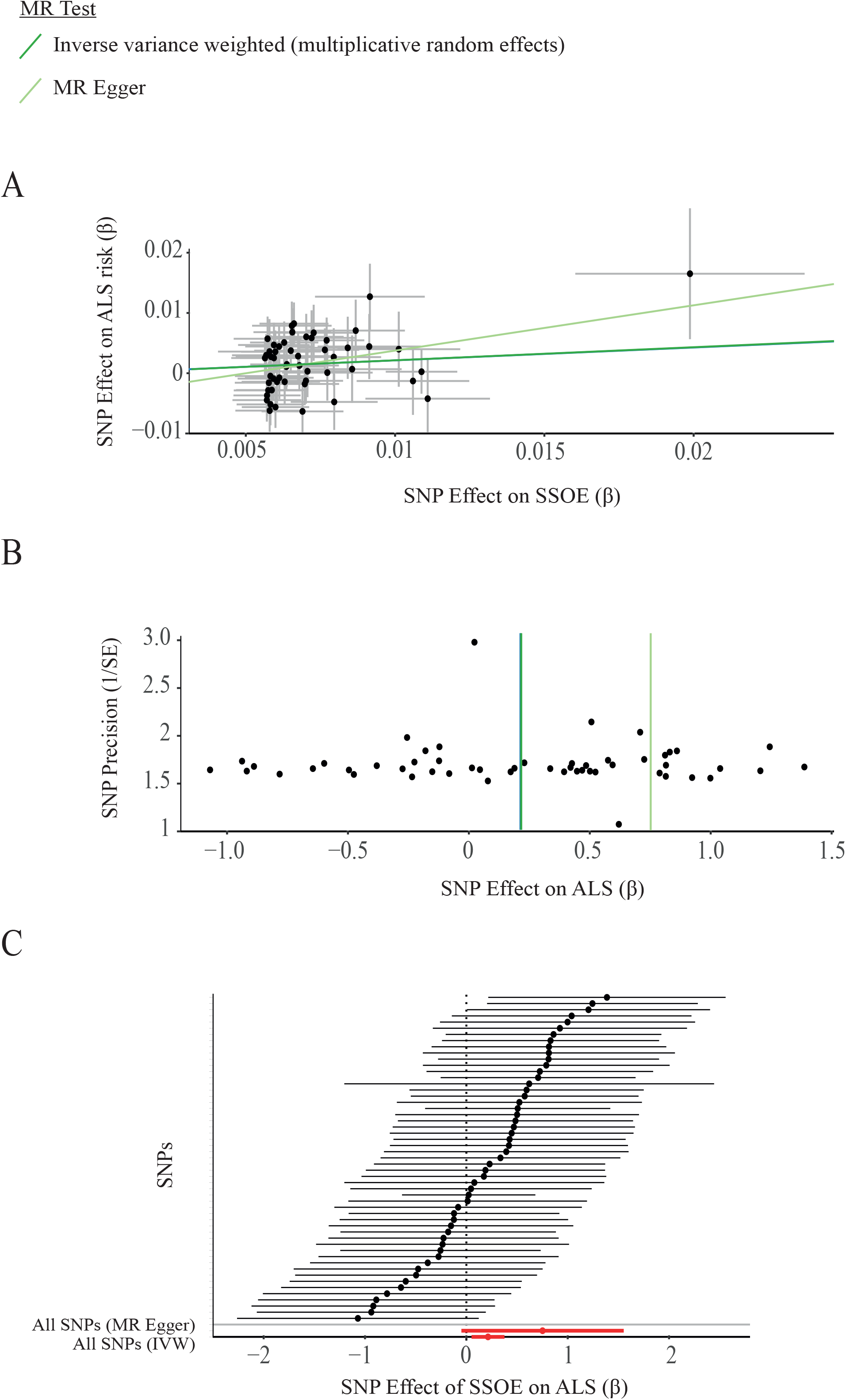
Genetic liability to frequent, strenuous, leisure-time exercise is a risk factor for ALS. **(A)** Scatter plot demonstrating the positive correlation between genetic liability to strenuous sport and other exercise (SSOE) and ALS, as measured with a liberal instrument. Points indicate effect size (β) and standard errors for each SNP-outcome relationship (i.e. ALS and SSOE). The relationship is not significantly altered by removal of any single SNP. The weighted median estimator is not significantly different to the IVW (β = 0.22 and 0.21 respectively) and therefore cannot be independently visualised. **(B)** A symmetrical funnel plot (vertical line of best fit) demonstrates that SNP effect size (β) is not correlated with SNP precision i.e. inaccurate instruments are not overvalued which could lead to directional pleiotropy (**C)** Forest plot illustrates that the effect of SSOE on ALS is consistent whether measured by individual SNPs or by MR Egger (upper red line) or IVW (lower red line). The overlapping confidence intervals of each causal estimate show there is no significant heterogeneity.

Movement alone was not significantly associated with ALS, which is consistent with our hypothesis that ALS is linked to strenuous exercise. MR determined a non-significant association with ALS for fraction of accelerations >425 milligravities (IVW, p=0.13, beta=-0.11, **Supplementary Table 2, Supplementary Figure 1**) and average accelerations (IVW, p=0.31, beta=-0.005, **Supplementary Table 3, Supplementary Figure 2**).

### Sedentary behaviour is not significantly associated with ALS

We argued that if strenuous exercise is a risk factor for ALS, then sedentary behaviour may be protective. However, our MR study does not support this conclusion. Sedentary behaviour is not significantly related to ALS (IVW p=0.29, beta=-0.04, **Supplementary Table 7, Supplementary Figure 3**).

### Association between physical exercise and ALS is not mediated by body fat percentage

Body fat percentage has previously been described as a potential risk factor for ALS.^21^ If true this would represent a possible source of pleiotropy or confounding in the SSOE-ALS relationship. Our analysis demonstrates that body fat percentage is not significantly associated with ALS (fixed effects IVW, p=0.43, beta=-0.03, **Supplementary Table 5, Supplementary Figure 4**) and is therefore unrelated to the pathophysiological impact of SSOE. This finding is in keeping with a previous MR which concluded that there is no causal relationship between BMI and ALS. ^22^

### Association between exercise and ALS is not mediated by educational attainment

The relationship between years of education and ALS was measured to identify whether this variable might have been a confounder for the SSOE-ALS relationship. Previous studies have demonstrated that leisure-time SSOE is positively correlated with educational attainment suggesting that our result could be confounded by a link between lower educational attainment and increased risk of ALS.^20^ Due to the large number of SNPs identified at genome-wide significance which were eligible for analysis (n=298), only a conservative instrument was used. This demonstrated no significant relationship between ALS and educational attainment (fixed effects IVW p=0.64, beta=-0.01, **Supplementary Table 6, Supplementary Figure 5**).

### Exercise-induced pathways are enriched with ALS genetic risk

Our results suggest that physical exercise is a risk factor for ALS. Next we set out to identify risk genotypes for exercise-induced ALS using transcriptomics. We hypothesised that risk genes for exercised-induced ALS should be differentially expressed in response to exercise. This could provide a functional link whereby exercise potentially amplifes toxicity resulting from a genetic mutation. We measured gene expression changes in peripheral blood mononuclear cells (PBMCs) in response to exercise and identified 323 biological pathways are differentially expressed in response to acute exercise including ‘ALS signaling’.^23^ We tested for enrichment of ALS genetic risk within each of these pathways by rare variant burden testing utilising whole-genome sequencing (WGS) data from 4,495 sporadic ALS patients and 1,925 controls (**Methods** and ^24^). Strikingly 72 pathways (22%) are significantly enriched with ALS-associated rare variants after multiple testing correction (FDR<0.05, **Figure 3A, Supplementary table 8**) including the ‘ALS signaling’ pathway (p=0.0007). One pathway was significantly enriched with ALS-associated rare variants even after a stringent Bonferroni multiple testing correction: fibroblast growth factor (FGF) signaling (p=0.0001, **Figure 3B**). Nerve growth factor (NGF) signaling is closely related to FGF signalling and is also highly enriched with ALS-associated rare variants (p=0.0002, **Figure 3B**).

**Figure 3:**
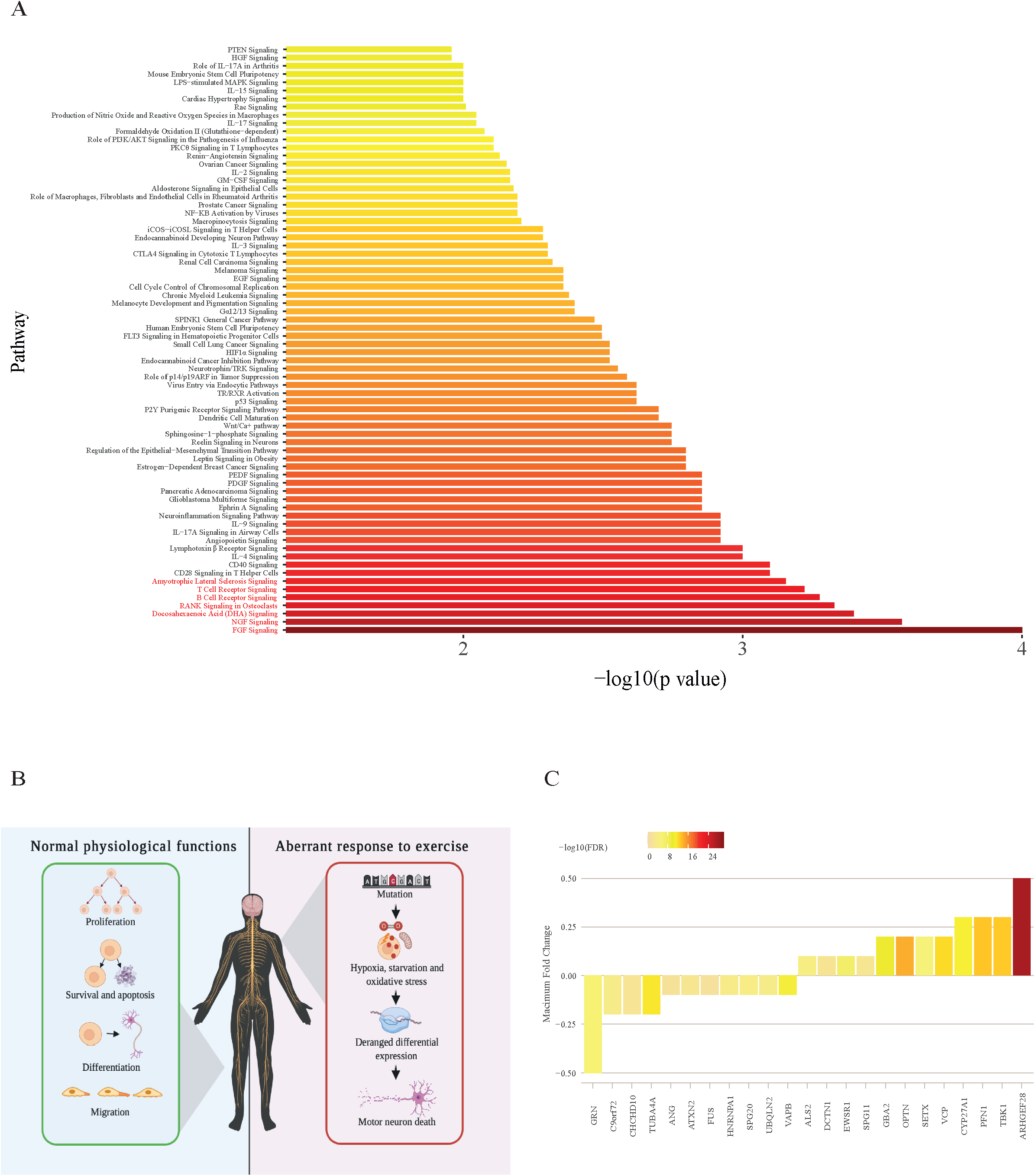
Exercise-induced pathways are enriched with ALS genetic risk. **(A)** Transcriptome analysis of peripheral blood mononuclear cells (PBMCs) reveals that biological pathways differentially expressed following acute exercise are significantly enriched with ALS-associated rare variants; pathways depicted pass multiple testing correction (FDR<0.05). **(B)** Differentially expressed biological pathways are closely related to neuronal health. Broadly, the roles of FGF and NGF pathways are cell proliferation, apoptosis and cell survival, cell differentiation and cell migration. The NGF and FGF pathways are significantly related to ALS and are enriched following acute exercise. In ALS, it is possible that mutation in these pathways leads to deranged differential expression in response to hypoxia, oxidative stress and starvation during exercise and this may precipitate damage to motor neurons. **(C)** ALS risk genes which are differentially expressed in response to exercise are shown with fold change and significance.

We hypothesised that, if exercise-induced gene expression changes are enriched with ALS risk, then known ALS genes should also be differentially expressed following exercise.^23^ Consistent with our hypothesis 52% of clinically validated ALS-related genes are differentially expressed following acute exercise and this enrichment is statistically significant (p<0.01, **Figure 3C, Supplementary Table 9, Methods**). The list of differentially expressed genes included down- regulation of *C9ORF72* (fold change=-0.2, FDR=0.0002).

### Case-control study of *C9ORF72*-ALS suggests an exercise threshold linked to penetrance

To investigate the possible gene-environment interaction further, we studied historical physical activity (PA) using the validated HAPAQ questionnaire^25^ in a cohort of *C9ORF72-*ALS patients (n=18) compared to age and sex matched non-*C9ORF72*-ALS patients (n=36) and neurologically normal controls (n=36) (**Table 4**). We proposed that variable disease penetrance in carriers of a G4C2-repeat expansion of *C9ORF72* is in part due to differences in exercise history. In our model, an individual carrying a *C9ORF72* expansion is likely to develop ALS only when they receive a certain ‘dose’ of exercise. Consistent with our model, age of ALS onset was negatively correlated with average daily PA in *C9ORF72*-ALS patients (linear regression, t=-2.28, r=-0.49, p=0.036, **Figure 4A**). The same was not true for non-C9ORF72-ALS patients (linear regression, t=-1.39, r=-0.23, p=0.17). Our model predicts a higher minimum level of PA in the group of *C9ORF72*- ALS patients compared to non-*C9ORF72*-ALS patients and neurologically normal controls which matched our observations (**Figure 4B**); the observed minimum average daily PA for *C9ORF72*- ALS patients was 20.9kJ/kg/day which may represent a threshold for disease penetrance. Variability in average PA was lower in *C9ORF72*-ALS compared to both non*-C9ORF72*-ALS (F-test, p=0.001) and neurologically normal controls (F-test, p=0.04). Maximum average daily PA was lowest in *C9ORF72*-ALS patients which could be the result of an attenuation due to pre-symptomatic deleterious effects of exercise occurring above the threshold for penetrance. Expressing the average daily PA per individual as a proportion of the minimum value for each group demonstrates a significant difference between the *C9ORF72*-ALS group and the non- *C9ORF72* ALS controls (t-test, p=1.1×10^−6^) and the neurologically normal controls (t-test, p=0.002).

**Table 3:** Two-sample mendelian randomisation demonstrates that strenuous sport and other exercise (SSOE) leads to reduced body fat percentage. The liberal instrument appears to produce more precise estimates with smaller standard errors and greater power, due to the larger number of SNPs.

| Positive control instrument | Multiplicative random effects IVW beta | Fixed effects IVW p-value | Multiplicative random effects IVW p-value | Weighted median p-value | MR-Egger p-value |
| --- | --- | --- | --- | --- | --- |
| **Conservative (p<5E-08)** | -0.78 | 0.003 | 0.01 | 0.23 | 0.97 |
| **Liberal (p<1E-06)** | -0.64 | 0.000004 | 0.00002 | 0.03 | 0.85 |

**Table 4:** Clinical characteristics of *C9ORF72*-ALS cases and controls from case-control analysis.

|  | **C9ORF72-ALS** | **Non-C9ORF72-ALS** | **Neurologically Normal Controls** |
| --- | --- | --- | --- |
| Number in group | 18 | 36 | 36 |
| Proportion male (%) | 58.8 | 58.8 | 58.5 |
| Age (years) | 56.6 (36-71) | 57.1 (35-72) | 57 (36-74) |
| ALS (%) | 15 (88.2) | 34 (100) | - |
| PMA (%) | 2 (11.8) | 0 | - |
| PLS (%) | 0 | 0 | - |

ALS = amyotrophic lateral sclerosis; PMA= progressive muscular atrophy variant; PLS= primary lateral sclerosis variant.

**Figure 4:**
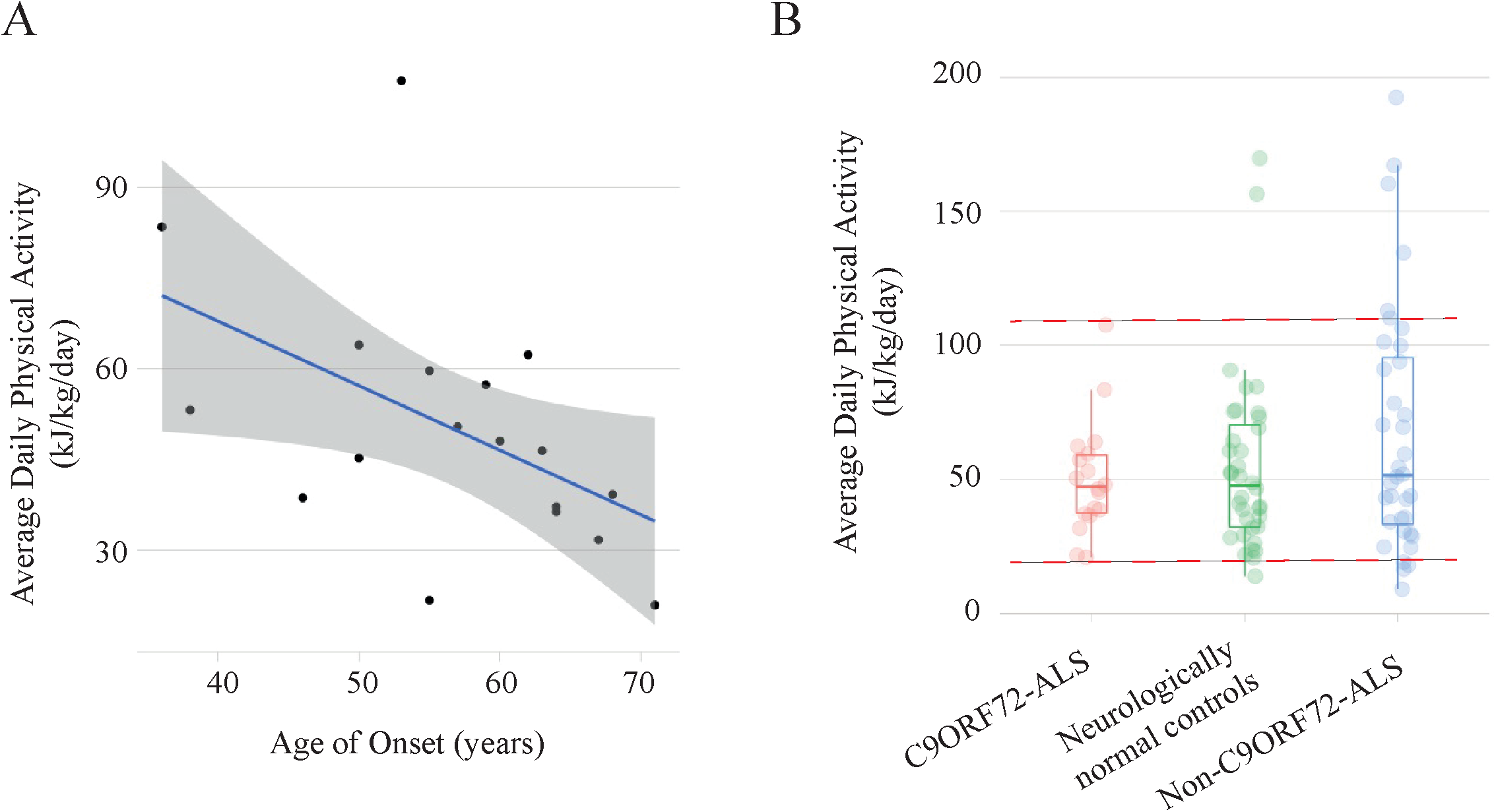
Case-control study of *C9ORF72*-ALS suggests a gene-environment interaction with physical activity (PA). **(A)** Historical PA measured by the validated HAPAQ questionnaire is inversely related to age of disease onset in C9ORF72-ALS (p<0.05, r=-0.47); line of best-fit shown with 95% confidence interval. **(B)** Measured historical PA is less variable in C9ORF72-ALS compared to non- C9ORF72-ALS patients and neurologically normal controls; the upper and lower limits of measured PA in *C9ORF72*-ALS (red dotted lines) are significantly attenuated consistent with a threshold effect.

## DISCUSSION

ALS is an archetypal complex disease determined by interaction between genetic risk factors and environmental modifiers.^2^ Understanding this gene-environment interaction will lead to new personalised medicine approaches. For rare forms of ALS which are purely genetically determined, this is already a reality.^26^ It is likely that the search for environmental risk factors has been hindered by studies which have failed to account for heterogeneity in both environmental exposures and ALS genetic subtypes: specific exposures are likely to be relevant only in the presence of specific genotypes. We have used two-sample MR to establish the basis for strenuous leisure-time exercise as a risk factor for ALS and we have also developed understanding of the specific genetic subtypes of ALS which may be responsible for this interaction.

The link between exercise and ALS risk has previously been controversial.^8,9^ The advantage of a two-sample MR methodology is that it can avoid much of the bias and confounding factors which have hindered previous analyses. Moreover, exercise is not one homogeneous exposure; in reality different types of exercise can impact different biological pathways and even different motor neurons. Early in the disease process ALS is known to selectively affect motor neurons supplying type IIb fast twitch muscle fibres important for high intensity anaerobic exercise.^16,17^ Consistent with this, our MR study does not support a causal role for low-intensity, infrequent exercise but does support toxicity resulting from high-intensity, frequent, leisure-time exercise. The SSOE measure of exercise we utilised is negatively correlated with occupational activity suggesting that it is a specific measure of strenuous leisure-time activity.^20^ Variations in the cardiovascular impact of occupational and leisure time activity are thought to represent more static occupational activity versus more dynamic leisure-time activity.^19,27^ This suggests that intense leisure-time activity is more likely to include strenuous anaerobic activity and might explain why this type of physical activity shows a strong association with ALS.

We discovered that ALS risk genes are differentially expressed following exercise and similarly, a significant number of exercise-induced biological pathways are enriched with ALS genetic risk. This provides a functional link between exercise and the activity of ALS risk genes. Many of these pathways have previously been linked to neurotoxicity, but our work places them upstream in the pathogenesis of exercise-induced ALS. Most prominently genes linked to fibroblast growth factor (FGF) and nerve growth factor (NGF) signaling were highly enriched with ALS-associated rare variants. FGF1 is known to induce NGF expression in astrocytes and therefore these represent related pathways.^28^ FGFs are highly expressed in motor neurons and FGF1 secretion can be stimulated by oxidative stress, hypoxia and serum starvation.^28^ Produced by astrocytes, both NGF and FGF have been shown to cause motor neuron apoptosis under specific conditions *in vitro* and some authors have proposed that this may be implicated in ALS pathophysiology.^28,29^ Additionally, NGF levels are increased in the muscle of ALS patients. ^29^ We suggest that both of these pathways, and the other pathways and genes we have highlighted, should be explored in detail to define a risk genotype for exercise-induced ALS.

G4C2-repeat expansion of *C9ORF72* is the most common genetic risk factor for ALS, but the phenotype is extremely variable and penetrance is incomplete suggesting a role for environmental modifiers.^14^ We hypothesised that exercise may modify the penetrance of *C9ORF72*-expansions. Consistent with this we have shown that *C9ORF72* is down-regulated during exercise, which could act synergistically with haploinsufficiency caused by the expansion, so enhancing neurotoxicity.^30,31^ Secondly we have shown that age of onset in *C9ORF72*-ALS, but not non- *C9ORF72*-ALS, is correlated with the ‘dose’ of historical physical activity. Finally, our data are consistent with a link between *C9ORF72* penetrance and exercise: *C9ORF72*-ALS patients with <20.9kJ/kg/day activity throughout adult life were notably absent from our cohort, whereas they were present in both non-*C9ORF72*-ALS patients and neurologically normal controls. This threshold is equivalent to ∼45 minutes of vigorous exercise per day. We propose that these individuals are missing because they have not developed ALS. If this proposal is correct then this could lead to lifestyle recommendations and potentially disease prevention.

In conclusion, the current evidence supports a complex causal relationship between physical exercise and ALS. However, it is clear that, for the majority of individuals, the health benefits of a physically active lifestyle markedly outweigh the risks. The key objective for future research is to understand which individuals are at risk of developing ALS if they exercise excessively and provide appropriate lifestyle counselling. Our work goes some way to developing this aim and in particular, we propose that *C9ORF72* penetrance may be influenced by high levels of physical activity.

## Data Availability

Data available at the links URLs

http://databrowser.projectmine.com/

## ACKNOWLEDGEMENTS

This work was supported by research grants from the National Institutes of Health (CEGS 5P50HG00773504, 1P50HL083800, 1R01HL101388, 1R01-HL122939, S10OD025212, and P30DK116074, UM1HG009442 to MPS) and the Wellcome Trust (216596/Z/19/Z to JCK), a Kingsland Fellowship (TM), an NIHR Foundation Fellowship (THJ) and an NIHR Senior Investigator Award (NF-SI-0617-10077 to PJS). This work was also supported by the NIHR Sheffield Biomedical Research Centre for Translational Neuroscience (IS-BRC-1215-20017). We thank Dr Thomas Jenkins, Dr Channa Hewamadduma, Prof Christopher McDermott, Professor Martin Turner and Professor Kevin Talbot who referred ALS patients to our study. We are very grateful to those ALS patients and control subjects who generously donated the biosamples underpinning this study.

## AUTHOR CONTRIBUTIONS

PJS, JCK and THJ conceived and designed the study. THJ, NG, ADFB, YCK, MPS, JCK and PJS were responsible for data acquisition. THJ, NG, ADFB, YCK, MN, SZ, JCK and PJS were responsible for analysis of data. THJ, NG, ADFB, TM, YCK, MN, SZ, MPS, JCK and PJS were responsible for interpretation of data. THJ, JCK and PJS. prepared the manuscript with assistance from all authors. All authors meet the four ICMJE authorship criteria, and were responsible for revising the manuscript, approving the final version for publication, and for the accuracy and integrity of the work.

## SUPPLEMENTARY TABLES

**Supplementary Table 1:** Mendelian randomisation including both conservative and liberal analysis of the SSOE-ALS relationship. This table contains all data used to analyse the SSOE- ALS relationship, the sensitivity and instrument strength measures, and the results.

**Supplementary Table 2:** Mendelian randomisation including both the conservative and liberal analysis of the fraction of accelerations >425 milligravities - ALS relationship. This table contains all data used to analyse the accelerations >425 milligravities - ALS relationship, the sensitivity and instrument strength measures, and the results.

**Supplementary Table 3:** Mendelian randomisation including both the conservative and liberal analysis of the average accelerations-ALS relationship. This table contains all data used to analyse the average accelerations - ALS relationship, the sensitivity and instrument strength measures, and the results.

**Supplementary Table 4:** Mendelian randomisation for the positive control data with liberal and conservative SSOE instruments which assesses the strength of instruments to identify a negative association with body fat percentage. This table includes all data used to perform the analysis as well as the evaluations of instrument strength and the results.

**Supplementary Table 5:** Mendelian randomisation including both the conservative and liberal analysis of the body fat percentage-ALS relationship. This table contains all data used to analyse the body fat percentage - ALS relationship, the sensitivity and instrument strength measures, and the results.

**Supplementary Table 6:** Mendelian randomisation including both the conservative analysis of the educational attainment-ALS relationship.This table contains all data used to analyse the educational attainment - ALS relationship, the sensitivity and instrument strength measures, and the results.

**Supplementary Table 7:** Mendelian randomisation for the conservative and liberal analysis of the sedentary behaviour- ALS relationship. This table includes all data used to perform the analysis as well as the evaluations of instrument strength and the results.

**Supplementary Table 8:** Burden testing results which described the significance of pathways which are differentially expressed during exercise in an ALS cohort. The results are delivered with respect to significance of the pathways and do not focus upon the significance of individual genes.

**Supplementary Table 9:** A table which described the differential expression of known ALS genes during exercise including their FDR and fold change at several time points post-exercise. Additionally, the full genetic panel which was included in the methods is provided.

## SUPPLEMENTARY FIGURES

**Supplementary Figure 1:** A) a scatter plot for the non-significant fraction of accelerations >425 milligravities - ALS relationship. B) a funnel plot for the fraction of accelerations >425 milligravities - ALS relationship.

**Supplementary Figure 2:** A) a scatter plot for the non-significant average accelerations - ALS relationship. B) a funnel plot for the average accelerations - ALS analysis.

**Supplementary Figure 3:** A) a scatter plot for the non-significant sedentary behaviour-ALS relationship. B) a funnel plot for the sedentary behaviour - ALS analysis.

**Supplementary Figure 4:** A) a scatter plot for the non-significant body fat % - ALS relationship. B) a funnel plot for the body fat % - ALS analysis.

**Supplementary Figure 5:** A) a scatter plot for the non-significant educational attainment - ALS relationship. B) a funnel plot for the educational attainment -ALS analysis,

## METHODS

### Two-sample mendelian randomisation (MR)

#### Objective

In MR, SNPs associated with an exposure of interest are used as instrumental variables (IV) to explore the causal relationship between that exposure and an outcome of interest. According to Mendel’s laws of segregation and independent assortment, this methodology can be considered a natural experiment in which individuals are at conception randomly assigned to groups with differing levels of genetic liability to an exposure of interest. A core strength of MR is the ability to remove unmeasured confounding and to differentiate between a shared genetic basis for the exposure and outcome (known as genetic pleiotropy), and potential causation. By utilising SNPs associated with various forms of exercise, we are therefore able to interrogate the relationship between exercise and ALS. SNPs which make up the IV are usually selected on the basis of genome wide association studies (GWAS) used to assess the significance of the relationship between specific genetic variants and an exposure of interest. ^32^

#### Study participants

In this study, the instrumental variables (IV) were derived from publicly available GWAS. None of the exposure GWAS participants overlapped with those in the outcome GWAS (i.e. ALS) which is important to avoid bias.

SSOE was measured via questionnaire in in UK Biobank (UKB) participants.^20^ Individuals completing the physical activity questionnaire were also asked “In the last 4 weeks did you spend any time doing the following?” with the potential answers were: ‘walking for pleasure’, ‘other exercises’, ‘strenuous sports’, ‘light DIY’, ‘heavy DIY’, ‘none of the above’, and ‘prefer not to answer’. To address the question of which SNPs were associated with participation in “strenuous sports” and “other exercise” (SSOE) the responses were dichotomised to those who reported spending two-three days per week or more performing SSOE for a duration of 15-30 minutes or greater, and those who did not. A total of 124,842 cases and 225,650 controls contributed to the SSOE GWAS. The data were analysed with a linear mixed model analysis.

Accelerometer data was measured by a Axivity AX3 wrist-worn accelerometer which was worn by UKB subjects for one week, with GWAS linear mixed model analysis conducted on the basis of three measures : 1) overall average acceleration, 2) fraction of accelerations > 425 milligravities, 3) sedentary behaviour as previously defined.^20,33^ 91,084 individuals data contributed to the average accelerations analysis, 90,667 to the GWAS of fraction of accelerations >425 milligravities and 91,105 to the GWAS of sedentary behaviour.^33^

Body fat was measured by dual energy x-ray absorptiometry or bioimpedance analysis.^34^ Body fat GWAS was performed as a meta-analysis and included 100,716 individuals.

Educational attainment was quantified as years of education measured at an age of at least 30 years; GWAS was performed in a meta-analysis of 1,131,881 individuals.^35^

ALS GWAS was performed utilising a linear mixed model as previously described.^36^ This is the largest published ALS GWAS to date including 12,577 cases and 23,475 controls from 41 cohorts.

Limitations of GWAS datasets are discussed in the **Supplementary Note**.

#### Selection of exposure instrumental variables (IV)

Exposure IV are chosen based on an arbitrary p-value cut off.^37,38^ A cut-off which is too low will lose informative instruments, but a cut-off which is too high could introduce non-informative instruments. We chose to utilise a positive control to determine the optimum number of IV for our study of physical exercise and ALS. Body fat was chosen as a positive control with a clear and described biological mechanism linking increased exercise to reduced body fat. Our previous work has suggested that leisure-time strenuous activity may be linked to risk of ALS which is captured by the SSOE GWAS.^9^ We compared a liberal (p<1E-06) and conservative (p<5E-08) instrument to test the relationship between SSOE and body fat; both results were significant but the liberal instrument had a greater power (**Table 3, Supplementary Table 4**). Consequently, we have used a liberal instrument at all stages of our analysis.

Identified SNPs at each significance threshold were clumped for independence using PLINK clumping in TwoSampleMR.^39^ A stringent cut-off of R^2^ ≤ 0.001 and a window of 10,000kb was used for clumping within a European reference panel. Where SNPs were in linkage disequilibrium (LD), those with the lowest p-value were retained. SNPs which were not present in the reference panel were excluded. Where an exposure SNP was unavailable in the outcome dataset, a proxy with a high degree of LD (R^2^ ≥ 0.9) was identified in LDLink within a European reference population.^40^ Where a proxy was identified to be present in both datasets, the target SNP was replaced with the proxy in both exposure and outcome datasets in order to avoid phasing issues.^41^ Where a SNP was not present in both datasets and no SNP was available in sufficient LD, the SNP was excluded from the analysis. All SNPs selected for inclusion in this study are presented in the supplementary data in order to allow replication.

#### Exposure-outcome instrument harmonisation

The effects of SNPs on outcomes and exposures were harmonised in order to ensure that the beta values were signed with respect to the same alleles. For palindromic alleles, those with minor allele frequency (MAF) > 0.42 were omitted from the analysis in order to reduce the risk of errors due to strand issues.^41^

#### Assumptions and robust analysis

The MR measure with the greatest power is the inverse-variance weighted (IVW) method, but this is contingent upon the exposure IV assumptions being satisfied.^42^ With the inclusion of a large number of SNPs within the exposure IV, it is possible that not all variants included are valid instruments and therefore it is necessary to include a range of robust methods which provide valid results under various violations of MR principles at the expense of power.^32^ Robust methods applied in this study include MR-Egger, MR-PRESSO and weighted median. Where a causal relationship was suggested, we additionally applied the weighted mode.

With respect to the IVW analysis, a fixed-effects (FE) model is indicated in the case of homogeneous data whilst a multiplicative random effects (MRE) model is more suitable for heterogeneous data. Burgess *et al* recommend that an MRE model is implemented when using GWAS summary data to account for heterogeneity in variant-specific causal estimates.^32^ In the interest of transparency, we shall calculate both results but present the MRE in the text.

MR analyses should include evaluation of exposure IV strength. In order to achieve this, we provide the F-statistic, MR-Egger intercept, MR-PRESSO global test, Cochran’s Q test and I^2^ for our data. The F-statistic is a measure of instrument strength with >10 indicating a sufficiently strong instrument.^43^ We provide F-statistics for individual exposure SNPs and the instrument as a whole. Cochran’s Q test is an indicator of heterogeneity in the exposure dataset and serves as a useful indicator that horizontal pleiotropy is present as well as directing decisions to implement FE or MRE IVW approaches.^44^ The MR-Egger intercept test determines whether there is directional, horizontal pleiotropy. The MR-PRESSO global test determines if there are statistically significant outliers within the exposure-outcome analysis.^45^ I^2^ was calculated as a measure of heterogeneity between variant specific causal estimates, with a low I^2^ indicating that Egger is more likely to be biased towards the null.^46^ Finally, we performed a leave-one-out analysis using the method of best fit for each exposure SNP within the IV in order to determine if any single variants were exerting a disproportionate effect upon the results of our analysis.^32^

### Burden testing

Whilst GWAS is useful for exploring the genetic architecture of disease, this methodology does not capture low-frequency or rare variation ^47^ which is key to the genetic architecture of ALS. ^36^ Rare variants are addressed by rare variant burden testing.^48^ In this study, we grouped genes within pathways which changed expression in response to exercise. We measured the rate of observed rare mutations within exercise pathways in ALS patients compared to controls. By identifying ALS-associated genetic variation within pathways functionally related to acute exercise, we aimed to implicate specific genotypes in motor neuron vulnerability to PA.

Burden testing was performed using whole genome sequencing data from 4,425 ALS patients and 1,925 controls.^24^ We have previously identified genes and pathways differentially expressed in response to exercise.^23^ In brief 36 subjects underwent symptom-limited cardiopulmonary exercise (CPX) testing with whole transcriptome profiling by sequencing of RNA extracted from peripheral blood mononuclear cells at 2, 15, 30 and 60 minutes post exercise.^23^ For the purpose of the present research, pathways which were differentially expressed at the most immediate time point of 2 minutes post-CPX testing (p<0.001) were selected for burden testing. For each pathway we calculated the proportion of genes which were significantly enriched with rare (MAF<1%) missense ALS-associated mutations. To determine significant enrichment of each pathway with ALS-associated mutations we calculated the equivalent enrichment within 5,000-30,000 random gene sets of equivalent length. Reported p- and FDR values for pathway enrichment refer to the proportion of random gene sets with equivalent enrichment. To ensure stable p-value estimates the number of random gene sets was increased such that ≥3 random sets were discovered with equivalent enrichment to each pathway under consideration.

### Expression of ALS-related genes during exercise

In order to determine whether ALS-related genes are differentially expressed during physical exercise, we measured expression changes of known ALS genes post-exercise.^23^ We utilised a clinical ALS gene panel ([Next Generation Sequencing](https://www.sheffieldchildrens.nhs.uk/download/321/ngs/9291/next-generation-sequencing-v7.pdf) at Sheffield NHS Children’s Hospital, **Supplementary Table 9**).^49^ The proportion of ALS genes which were differentially expressed with exercise was compared to a 1,000 random gene sets of the same length.

### A *C9ORF72*-specific case-control study of historical PA

#### Study participants

ALS cases were identified by a Consultant Neurologist according to revised El-Escorial criteria. Relevant National Health Service (NHS) and university research ethical approvals were obtained (IRAS189432 / STH19120) and procedures followed were in accordance with these standards. All study participants provided informed consent. Cases with family history or other clinical features consistent with *C9ORF72*-disease such as young age of onset or extra-motor features underwent genetic testing for the G4C2-expansion. Inclusion criteria for the study were as follows: (1) Confirmed *C9ORF72* pathological G4C2-repeat expansion; (2) patients were over 18 years of age whose disease manifested in adult life, and (3) diagnosis had taken place within the last two years. Exclusion criteria were as follows: (1) patients with concurrent neurological disease; (2) patients with symptoms incompatible with completing the HAPAQ questionnaire such as overt cognitive impairment. Two control groups were used in this study: those with ALS but without the *C9ORF72* G4C2-expansion, and neurologically normal controls. Both groups were matched to *C9ORF72*- ALS patients for age and gender. Control data were obtained from a previous study.^9^ Participants in the study are summarised in **Table 5**.

#### Measurement of PA

PA was measured using the HAPAQ questionnaire which has been previously validated for the determination of historical PA.^25^ Briefly a life calendar was used to orientate participants and aid recall. The questionnaire is structured by time period, of which two broad lengths of time are covered (a) the most recent 15 years, split into three 5-year categories and (b) the whole of adulthood, by decade, starting from the age of 20 years. In each discrete time period a series of questions are asked about physical activity within four distinct domains: (a) in and around the home; (b) at work or commuting to work; (c) physical activity that makes you out of breath/sweat and (d) physical activity that does not make you out of breath or sweat. For each domain, closed questions are asked about the type, duration, and frequency of the PA.

#### Statistical Analysis

Every unique activity was assigned a MET value according to the Compendium of Physical Activities.^50^ Based on these values we calculated a measure of the average daily physical activity in kJ/kg/day for each patient during each time period which accounted for type, duration, intensity, and frequency of PA. To compare between subjects an overall average daily physical activity value was calculated for the most recent 20 years excluding the most recent 5 years which may have been confounded by subclinical symptom onset.

### Software

The TwoSampleMR (version 0.5.5) package in R (version 4.0.2) was used to perform mendelian randomisation.^51^ Proxy SNPs for mendelian randomisation were identified using the LDLinkR (version 1.0.2) package.^52^ The code which we utilised for the more statistically complex aspects of the work (MR and burden analysis) is provided in the **Supplementary Note**. R language was also utilised for burden testing and various basic calculations, performed using base R and dplyr functionality. The statistics program STATA/IC (version 15.0) was used for statistical analysis of case-control data.

## REFERENCES

1 Brown RH, Al-Chalabi A. Amyotrophic Lateral Sclerosis. N Engl J Med 2017; 377 : 162–72.

2 Cooper-Knock J, Jenkins T, Shaw PJ. Clinical and Molecular Aspects of Motor Neuron Disease. Biota Publishing, 2013.

3 Ryan M, Heverin M, McLaughlin RL, Hardiman O. Lifetime Risk and Heritability of Amyotrophic Lateral Sclerosis. JAMA Neurol 2019; published online July 22. DOI :10.1001/jamaneurol.2019.2044.

4 Trabjerg BB, Garton FC, van Rheenen W, et al. ALS in Danish Registries: Heritability and links to psychiatric and cardiovascular disorders. Neurol Gene t 2020; 6 : e398.

5 Al-Chalabi A, Calvo A, Chio A, et al. Analysis of amyotrophic lateral sclerosis as a multistep process: a population-based modelling study. Lancet Neurol 2014; 13 : 1108–13.

6 Chiò A, Mazzini L, D’Alfonso S, et al. The multistep hypothesis of ALS revisited: The role of genetic mutations. Neurology 2018; 91 : e635–42.

7 Chio A, Calvo A, Dossena M, Ghiglione P, Mutani R, Mora G. ALS in Italian professional soccer players: the risk is still present and could be soccer-specific. Amyotroph Lateral Scler 2009; 10 : 205–9.

8 Hamidou B, Couratier P, Besançon C, Nicol M, Preux PM, Marin B. Epidemiological evidence that physical activity is not a risk factor for ALS. European Journal of Epidemiology. 2014; 29 : 459–75.

9 Harwood CA, Westgate K, Gunstone S, et al. Long-term physical activity: an exogenous risk factor for sporadic amyotrophic lateral sclerosis? Amyotroph Lateral Scler Frontotemporal Degener 2016; 17 : 377–84.

10 Lacorte E, Ferrigno L, Leoncini E, Corbo M, Boccia S, Vanacore N. Physical activity, and physical activity related to sports, leisure and occupational activity as risk factors for ALS: A systematic review. Neuroscience & Biobehavioral Reviews. 2016; 66 : 61–79.

11 Bandres-Ciga S, Noyce AJ, Hemani G, et al. Shared polygenic risk and causal inferences in amyotrophic lateral sclerosis. Ann Neurol 2019; 85 : 470–81.

12 DeJesus-Hernandez M, Mackenzie IR, Boeve BF, et al. Expanded GGGGCC hexanucleotide repeat in noncoding region of C9ORF72 causes chromosome 9p-linked FTD and ALS. Neuron 2011; 72 : 245–56.

13 Cooper-Knock J, Hewitt C, Highley JR, et al. Clinico-pathological features in amyotrophic lateral sclerosis with expansions in C9ORF72. Brain 2012; 135 : 751–64.

14 Majounie E, Renton AE, Mok K, et al. Frequency of the C9orf72 hexanucleotide repeat expansion in patients with amyotrophic lateral sclerosis and frontotemporal dementia: a cross-sectional study. Lancet Neurol 2012; 11 : 323–30.

15 Karpati G, Hilton-Jones D, Griggs RC, Bushby K. Disorders of Voluntary Muscle. Cambridge University Press, 2008.

16 Nijssen J, Comley LH, Hedlund E. Motor neuron vulnerability and resistance in amyotrophic lateral sclerosis. Acta Neuropathol 2017; 133 : 863.

17 Ragagnin AMG, Shadfar S, Vidal M, Jamali MS, Atkin JD. Motor Neuron Susceptibility in ALS/FTD. Front Neurosci 2019; 13 : 532.

18 Huisman MHB, Seelen M, de Jong SW, et al. Lifetime physical activity and the risk of amyotrophic lateral sclerosis. J Neurol Neurosurg Psychiatry 2013; 84 : 976–81.

19 Clays E, De Bacquer D, Van Herck K, De Backer G, Kittel F, Holtermann A. Occupational and leisure time physical activity in contrasting relation to ambulatory blood pressure. BMC Public Health 2012; 12 : 1002.

20 Klimentidis YC, Raichlen DA, Bea J, et al. Genome-wide association study of habitual physical activity in over 377,000 UK Biobank participants identifies multiple variants including CADM2 and APOE. Int J Obes 2018; 42 : 1161–76.

21 Mariosa D, Beard JD, Umbach DM, et al. Body Mass Index and Amyotrophic Lateral Sclerosis: A Study of US Military Veterans. Am J Epidemiol 2017; 185 : 362–71.

22 Zeng P, Yu X, Xu H. Association Between Premorbid Body Mass Index and Amyotrophic Lateral Sclerosis: Causal Inference Through Genetic Approaches. Front Neurol 2019; 10 : 543.

23 Contrepois K, Wu S, Moneghetti KJ, et al. Molecular Choreography of Acute Exercise. Cell 2020; 181 : 1112–30.e16.

24 van der Spek RAA, Van Rheenen W, Pulit SL, et al. The Project MinE databrowser: bringing large-scale whole-genome sequencing in ALS to researchers and the public. Amyotroph Lateral Scler Frontotemporal Degener 2019; 20 : 432–40.

25 Besson H, Harwood CA, Ekelund U, et al. Validation of the historical adulthood physical activity questionnaire (HAPAQ) against objective measurements of physical activity. Int J Behav Nutr Phys Act 2010; 7 : 54.

26 Miller T, Cudkowicz M, Shaw PJ, et al. Phase 1–2 Trial of Antisense Oligonucleotide Tofersen for SOD1 ALS. New England Journal of Medicine. 2020; 383 : 109–19.

27 Holtermann A, Krause N, van der Beek AJ, Straker L. The physical activity paradox: six reasons why occupational physical activity (OPA) does not confer the cardiovascular health benefits that leisure time physical activity does. British Journal of Sports Medicine. 2018; 52 : 149–50.

28 Cassina P, Pehar M, Vargas MR, et al. Astrocyte activation by fibroblast growth factor-1 and motor neuron apoptosis: implications for amyotrophic lateral sclerosis. J Neurochem 2005; 93 : 38–46.

29 Barbeito LH, Pehar M, Cassina P, et al. A role for astrocytes in motor neuron loss in amyotrophic lateral sclerosis. Brain Res Brain Res Rev 2004; 47 : 263–74.

30 Cooper-Knock J, Higginbottom A, Connor-Robson N, et al. C9ORF72 transcription in a frontotemporal dementia case with two expanded alleles. Neurology 2013; 81 : 1719–21.

31 Cooper-Knock J, Shaw PJ, Kirby J. The widening spectrum of C9ORF72-related disease; genotype/phenotype correlations and potential modifiers of clinical phenotype. Acta Neuropathol 2014; 127 : 333–45.

32 Burgess S, Smith GD, Davies NM, et al. Guidelines for performing Mendelian randomization investigations. Wellcome Open Research 2019; 4. DOI :10.12688/wellcomeopenres.15555.2.

33 Doherty A, Smith-Byrne K, Ferreira T, et al. GWAS identifies 14 loci for device-measured physical activity and sleep duration. Nat Commun 2018; 9 : 5257.

34 Lu Y, Day FR, Gustafsson S, et al. New loci for body fat percentage reveal link between adiposity and cardiometabolic disease risk. Nat Commun 2016; 7 : 10495.

35 Lee JJ, Wedow R, Okbay A, et al. Gene discovery and polygenic prediction from a genome-wide association study of educational attainment in 1.1 million individuals. Nat Genet 2018; 50 : 1112–21.

36 van Rheenen W, Shatunov A, Dekker AM, et al. Genome-wide association analyses identify new risk variants and the genetic architecture of amyotrophic lateral sclerosis. Nat Genet 2016; 48 : 1043–8.

37 Choi KW, Chen C-Y, Stein MB, et al. Assessment of Bidirectional Relationships Between Physical Activity and Depression Among Adults: A 2-Sample Mendelian Randomization Study. JAMA Psychiatry 2019; 76 : 399–408.

38 Wootton RE, Lawn RB, Millard LAC, et al. Evaluation of the causal effects between subjective wellbeing and cardiometabolic health: mendelian randomisation study. BMJ 2018; 362 : k3788.

39 Purcell S, Neale B, Todd-Brown K, et al. PLINK: a tool set for whole-genome association and population-based linkage analyses. Am J Hum Genet 2007; 81 : 559–75.

40 Machiela MJ, Chanock SJ. LDlink: a web-based application for exploring population-specific haplotype structure and linking correlated alleles of possible functional variants. Bioinformatics 2015; 31 : 3555–7.

41 Hartwig FP, Davies NM, Hemani G, Davey Smith G. Two-sample Mendelian randomization: avoiding the downsides of a powerful, widely applicable but potentially fallible technique. Int J Epidemiol 2016; 45 : 1717–26.

42 Burgess S, Thompson SG. Erratum to: Interpreting findings from Mendelian randomization using the MR-Egger method. Eur J Epidemiol 2017; 32 : 391–2.

43 Burgess S, Thompson SG, CRP CHD Genetics Collaboration. Avoiding bias from weak instruments in Mendelian randomization studies. Int J Epidemiol 2011; 40 : 755–64.

44 Bowden J, Hemani G, Smith GD. Invited Commentary: Detecting Individual and Global Horizontal Pleiotropy in Mendelian Randomization—A Job for the Humble Heterogeneity Statistic? American Journal of Epidemiology. 2018. DOI :10.1093/aje/kwy185.

45 Verbanck M, Chen C-Y, Neale B, Do R. Publisher Correction: Detection of widespread horizontal pleiotropy in causal relationships inferred from Mendelian randomization between complex traits and diseases. Nat Genet 2018; 50 : 1196.

46 Bowden J, Del Greco M. F, Minelli C, Smith GD, Sheehan NA, Thompson JR. Assessing the suitability of summary data for two-sample Mendelian randomization analyses using MR-Egger regression: the role of the I2 statistic. International Journal of Epidemiology. 2016; : dyw220.

47 Lee S, Abecasis GR, Boehnke M, Lin X. Rare-variant association analysis: study designs and statistical tests. Am J Hum Genet 2014; 95 : 5–23.

48 Cirulli ET, Goldstein DB. Uncovering the roles of rare variants in common disease through whole-genome sequencing. Nat Rev Genet 2010; 11 : 415–25.

49 Sheffield Children’s NHS Foundation Trust. Next Generation Sequencing Service at SDGS. Sheffield Children’s NHS Foundation Trust. 2019; published online Nov 18. https://www.sheffieldchildrens.nhs.uk/download/321/ngs/9291/next-generation-sequencing-v7.pdf (accessed April 11, 2020).

50 Ainsworth BE, Haskell WL, Herrmann SD, et al. 2011 Compendium of Physical Activities: a second update of codes and MET values. Med Sci Sports Exerc 2011; 43 : 1575–81.

51 Walker VM, Davies NM, Hemani G, et al. Using the MR-Base platform to investigate risk factors and drug targets for thousands of phenotypes. Wellcome Open Res 2019; 4 : 113.

52 Myers TA, Chanock SJ, Machiela MJ. LDlinkR: An R Package for Rapidly Calculating Linkage Disequilibrium Statistics in Diverse Populations. Front Genet 2020; 11 : 157.

