## Supplementary Figures 1-5, Supplementary Tables 1-9, and Supplementary Note for "Physical exercise is a risk factor for amyotrophic lateral sclerosis: Convergent evidence from mendelian randomisation, transcriptomics and risk genotypes"

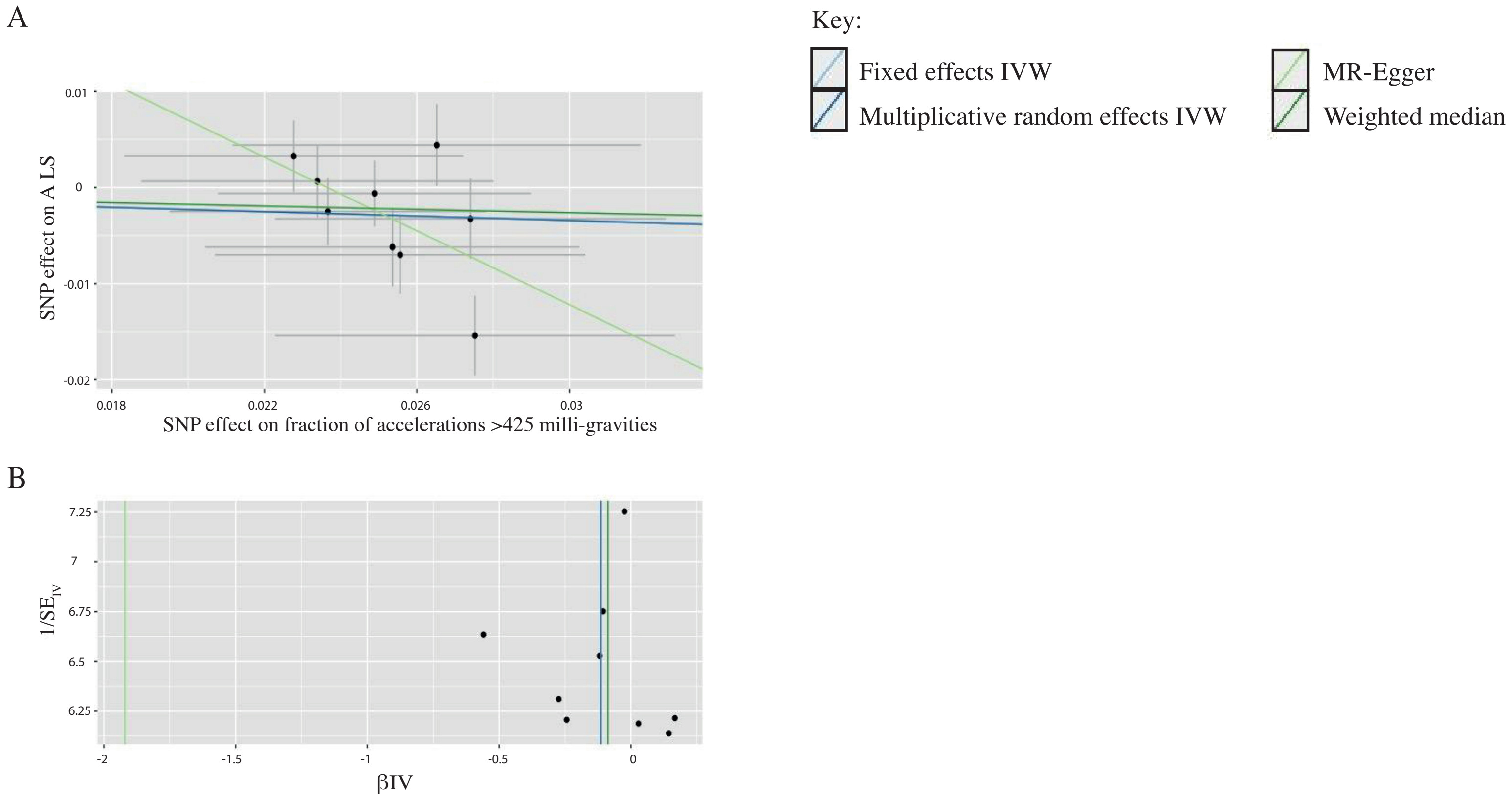
A
B
Key:
Fixed effects IVW
Multiplicative random effects IVW
MR-Egger
Weighted median
SNP effect on A LS
SNP effect on fraction of accelerations >425 milli-gravities
0.01
0
-0.01
-0.02
0.018
0.022
0.026
0.03
1/SE$_{IV}$
βIV
7.25
7
6.75
6.5
6.25
-2
-1.5
-1
-0.5
0

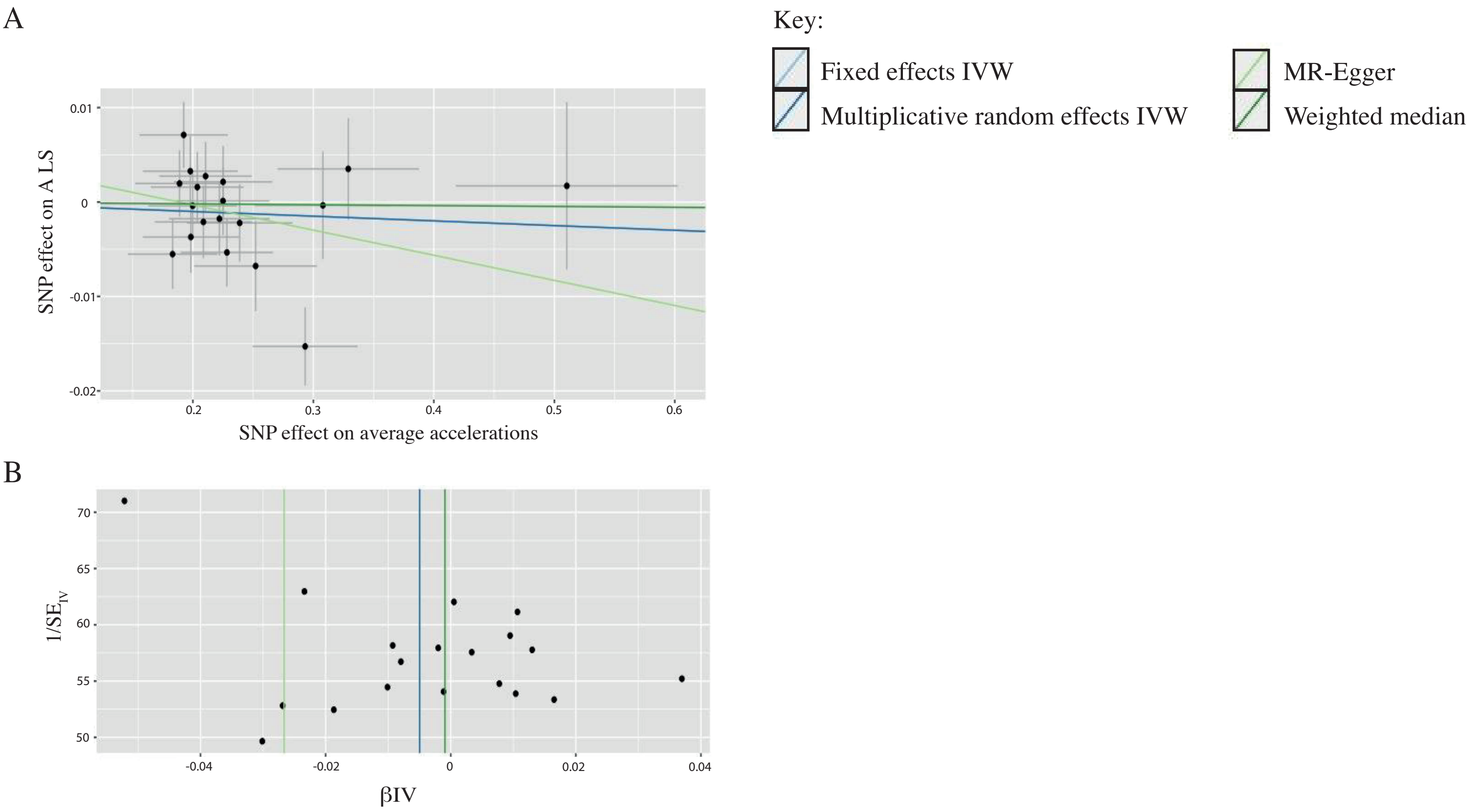
A
B
Key:
Fixed effects IVW
Multiplicative random effects IVW
MR-Egger
Weighted median
SNP effect on A LS
SNP effect on average accelerations
0.01
0
-0.01
-0.02
0.2
0.3
0.4
0.5
0.6
1/SE
IV
βIV
70
65
60
55
50
-0.04
-0.02
0
0.02
0.04

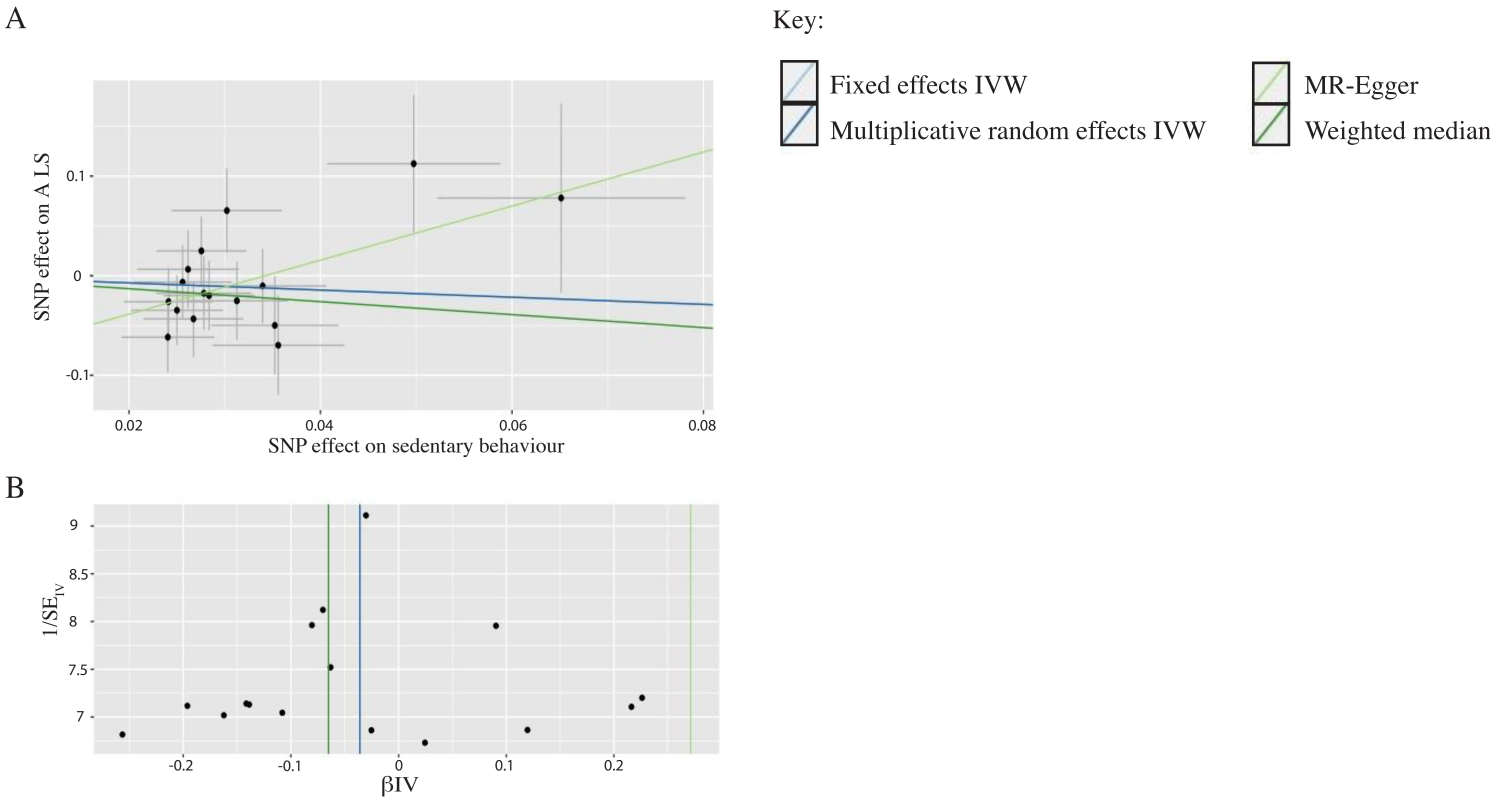
A
Key:
Fixed effects IVW
Multiplicative random effects IVW
MR-Egger
Weighted median
SNP effect on A LS
0.1
0
-0.1
0.02
0.04
0.06
0.08
SNP effect on sedentary behaviour
B
$1/SE_{IV}$
9
8.5
8
7.5
7
-0.2
-0.1
0
0.1
0.2
βIV

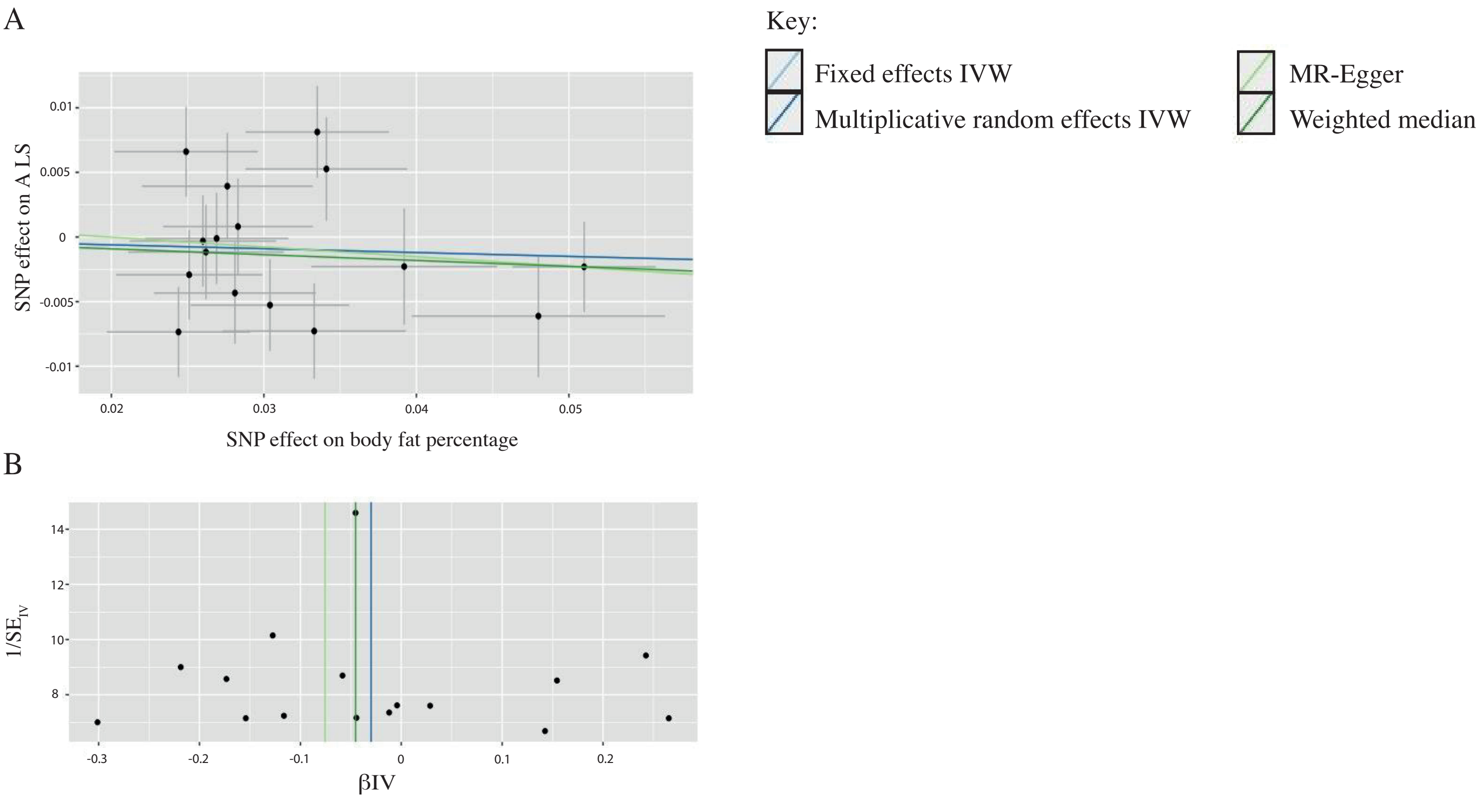
A
Key:
Fixed effects IVW
Multiplicative random effects IVW
MR-Egger
Weighted median
SNP effect on A LS
SNP effect on body fat percentage
0.01
0.005
0
-0.005
-0.01
0.02
0.03
0.04
0.05
B
1/SE_IV
βIV
14
12
10
8
-0.3
-0.2
-0.1
0
0.1
0.2

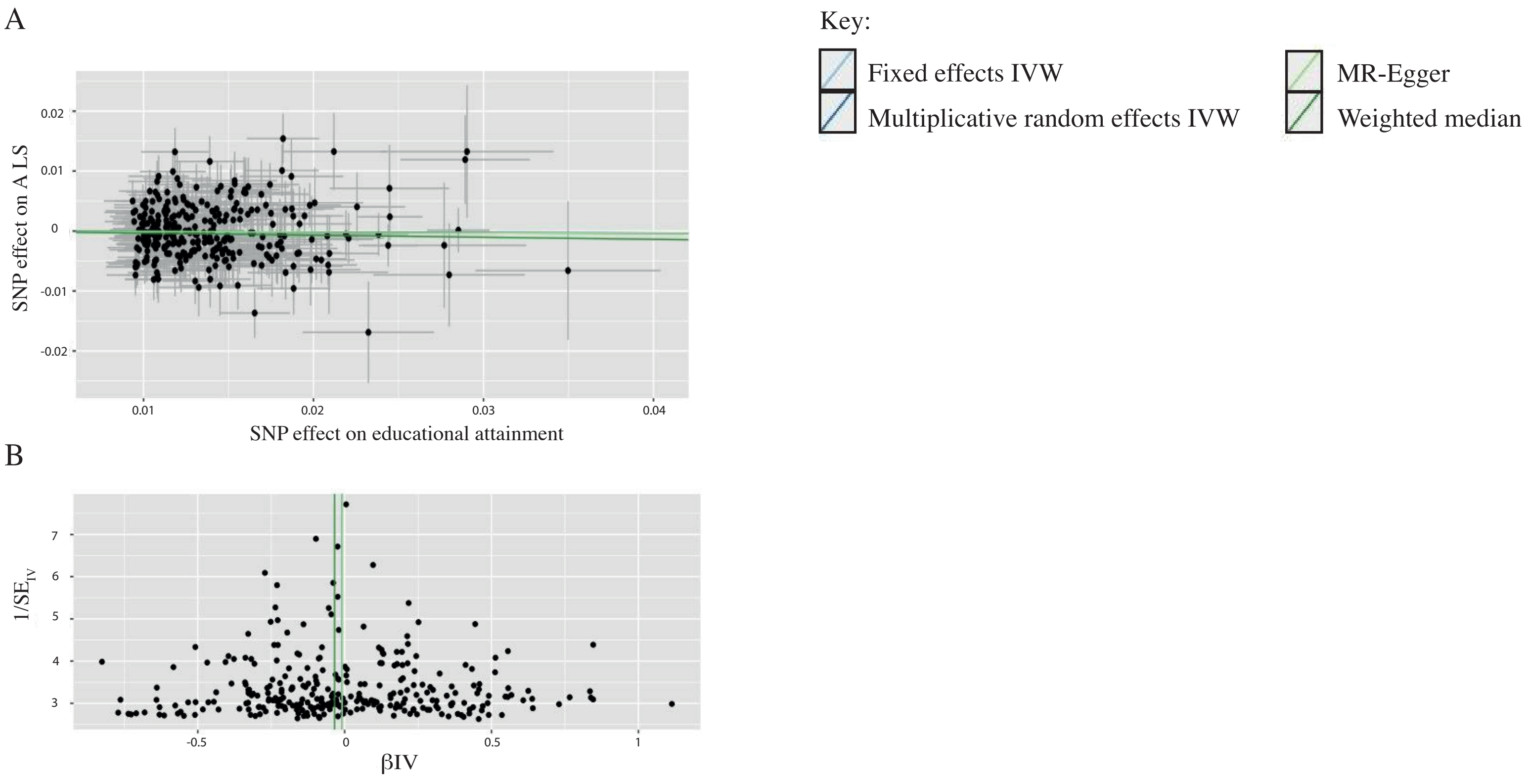
A
B
Key:
Fixed effects IVW
Multiplicative random effects IVW
MR-Egger
Weighted median
SNP effect on A LS
SNP effect on educational attainment
0.02
0.01
0
-0.01
-0.02
0.01
0.02
0.03
0.04
$1/SE_{IV}$
βIV
7
6
5
4
3
-0.5
0
0.5
1

#### Supplementary Table 1: Supplementary content relating to the MR analysis of the SSOE - ALS relationship.

**Key**: SNP = single nucleotide polymorphism, PVE = proportion of variance explained, SE = standard error, EAF = effect allele frequency , ALS = amyotrophic lateral sclerosis , SSOE = strenuous sport or other exercise, VPA = vigorous physical activity, MVPA = moderate-vigorous physical activity, EA = educational attainment, BF% = body fat percentage, Chrom = chromosome, IVW = Inverse variance weighted, df = degrees of freedom, fe = fixed effects, mre = multiplicative random effects

**Conservative instrument, SNP-SSOE p<5E-08**

**Table 1A: SNPs identified after clumping and inclusion of proxy variants**

| Index | Chrom | SNP | Proxy SNP | Effect Allele | SSOE beta | ALS beta | SSOE EAF | ALS EAF | ALS SE | ALS p value | SSOE SE | SSOE p value | Pallindromic | PVE | F statistic |
|---|---|---|---|---|---|---|---|---|---|---|---|---|---|---|---|
| 1 | 6 | rs10946808 | N/A | A | -0.008 | -0.005 | 0.73 | 0.71 | 0.004 | 0.15 | 0.001 | 9.90E-10 | No | 1.07E-04 | 37.34 |
| 2 | 1 | rs1200154 | N/A | G | -0.006 | -0.005 | 0.41 | 0.42 | 0.003 | 0.14 | 0.001 | 3.90E-08 | No | 8.62E-05 | 30.21 |
| 3 | 5 | rs159544 | N/A | A | -0.007 | 0.002 | 0.61 | 0.60 | 0.004 | 0.61 | 0.001 | 1.30E-09 | No | 1.05E-04 | 36.76 |
| 4 | 17 | rs166840 | N/A | G | 0.008 | 0.004 | 0.59 | 0.60 | 0.004 | 0.28 | 0.001 | 3.10E-11 | No | 1.26E-04 | 44.13 |
| 5 | 2 | rs288070 | N/A | G | -0.01 | 0.001 | 0.90 | 0.89 | 0.006 | 0.82 | 0.002 | 1.90E-08 | No | 9.01E-05 | 31.57 |
| 6 | 1 | rs2994326 | N/A | T | -0.008 | 0.005 | 0.19 | 0.18 | 0.005 | 0.31 | 0.001 | 4.50E-08 | No | 8.54E-05 | 29.93 |
| 7 | 13 | rs4411372 | N/A | T | -0.007 | -0.006 | 0.72 | 0.72 | 0.004 | 0.11 | 0.001 | 2.00E-08 | No | 8.97E-05 | 31.46 |
| 8 | 5 | rs4865667 | N/A | C | 0.007 | 0.008 | 0.61 | 0.61 | 0.004 | 0.02 | 0.001 | 1.00E-08 | No | 9.35E-05 | 32.77 |
| 9 | 3 | rs62253088 | N/A | T | 0.01 | 0.0003 | 0.33 | 0.33 | 0.004 | 0.95 | 0.001 | 1.00E-19 | No | 2.36E-04 | 82.57 |
| 10 | 3 | rs7627864 | N/A | C | 0.007 | -0.002 | 0.57 | 0.58 | 0.003 | 0.61 | 0.001 | 7.60E-09 | Yes | 9.52E-05 | 33.37 |
| 11 | 6 | rs896302 | N/A | C | 0.007 | -0.001 | 0.29 | 0.29 | 0.004 | 0.74 | 0.001 | 1.70E-08 | No | 9.06E-05 | 31.76 |

**Table 1B: Cochran's Q test**

| Method | Q | df | p value |
|---|---|---|---|
| MR Egger | 8.92 | 8 | 0.35 |
| IVW | 11.62 | 9 | 0.24 |

**Table 1C: MR-Egger intercept test**

| Egger intercept | SE | p value |
|---|---|---|
| 0.01 | 0.007 | 0.16 |

**Table 1D: I^2 statistic**

| | |
|---|---|
| **I^2** | 0.97 |

**Table 1E: F statistic for the whole instrument**

| F statistic | 38.9 |
|---|---|

**Table 1F: MR-PRESSO global test**

| Global test | p value | Outlier index | Outlier test | Distortion test p value |
|---|---|---|---|---|
| 14.08 | 0.26 | N/A | N/A | N/A |

**Table 1G: Causal estimates**

| Method | SNPs (n) | Beta | SE | p value |
|---|---|---|---|---|
| IVW (fe) | 10 | 0.28 | 0.16 | 0.07 |
| IVW (mre) | 10 | 0.28 | 0.18 | 0.11 |
| MR Egger | 10 | -1.09 | 0.89 | 0.26 |
| Weighted median | 10 | 0.17 | 0.22 | 0.44 |

**Table 1H: Leave one out analysis with fixed effects IVW**

| SNP removed | Beta | SE | p value |
|---|---|---|---|
| rs10946808 | 0.23 | 0.16 | 0.16 |
| rs1200154 | 0.23 | 0.16 | 0.15 |
| rs159544 | 0.34 | 0.16 | 0.04 |
| rs166840 | 0.25 | 0.16 | 0.13 |
| rs288070 | 0.32 | 0.16 | 0.05 |
| rs2994326 | 0.35 | 0.16 | 0.03 |
| rs4411372 | 0.23 | 0.16 | 0.16 |
| rs4865667 | 0.19 | 0.16 | 0.24 |
| rs62253088 | 0.35 | 0.17 | 0.05 |
| rs896302 | 0.32 | 0.16 | 0.05 |

**Liberal instrument, SNP-SSOE p<1E-06**

**Table 1I: SNPs identified after clumping and inclusion of proxy variants**

| Index | Chrom | SNP | Proxy SNP | Effect Allele | SSOE beta | ALS beta | SSOE EAF | ALS EAF | ALS SE | ALS p value | SSOE SE | SSOE p value | Pallindromic | PVE | F statistic |
|---|---|---|---|---|---|---|---|---|---|---|---|---|---|---|---|
| **1** | 6 | rs10946808 | N/A | A | -0.008 | -0.005 | 0.73 | 0.71 | 0.004 | 0.15 | 0.001 | 9.90E-10 | No | 1.07E-04 | 37.34 |
| **2** | 9 | rs11143585 | N/A | C | 0.007 | 0.003 | 0.75 | 0.76 | 0.004 | 0.48 | 0.001 | 2.50E-07 | No | 7.58E-05 | 26.57 |
| **3** | 1 | rs11210230 | N/A | T | -0.009 | -0.004 | 0.88 | 0.89 | 0.005 | 0.41 | 0.002 | 1.30E-07 | No | 7.96E-05 | 27.89 |
| **4** | 10 | rs113428529 | N/A | A | 0.009 | 0.001 | 0.86 | 0.88 | 0.006 | 0.90 | 0.002 | 2.40E-07 | No | 7.60E-05 | 26.65 |
| **5** | 14 | rs11625806 | N/A | G | -0.006 | -0.003 | 0.59 | 0.59 | 0.004 | 0.40 | 0.001 | 6.80E-07 | No | 7.04E-05 | 24.68 |
| **6** | 14 | rs11628052 | N/A | A | -0.006 | -0.003 | 0.54 | 0.54 | 0.003 | 0.47 | 0.001 | 5.90E-07 | No | 7.12E-05 | 24.95 |
| **7** | 2 | rs11687833 | N/A | C | 0.006 | -0.003 | 0.60 | 0.59 | 0.004 | 0.42 | 0.001 | 6.30E-07 | No | 7.08E-05 | 24.82 |
| **8** | 20 | rs118165004 | N/A | C | 0.007 | 0.007 | 0.82 | 0.83 | 0.005 | 0.15 | 0.001 | 8.60E-07 | No | 6.91E-05 | 24.21 |
| **9** | 1 | rs1200154 | N/A | G | -0.006 | -0.005 | 0.41 | 0.42 | 0.003 | 0.14 | 0.001 | 3.90E-08 | No | 8.62E-05 | 30.21 |
| **10** | 12 | rs12821008 | rs12298884 | G | -0.006 | -0.004 | 0.59 | 0.60 | 0.004 | 0.31 | 0.001 | 1.70E-07 | No | 7.79E-05 | 27.30 |
| **11** | 5 | rs13180692 | N/A | G | -0.006 | -0.003 | 0.56 | 0.56 | 0.003 | 0.47 | 0.001 | 1.70E-07 | No | 7.81E-05 | 27.39 |
| **12** | 4 | rs144949097 | N/A | G | -0.020 | -0.017 | 0.98 | 0.97 | 0.011 | 0.13 | 0.004 | 2.30E-07 | No | 7.64E-05 | 26.78 |
| **13** | 5 | rs159544 | N/A | A | -0.007 | 0.002 | 0.61 | 0.60 | 0.004 | 0.61 | 0.001 | 1.30E-09 | No | 1.05E-04 | 36.76 |
| **14** | 17 | rs166840 | N/A | G | 0.008 | 0.004 | 0.59 | 0.60 | 0.004 | 0.28 | 0.001 | 3.10E-11 | No | 1.26E-04 | 44.13 |
| **15** | 18 | rs1788826 | N/A | G | -0.006 | -0.003 | 0.35 | 0.36 | 0.004 | 0.44 | 0.001 | 7.60E-07 | No | 6.97E-05 | 24.45 |
| **16** | 10 | rs2148201 | N/A | G | -0.006 | 0.002 | 0.51 | 0.51 | 0.003 | 0.65 | 0.001 | 4.10E-07 | No | 7.32E-05 | 25.64 |
| **17** | 17 | rs216211 | N/A | A | 0.007 | 0.007 | 0.25 | 0.26 | 0.004 | 0.08 | 0.001 | 4.60E-07 | No | 7.25E-05 | 25.41 |
| **18** | 12 | rs2238046 | N/A | A | -0.006 | 0.004 | 0.61 | 0.63 | 0.004 | 0.21 | 0.001 | 7.40E-07 | No | 6.99E-05 | 24.51 |
| **19** | 7 | rs246743 | N/A | T | 0.008 | 0.003 | 0.15 | 0.15 | 0.005 | 0.58 | 0.002 | 5.50E-07 | No | 7.15E-05 | 25.07 |
| **20** | 8 | rs2725370 | N/A | T | -0.007 | -0.004 | 0.30 | 0.30 | 0.004 | 0.32 | 0.001 | 1.10E-07 | No | 8.04E-05 | 28.19 |
| **21** | 4 | rs2749778 | N/A | T | 0.006 | -0.001 | 0.40 | 0.40 | 0.004 | 0.83 | 0.001 | 1.00E-07 | No | 8.08E-05 | 28.33 |
| **22** | 16 | rs28551054 | N/A | A | -0.007 | -0.008 | 0.76 | 0.75 | 0.004 | 0.05 | 0.001 | 9.00E-07 | No | 6.89E-05 | 24.14 |
| **23** | 2 | rs2871340 | N/A | G | -0.006 | 0.006 | 0.50 | 0.51 | 0.003 | 0.10 | 0.001 | 1.10E-07 | No | 8.04E-05 | 28.18 |
| **24** | 2 | rs288070 | N/A | G | -0.01 | 0.001 | 0.90 | 0.89 | 0.006 | 0.82 | 0.002 | 1.90E-08 | No | 9.01E-05 | 31.57 |
| **25** | 1 | rs2994326 | N/A | T | -0.008 | 0.005 | 0.19 | 0.18 | 0.005 | 0.31 | 0.001 | 4.50E-08 | No | 8.54E-05 | 29.93 |
| **26** | 3 | rs336620 | N/A | C | 0.006 | 0.001 | 0.30 | 0.31 | 0.004 | 0.69 | 0.001 | 2.00E-07 | No | 7.72E-05 | 27.05 |
| **27** | 4 | rs35518360 | N/A | A | 0.01 | -0.004 | 0.92 | 0.92 | 0.007 | 0.52 | 0.002 | 1.10E-07 | No | 8.06E-05 | 28.26 |
| **28** | 5 | rs35786746 | N/A | C | 0.007 | 0.006 | 0.81 | 0.83 | 0.005 | 0.20 | 0.001 | 6.10E-07 | No | 7.10E-05 | 24.88 |
| **29** | 2 | rs4084828 | N/A | C | -0.006 | 0.006 | 0.46 | 0.47 | 0.004 | 0.08 | 0.001 | 4.50E-07 | No | 7.26E-05 | 25.45 |
| **30** | 13 | rs4411372 | N/A | T | -0.007 | -0.006 | 0.72 | 0.72 | 0.004 | 0.11 | 0.001 | 2.00E-08 | No | 8.97E-05 | 31.46 |

| | | | | | | | | | | | | | | | |
|---|---|---|---|---|---|---|---|---|---|---|---|---|---|---|---|
| **31** | 20 | rs4811141 | N/A | G | -0.008 | -0.004 | 0.13 | 0.13 | 0.005 | 0.41 | 0.002 | 6.50E-07 | No | 7.06E-05 | 24.75 |
| **32** | 5 | rs4865667 | N/A | C | 0.007 | 0.008 | 0.61 | 0.61 | 0.004 | 0.02 | 0.001 | 1.00E-08 | No | 9.35E-05 | 32.77 |
| **33** | 11 | rs547118 | N/A | C | 0.006 | -0.008 | 0.44 | 0.43 | 0.003 | 0.02 | 0.001 | 6.40E-07 | Yes | 7.07E-05 | 24.77 |
| **34** | 11 | rs551243 | N/A | G | 0.006 | -0.006 | 0.53 | 0.52 | 0.003 | 0.10 | 0.001 | 6.50E-08 | Yes | 8.34E-05 | 29.22 |
| **35** | 10 | rs577525 | N/A | T | 0.006 | -0.005 | 0.44 | 0.44 | 0.003 | 0.14 | 0.001 | 2.70E-07 | No | 7.54E-05 | 26.43 |
| **36** | 19 | rs62109945 | N/A | T | -0.006 | 0.001 | 0.50 | 0.51 | 0.004 | 0.71 | 0.001 | 3.30E-07 | No | 7.43E-05 | 26.05 |
| **37** | 3 | rs62253088 | N/A | T | 0.01 | 0.000 | 0.33 | 0.33 | 0.004 | 0.95 | 0.001 | 1.00E-19 | No | 2.36E-04 | 82.57 |
| **38** | 6 | rs62422094 | N/A | A | -0.007 | -0.001 | 0.76 | 0.77 | 0.004 | 0.75 | 0.001 | 2.90E-07 | No | 7.51E-05 | 26.32 |
| **39** | 7 | rs62472745 | N/A | C | -0.007 | 0.006 | 0.79 | 0.79 | 0.004 | 0.13 | 0.001 | 4.90E-07 | No | 7.22E-05 | 25.32 |
| **40** | 2 | rs6432141 | N/A | A | 0.006 | 0.004 | 0.45 | 0.44 | 0.003 | 0.20 | 0.001 | 6.90E-08 | No | 8.30E-05 | 29.11 |
| **41** | 9 | rs7031064 | N/A | A | -0.006 | 0.004 | 0.52 | 0.53 | 0.003 | 0.29 | 0.001 | 3.80E-07 | No | 7.36E-05 | 25.80 |
| **42** | 9 | rs7034243 | N/A | A | -0.006 | 0.001 | 0.66 | 0.66 | 0.004 | 0.70 | 0.001 | 1.00E-07 | No | 8.08E-05 | 28.34 |
| **43** | 1 | rs705692 | N/A | C | -0.009 | -0.013 | 0.11 | 0.11 | 0.005 | 0.02 | 0.002 | 6.30E-07 | No | 7.08E-05 | 24.81 |
| **44** | 6 | rs72910629 | N/A | A | 0.009 | 0.007 | 0.86 | 0.87 | 0.005 | 0.17 | 0.002 | 1.40E-07 | No | 7.92E-05 | 27.76 |
| **45** | 8 | rs75276987 | N/A | T | -0.010 | -0.004 | 0.92 | 0.92 | 0.006 | 0.52 | 0.002 | 8.80E-07 | No | 6.90E-05 | 24.18 |
| **46** | 3 | rs7627864 | N/A | C | 0.007 | -0.002 | 0.57 | 0.58 | 0.003 | 0.61 | 0.001 | 7.60E-09 | Yes | 9.52E-05 | 33.37 |
| **47** | 6 | rs763187 | N/A | A | -0.008 | 0.000 | 0.17 | 0.17 | 0.005 | 0.98 | 0.002 | 3.50E-07 | No | 7.40E-05 | 25.93 |
| **48** | 6 | rs7740107 | N/A | T | -0.006 | -0.001 | 0.26 | 0.26 | 0.004 | 0.78 | 0.001 | 6.10E-07 | No | 7.10E-05 | 24.88 |
| **49** | 4 | rs795973 | N/A | A | -0.006 | 0.003 | 0.67 | 0.67 | 0.004 | 0.45 | 0.001 | 9.30E-07 | No | 6.87E-05 | 24.08 |
| **50** | 18 | rs8093216 | N/A | G | -0.006 | 0.000 | 0.65 | 0.66 | 0.004 | 0.90 | 0.001 | 9.80E-07 | No | 6.84E-05 | 23.97 |
| **51** | 19 | rs111901094 | rs8182472 | T | 0.006 | 0.004 | 0.88 | 0.89 | 0.005 | 0.51 | 0.002 | 7.80E-04 | No | 3.22E-05 | 11.29 |
| **52** | 6 | rs896302 | N/A | C | 0.007 | -0.001 | 0.29 | 0.29 | 0.004 | 0.74 | 0.001 | 1.70E-08 | No | 9.06E-05 | 31.76 |
| **53** | 7 | rs9177 | N/A | A | 0.006 | 0.006 | 0.51 | 0.52 | 0.004 | 0.12 | 0.001 | 4.60E-07 | No | 7.26E-05 | 25.43 |
| **54** | 22 | rs9604803 | N/A | C | 0.006 | -0.001 | 0.67 | 0.66 | 0.004 | 0.81 | 0.001 | 8.00E-07 | No | 6.95E-05 | 24.36 |
| **55** | 3 | rs9881048 | N/A | A | -0.007 | -0.0003 | 0.19 | 0.20 | 0.004 | 0.94 | 0.001 | 7.70E-07 | No | 6.97E-05 | 24.44 |
| **56** | 17 | rs9903845 | N/A | C | 0.006 | 0.005 | 0.67 | 0.68 | 0.004 | 0.20 | 0.001 | 7.30E-07 | No | 7.00E-05 | 24.53 |

**Table 1J: Cochran's Q test**

| **Method** | **Q** | **df** | **p value** |
|---|---|---|---|
| MR Egger | 54.49 | 51.00 | 0.34 |
| IVW | 56.43 | 52.00 | 0.31 |

**Table 1K: Mr-Egger intercept test**

| Egger intercept | SE | p value |
|---|---|---|
| -0.004 | 0.003 | 0.18 |

**Table 1L: I^2 statistic**

| | |
|---|---|
| I^2 | 0.96 |

**Table 1M: F statistic for the whole instrument**

| | |
|---|---|
| F statistic | 28.15 |

**Table 1N: MR-PRESSO global test**

| Global test | p value | Outlier index | Outlier test | Distortion test p value |
|---|---|---|---|---|
| 58.59 | 0.33 | N/A | N/A | N/A |

**Table 1O: Causal estimates**

| Method | SNPs (n) | Beta | SE | p value |
|---|---|---|---|---|
| IVE (fe) | 53 | 0.21355 | 0.07959 | 0.00729 |
| IVW (mre) | 53 | 0.21355 | 0.08290 | 0.01000 |
| MR Egger | 53 | 0.75186 | 0.40859 | 0.07157 |
| Weighted median | 53 | 0.21645 | 0.11163 | 0.05250 |
| Weighted mode | 53 | 0.51895 | 0.24563 | 0.03944 |

**Table 1P: Leave one out analysis with fixed effects IVW**

| SNP removed | Beta | SE | p value |
|---|---|---|---|
| rs10946808 | 0.20 | 0.08 | 0.01 |
| rs11143585 | 0.21 | 0.08 | 0.009 |
| rs11210230 | 0.21 | 0.08 | 0.009 |
| rs113428529 | 0.22 | 0.08 | 0.007 |
| rs11625806 | 0.21 | 0.08 | 0.009 |
| rs11628052 | 0.21 | 0.08 | 0.009 |

| | | | |
|---|---|---|---|
| rs11687833 | 0.23 | 0.08 | 0.005 |
| rs118165004 | 0.20 | 0.08 | 0.01 |
| rs1200154 | 0.20 | 0.08 | 0.01 |
| rs12298884 | 0.21 | 0.08 | 0.01 |
| rs13180692 | 0.21 | 0.08 | 0.009 |
| rs144949097 | 0.20 | 0.08 | 0.01 |
| rs159544 | 0.23 | 0.08 | 0.005 |
| rs166840 | 0.20 | 0.08 | 0.01 |
| rs1788826 | 0.21 | 0.08 | 0.009 |
| rs2148201 | 0.22 | 0.08 | 0.006 |
| rs216211 | 0.20 | 0.08 | 0.013 |
| rs2238046 | 0.23 | 0.08 | 0.004 |
| rs246743 | 0.21 | 0.08 | 0.008 |
| rs2725370 | 0.21 | 0.08 | 0.010 |
| rs2749778 | 0.22 | 0.08 | 0.006 |
| rs28551054 | 0.20 | 0.08 | 0.01 |
| rs2871340 | 0.24 | 0.08 | 0.003 |
| rs288070 | 0.22 | 0.08 | 0.006 |
| rs2994326 | 0.23 | 0.08 | 0.004 |
| rs336620 | 0.21 | 0.08 | 0.008 |
| rs35518360 | 0.22 | 0.08 | 0.005 |
| rs35786746 | 0.20 | 0.08 | 0.01 |
| rs4084828 | 0.24 | 0.08 | 0.003 |
| rs4411372 | 0.20 | 0.08 | 0.013 |
| rs4811141 | 0.21 | 0.08 | 0.009 |
| rs4865667 | 0.19 | 0.08 | 0.02 |
| rs577525 | 0.23 | 0.08 | 0.004 |
| rs62109945 | 0.22 | 0.08 | 0.006 |
| rs62253088 | 0.22 | 0.08 | 0.006 |
| rs62422094 | 0.21 | 0.08 | 0.008 |
| rs62472745 | 0.23 | 0.08 | 0.004 |
| rs6432141 | 0.20 | 0.08 | 0.01 |

| | | | |
|---|---|---|---|
| rs7031064 | 0.23 | 0.08 | 0.004 |
| rs7034243 | 0.22 | 0.08 | 0.006 |
| rs705692 | 0.19 | 0.08 | 0.02 |
| rs72910629 | 0.20 | 0.08 | 0.01 |
| rs75276987 | 0.21 | 0.08 | 0.009 |
| rs763187 | 0.22 | 0.08 | 0.007 |
| rs7740107 | 0.21 | 0.08 | 0.008 |
| rs795973 | 0.22 | 0.08 | 0.005 |
| rs8093216 | 0.22 | 0.08 | 0.006 |
| rs8182472 | 0.21 | 0.08 | 0.008 |
| rs896302 | 0.22 | 0.08 | 0.006 |
| rs9177 | 0.20 | 0.08 | 0.012 |
| rs9604803 | 0.22 | 0.08 | 0.006 |
| rs9881048 | 0.22 | 0.08 | 0.007 |
| rs9903845 | 0.20 | 0.08 | 0.01 |

**Supplementary Table 2: Supplementary content relating to the MR analysis of the accerations >425 milligravities - ALS relationship.**

**Key**: SNP = single nucleotide polymorphism, PVE = proportion of variance explained, SE = standard error, EAF = effect allele frequency , ALS = amyotrophic lateral sclerosis , SSOE = strenuous sport or other exercise, VPA = vigorous physical activity, MVPA = moderate-vigorous physical activity, EA = educational attainment, BF% = body fat percentage, Chrom = chromosome, IVW = Inverse variance weighted, df = degrees of freedom, fe = fixed effects, mre = multiplicative random effects

**Conservative instrument, SNP-accelerations >425 milligravities p<5E-08**

**Table 2A: SNPs identified after clumping and inclusion of proxy variants**

| Index | Chrom | SNP | Proxy SNP | Effect Allele | Accelerations beta | ALS beta | Accelerations EAF | ALS EAF | ALS SE | ALS p value | Accelerations SE | Accelerations p value | Pallindromic | PVE | F statistic |
|---|---|---|---|---|---|---|---|---|---|---|---|---|---|---|---|
| 1 | 2 | rs6433478 | N/A | T | -0.02 | 0.003 | 0.46 | 0.47 | 0.004 | 0.48 | 0.004 | 1.20E-08 | No | 0.0004 | 32.47 |
| 2 | 15 | rs743580 | N/A | A | 0.02 | -0.001 | 0.51 | 0.50 | 0.003 | 0.86 | 0.004 | 1.30E-09 | No | 0.0004 | 36.76 |

**Table 2B: Cochran's Q test**

| Method | Q | df | p value |
|---|---|---|---|
| IVW | 0.16 | 1 | 0.69 |

**Due to the small number of SNPs a MR-Egger intercept test and MR-PRESSO cannot be performed. I^2 is not relevant given that an MR-Egger cannot be performed.**

**Table 2C: F statistic for the whole instrument**

| | |
|---|---|
| **F statistic** | 36.29 |

**Table 2D:Causal estimates**

| Method | SNPs (n) | Beta | SE | p value |
|---|---|---|---|---|
| IVW (fe) | 2 | -0.06 | 0.1 | 0.54 |
| IVW (mre) | 2 | -0.06 | 0.04 | 0.12 |

**Liberal instrument, SNP-accelerations > 425 milligravities p<1E-06**

**Table 2E: SNPs identified after clumping and inclusion of proxy variants**

| Index | Chrom | SNP | Proxy SNP | Effect Allele | Accelerations beta | ALS beta | Accelerations EAF | ALS EAF | ALS SE | ALS p value | Accelerations SE | Accelerations p value | Pallindromic | PVE | F statistic |
|---|---|---|---|---|---|---|---|---|---|---|---|---|---|---|---|
| 1 | 16 | rs11866559 | N/A | G | -0.03 | -0.004 | 0.82 | 0.80 | 0.004 | 0.30 | 0.005 | 7.20E-07 | No | 0.0003 | 24.56 |

| | | | | | | | | | | | | | | | |
|---|---|---|---|---|---|---|---|---|---|---|---|---|---|---|---|
| **2** | 18 | rs1668835 | N/A | T | -0.02 | -0.003 | 0.69 | 0.69 | 0.004 | 0.38 | 0.004 | 3.10E-07 | No | 0.0003 | 26.18 |
| **3** | 1 | rs1856329 | N/A | A | 0.03 | -0.003 | 0.80 | 0.79 | 0.004 | 0.44 | 0.005 | 9.00E-08 | No | 0.0003 | 28.57 |
| **4** | 11 | rs4754194 | N/A | C | -0.03 | 0.006 | 0.77 | 0.77 | 0.004 | 0.13 | 0.005 | 2.40E-07 | No | 0.0003 | 26.64 |
| **5** | 7 | rs62443625 | N/A | T | -0.03 | 0.007 | 0.77 | 0.76 | 0.004 | 0.08 | 0.005 | 1.40E-07 | No | 0.0003 | 27.68 |
| **6** | 2 | rs6433478 | N/A | T | -0.02 | 0.003 | 0.46 | 0.47 | 0.004 | 0.48 | 0.004 | 1.20E-08 | No | 0.0004 | 32.47 |
| **7** | 8 | rs72633364 | N/A | G | -0.02 | -0.001 | 0.71 | 0.70 | 0.004 | 0.86 | 0.005 | 4.10E-07 | No | 0.0003 | 25.63 |
| **8** | 15 | rs743580 | N/A | A | 0.02 | -0.001 | 0.51 | 0.50 | 0.003 | 0.86 | 0.004 | 1.30E-09 | No | 0.0004 | 36.76 |
| **9** | 17 | rs80028338 | N/A | A | -0.03 | 0.02 | 0.79 | 0.78 | 0.004 | 0.0002 | 0.005 | 1.50E-07 | No | 0.0003 | 27.55 |

**Table 2F: Cochran's Q test**

| Method | Q | df | p value |
|---|---|---|---|
| MR Egger | 12.13 | 7 | 0.10 |
| IVW | 17.18 | 8 | 0.03 |

**Table 2G: MR-Egger intercept test**

| Egger intercept | SE | p value |
|---|---|---|
| 0.05 | 0.03 | 0.13 |

**Table 2H: I^2 statistic**

| | |
|---|---|
| **I^2** | 0.96 |

**Table 2I: F statistic for the whole instrument**

| | |
|---|---|
| **F statistic** | 28.28 |

**Table 2J: MR-PRESSO global test**

| Global test | p value | Outlier index | Outlier test | Distortion test p value |
|---|---|---|---|---|
| 21.7 | 0.03 | 9 | p=0.009 for index 9 | 0.08 |

**Table 2K: Causal estimates**

| Method | SNPs (n) | Beta | SE | p value |
|---|---|---|---|---|
| IVW (fe) | 9 | -0.11 | 0.05 | 0.03 |

| | | | | |
|---|---|---|---|---|
| IVW (mre) | 9 | -0.11 | 0.08 | 0.13 |
| MR Egger | 9 | -1.92 | 1.06 | 0.11 |
| Weighted median | 9 | -0.09 | 0.07 | 0.24 |

**Table 2L: Leave one out with multiplicative random effects IVW**

| SNP removed | Beta | SE | p value |
|---|---|---|---|
| rs11866559 | -0.15 | 0.08 | 0.05 |
| rs1668835 | -0.14 | 0.08 | 0.07 |
| rs1856329 | -0.11 | 0.09 | 0.18 |
| rs4754194 | -0.10 | 0.08 | 0.23 |
| rs62443625 | -0.10 | 0.08 | 0.25 |
| rs6433478 | -0.12 | 0.09 | 0.18 |
| rs72633364 | -0.13 | 0.08 | 0.12 |
| rs743580 | -0.13 | 0.09 | 0.13 |
| rs80028338* | -0.06 | 0.06 | 0.32 |

* SNP identified to be an outlier during MR-PRESSO. Removal of this SNP reduces significance of the exposure-outcome relationship.

**Supplementary Table 3: Supplementary content relating to the MR analysis of the average accelerations - ALS relationship.**

**Key**: SNP = single nucleotide polymorphism, PVE = proportion of variance explained, SE = standard error, EAF = effect allele frequency , ALS = amyotrophic lateral sclerosis , SSOE = strenuous sport or other exercise, VPA = vigorous physical activity, MVPA = moderate-vigorous physical activity, EA = educational attainment, BF% = body fat percentage, Chrom = chromosome, IVW = Inverse variance weighted, df = degrees of freedom, fe = fixed effects, mre = multiplicative random effects

**Conservative instrument, SNP-average accelerations p<5E-08**

**Table 3A: SNPs identified after clumping and inclusion of proxy variants**

| Index | Chrom | SNP | Proxy SNP | Effect Allele | Accelerations beta | ALS beta | Accelerations EAF | ALS EAF | ALS SE | ALS p value | Accelerations SE | Accelerations p value | Pallindromic | PVE | F statistic |
|---|---|---|---|---|---|---|---|---|---|---|---|---|---|---|---|
| **1** | 10 | rs11012732 | N/A | A | 0.22 | 0.0001 | 0.67 | 0.65 | 0.004 | 0.97 | 0.04 | 5.40E-09 | No | 0.0004 | 34.04 |
| **2** | 5 | rs12522261 | N/A | G | 0.21 | 0.003 | 0.66 | 0.66 | 0.004 | 0.45 | 0.04 | 3.90E-08 | No | 0.0003 | 30.21 |
| **3** | 11 | rs148193266 | N/A | A | -0.51 | -0.002 | 0.96 | 0.96 | 0.009 | 0.85 | 0.09 | 3.10E-08 | No | 0.0003 | 30.67 |
| **4** | 1 | rs34517439 | N/A | C | 0.31 | -0.0004 | 0.88 | 0.89 | 0.006 | 0.95 | 0.06 | 4.40E-08 | No | 0.0003 | 29.97 |
| **5** | 17 | rs56194509 | rs55725840 | T | -0.29 | 0.02 | 0.78 | 0.78 | 0.004 | 0.0002 | 0.04 | 1.70E-11 | No | 0.0005 | 45.31 |
| **6** | 18 | rs59499656 | N/A | A | -0.23 | 0.01 | 0.66 | 0.66 | 0.004 | 0.14 | 0.04 | 2.40E-09 | No | 0.0004 | 35.60 |
| **7** | 3 | rs6775319 | N/A | A | 0.23 | 0.002 | 0.27 | 0.28 | 0.004 | 0.58 | 0.04 | 3.50E-08 | No | 0.0003 | 30.43 |
| **8** | 5 | rs9293503 | N/A | T | 0.33 | 0.004 | 0.89 | 0.88 | 0.005 | 0.51 | 0.06 | 2.10E-08 | No | 0.0003 | 31.42 |

**Table 3B: Cochran's Q test**

| Method | Q | df | p value |
|---|---|---|---|
| MR Egger | 15.37 | 6 | 0.02 |
| IVW | 15.46 | 7 | 0.03 |

**Table 3C: MR-Egger intercept test**

| Egger intercept | SE | p value |
|---|---|---|
| 0.002 | 0.01 | 0.86 |

**Table 3D: I^2 statistic**

| | |
|---|---|
| **I^2** | 0.97 |

**Table 3E: F statistic for the whole instrument**

| | |
|---|---|
| **F statistic** | 33.11 |

**Table 3F: MR-PRESSO global test**

| Global test | p value | Outlier index | Outlier test | Distortion test p value |
|---|---|---|---|---|
| 21.57 | 0.03 | 5 | p=0.008 for index 9 | 0.002 |

**Table 3G: Causal estimates**

| Method | Snps (n) | Beta | SE | p value |
|---|---|---|---|---|
| IVW (fe) | 8 | -0.01 | 0.01 | 0.18 |
| IVW (mre) | 8 | -0.01 | 0.01 | 0.37 |
| MR Egger | 8 | -0.02 | 0.04 | 0.72 |
| Weighted median | 8 | 0.002 | 0.01 | 0.82 |

**Table 3H: Leave one out with multiplicative random effects IVW**

| SNP removed | Beta | SE | p value |
|---|---|---|---|
| rs11012732 | -0.00897 | 0.009894704 | 0.364522 |
| rs12522261 | -0.01037 | 0.009361873 | 0.2677774 |
| rs148193266 | -0.00914 | 0.009742485 | 0.3483083 |
| rs34517439 | -0.00846 | 0.009773026 | 0.3865476 |
| rs55725840* | 0.00136 | 0.004848901 | 0.7791853 |
| rs59499656 | -0.00533 | 0.009645179 | 0.5808343 |
| rs6775319 | -0.01004 | 0.009539118 | 0.2927191 |
| rs9293503 | -0.0104 | 0.009492063 | 0.2733435 |

* SNP identified as an outlier by MR-PRESSO, when removed the level of significance is reduced.

**Liberal instrument, SNP- average accelerations p<1E-06**

**Table 3I: SNPs identified after clumping and inclusion of proxy variants**

| Index | hromoson | SNP | Proxy SNP | Effect Allele | ccelerations be | ALS beta | ccelerations EA | ALS EAF | ALS SE | LS p valu | ccelerations S | celeration p va | romic alleles | PVE | F statistic |
|---|---|---|---|---|---|---|---|---|---|---|---|---|---|---|---|
| 1 | 12 | rs10880697 | N/A | G | -0.20 | -0.002 | 0.66 | 0.68 | 0.004 | 0.67 | 0.04 | 1.20E-07 | No | 0.0003 | 28 |
| 2 | 10 | rs11012732 | N/A | A | 0.22 | 0.000 | 0.67 | 0.65 | 0.004 | 0.97 | 0.04 | 5.40E-09 | No | 0.0004 | 34.04 |
| 3 | 16 | rs11865683 | N/A | C | -0.18 | -0.006 | 0.53 | 0.50 | 0.003 | 0.08 | 0.04 | 4.40E-07 | Yes | 0.0003 | 25.52 |

| | | | | | | | | | | | | | | | |
|---|---|---|---|---|---|---|---|---|---|---|---|---|---|---|---|
| **4** | 1 | rs12045968 | N/A | T | -0.24 | 0.002 | 0.78 | 0.78 | 0.004 | 0.59 | 0.04 | 5.10E-08 | No | 0.0003 | 29.66 |
| **5** | 5 | rs12522261 | N/A | G | 0.21 | 0.003 | 0.66 | 0.66 | 0.004 | 0.45 | 0.04 | 3.90E-08 | No | 0.0003 | 30.21 |
| **6** | 9 | rs1268539 | N/A | C | -0.19 | -0.002 | 0.58 | 0.58 | 0.004 | 0.58 | 0.04 | 2.70E-07 | No | 0.0003 | 26.43 |
| **7** | 11 | rs148193266 | N/A | A | -0.51 | -0.002 | 0.96 | 0.96 | 0.009 | 0.85 | 0.09 | 3.10E-08 | No | 0.0003 | 30.67 |
| **8** | 15 | rs1550435 | N/A | T | 0.20 | 0.000 | 0.57 | 0.56 | 0.003 | 0.91 | 0.04 | 5.00E-08 | No | 0.0003 | 29.72 |
| **9** | 18 | rs1668835 | N/A | T | -0.20 | -0.003 | 0.69 | 0.69 | 0.004 | 0.38 | 0.04 | 4.80E-07 | No | 0.0003 | 25.36 |
| **10** | 1 | rs34517439 | N/A | C | 0.31 | -0.0004 | 0.88 | 0.89 | 0.006 | 0.95 | 0.06 | 4.40E-08 | No | 0.0003 | 29.97 |
| **11** | 17 | rs56194509 | rs55725840 | T | -0.29 | 0.015 | 0.78 | 0.78 | 0.004 | 0.0002 | 0.04 | 1.70E-11 | No | 0.0005 | 45.31 |
| **12** | 18 | rs59499656 | N/A | A | -0.23 | 0.005 | 0.66 | 0.66 | 0.004 | 0.14 | 0.04 | 2.40E-09 | No | 0.0004 | 35.60 |
| **13** | 20 | rs6081105 | N/A | A | -0.18 | 0.006 | 0.40 | 0.39 | 0.004 | 0.13 | 0.04 | 8.20E-07 | No | 0.0003 | 24.31 |
| **14** | 2 | rs62168614 | N/A | G | 0.20 | -0.004 | 0.71 | 0.70 | 0.004 | 0.33 | 0.04 | 6.00E-07 | No | 0.0003 | 24.90 |
| **15** | 1 | rs6541235 | N/A | C | 0.25 | -0.007 | 0.85 | 0.85 | 0.005 | 0.16 | 0.05 | 7.30E-07 | No | 0.0003 | 24.53 |
| **16** | 3 | rs6775319 | N/A | A | 0.23 | 0.002 | 0.27 | 0.28 | 0.004 | 0.58 | 0.04 | 3.50E-08 | No | 0.0003 | 30.43 |
| **17** | 1 | rs791273 | N/A | A | -0.22 | 0.002 | 0.75 | 0.74 | 0.004 | 0.65 | 0.04 | 1.10E-07 | No | 0.0003 | 28.16 |
| **18** | 5 | rs9293503 | N/A | T | 0.33 | 0.004 | 0.89 | 0.88 | 0.005 | 0.51 | 0.06 | 2.10E-08 | No | 0.0003 | 31.42 |
| **19** | 6 | rs945890 | N/A | A | -0.21 | 0.002 | 0.29 | 0.28 | 0.004 | 0.58 | 0.04 | 2.20E-07 | No | 0.0003 | 26.89 |
| **20** | 16 | rs9938281 | N/A | A | -0.19 | -0.007 | 0.47 | 0.46 | 0.003 | 0.04 | 0.04 | 1.50E-07 | No | 0.0003 | 27.65 |

**Table 3J: Cochran's Q test**

| Method | Q | df | p value |
|---|---|---|---|
| MR Egger | 25.94 | 17 | 0.08 |
| IVW | 27.15 | 18 | 0.08 |

**Table 3K: MR-Egger intercept test**

| Egger intercept | se | pval |
|---|---|---|
| 0.005 | 0.006 | 0.39 |

**Table 3L: I^2 statistic**

| | |
|---|---|
| **I^2** | 0.97 |

**Table 3M: F statistic for the whole instrument**

| | |
|---|---|
| **F statistic** | 29.9 |

**Table 3N: MR-PRESSO global test**

| Global test | p value | Outlier index | Outlier test | Distortion test p value |
|---|---|---|---|---|
| 30.92 | 0.07 | N/A | N/A | N/A |

**Table 3O: Causal estimates**

| Method | Snps (n) | Beta | SE | p value |
|---|---|---|---|---|
| IVW (fe) | 19 | -0.005 | 0.004 | 0.21 |
| IVW (mre) | 19 | -0.005 | 0.005 | 0.31 |
| MR Egger | 19 | -0.027 | 0.02 | 0.30 |
| Weighted median | 19 | -0.001 | 0.01 | 0.87 |

**Table 3P: Leave one out with fixed effects IVW**

| SNP removed | Beta | SE | p value |
|---|---|---|---|
| rs10880697 | -0.006 | 0.004 | 0.17 |
| rs11012732 | -0.005 | 0.004 | 0.20 |
| rs12045968 | -0.005 | 0.004 | 0.25 |
| rs12522261 | -0.006 | 0.004 | 0.14 |
| rs1268539 | -0.006 | 0.004 | 0.16 |
| rs148193266 | -0.005 | 0.004 | 0.19 |
| rs1550435 | -0.005 | 0.004 | 0.21 |
| rs1668835 | -0.006 | 0.004 | 0.14 |
| rs34517439 | -0.005 | 0.004 | 0.21 |
| rs55725840 | -0.001 | 0.004 | 0.84 |
| rs59499656 | -0.004 | 0.004 | 0.37 |
| rs6081105 | -0.004 | 0.004 | 0.33 |
| rs62168614 | -0.004 | 0.004 | 0.29 |
| rs6541235 | -0.004 | 0.004 | 0.33 |
| rs6775319 | -0.006 | 0.004 | 0.16 |
| rs791273 | -0.005 | 0.004 | 0.24 |
| rs9293503 | -0.006 | 0.004 | 0.15 |

| | | | |
|---|---|---|---|
| rs945890 | -0.005 | 0.004 | 0.25 |
| rs9938281 | -0.007 | 0.004 | 0.08 |

**Supplementary Table 4: Supplementary content relating to the MR analysis of the SSOE - body fat percentage relationship.**

**Key**: SNP = single nucleotide polymorphism, PVE = proportion of variance explained, SE = standard error, EAF = effect allele frequency , ALS = amyotrophic lateral sclerosis , SSOE = strenuous sport or other exercise, VPA = vigorous physical activity, MVPA = moderate-vigorous physical activity, EA = educational attainment, BF% = body fat percentage, Chrom = chromosome, IVW = Inverse variance weighted, df = degrees of freedom, fe = fixed effects, mre = multiplicative random effects

**Conservative instrument, SNP-SSOE p<5E-08**

**Table 4A: SNPs identified after clumping and inclusion of proxy variants**

| Index | Chrom | SNP | Proxy SNP | Effect Allele | SSOE beta | BF% beta | SSOE EAF | BF% EAF | BF% SE | BF% p value | SSOE SE | SSOE p value | Palindromic | PVE | F statistic |
|---|---|---|---|---|---|---|---|---|---|---|---|---|---|---|---|
| **1** | 6 | rs10946808 | N/A | A | -0.008 | 0.02 | 0.73 | 0.67 | 0.005 | 0.0005 | 0.001 | 9.90E-10 | No | 0.0004 | 37.34 |
| **2** | 1 | rs1200154 | rs1200159 | T | -0.006 | 0.002 | 0.41 | 0.42 | 0.0047 | 0.69 | 0.001 | 1.10E-07 | No | 0.0003 | 28.16 |
| **3** | 6 | rs1265178 | N/A | G | 0.007 | 0.001 | 0.76 | 0.80 | 0.0067 | 0.93 | 0.001 | 3.20E-08 | No | 0.0003 | 30.57 |
| **4** | 5 | rs159544 | rs173946 | T | -0.007 | 0.003 | 0.61 | 0.60 | 0.0056 | 0.58 | 0.001 | 1.40E-09 | No | 0.0004 | 36.64 |
| **5** | 14 | rs75930676 | rs17767761 | C | -0.015 | -0.02 | 0.95 | 0.96 | 0.0172 | 0.26 | 0.003 | 5.10E-08 | No | 0.0003 | 29.69 |
| **6** | 2 | rs288070 | N/A | G | -0.01 | -0.02 | 0.90 | 0.88 | 0.0869 | 0.85 | 0.002 | 1.90E-08 | No | 0.0003 | 31.57 |
| **7** | 13 | rs4411372 | N/A | T | -0.007 | 0.01 | 0.72 | 0.72 | 0.0059 | 0.03 | 0.001 | 2.00E-08 | No | 0.0003 | 31.46 |
| **8** | 5 | rs4865667 | N/A | C | 0.007 | -0.005 | 0.61 | 0.61 | 0.0055 | 0.34 | 0.001 | 1.00E-08 | No | 0.0003 | 32.77 |
| **9** | 3 | rs7627864 | N/A | C | 0.007 | -0.01 | 0.57 | 0.61 | 0.0056 | 0.01 | 0.001 | 7.60E-09 | Yes | 0.0003 | 33.37 |
| **10** | 19 | rs111901094 | rs8182472 | T | 0.006 | 0.004 | 0.88 | 0.88 | 0.007 | 0.57 | 0.002 | 7.80E-04 | No | 0.0001 | 11.29 |
| **11** | 6 | rs896302 | N/A | C | 0.007 | -0.01 | 0.29 | 0.29 | 0.006 | 0.11 | 0.001 | 1.70E-08 | No | 0.0003 | 31.76 |
| **12** | 17 | rs166840 | rs9914273 | T | 0.007 | -0.003 | 0.60 | 0.61 | 0.0047 | 0.50 | 0.001 | 2.40E-10 | No | 0.0004 | 40.12 |

**Table 4B: Cochran's Q test**

| Method | Q | grees of freedo | p value |
|---|---|---|---|
| MR Egger | 13.43 | 9 | 0.14 |
| IVW | 13.67 | 10 | 0.19 |

**Table 4C: MR-Egger intercept test**

| Egger intercept | SE | p value |
|---|---|---|
| -0.006 | 0.02 | 0.7 |

**Table 4D: I^2 statistic**

| | |
|---|---|
| **I^2** | 0.97 |

**Table 4E: F statistic for the whole instrument**

| | |
|---|---|
| **F statistic** | 108.7 |

**Table 4F: MR-PRESSO global test**

| Global test | p value | Outlier index | Outlier test | Distortion test p value |
|---|---|---|---|---|
| 17.22 | 0.191 | N/A | N/A | N/A |

**Table 4G: Causal estimates**

| Method | Total SNPs | Beta | SE | p value |
|---|---|---|---|---|
| IVW (fe) | 11 | -0.78 | 0.26 | 0.003 |
| IVW (mre) | 11 | -0.78 | 0.30 | 0.01 |
| MR Egger | 11 | 0.07 | 2.15 | 0.97 |
| Weighted median | 11 | -0.44 | 0.37 | 0.23 |

**Table 4H: Leave one out with fixed effects IVW**

| SNP removed | Beta | SE | p value |
|---|---|---|---|
| rs10946808 | -0.50 | 0.28 | 0.08 |
| rs1200159 | -0.84 | 0.28 | 0.002 |
| rs1265178 | -0.86 | 0.27 | 0.002 |
| rs173946 | -0.82 | 0.27 | 0.003 |
| rs17767761 | -0.89 | 0.27 | 0.001 |
| rs288070 | -0.78 | 0.26 | 0.003 |
| rs4411372 | -0.67 | 0.27 | 0.01 |
| rs4865667 | -0.78 | 0.27 | 0.004 |
| rs8182472 | -0.85 | 0.27 | 0.001 |
| rs896302 | -0.72 | 0.27 | 0.01 |
| rs9914273 | -0.85 | 0.28 | 0.003 |

**Liberal instrument, SNP-SSOE p<1E-06**

**Table 4I: SNPs identified after clumping and inclusion of proxy variants**

| Index | Chrom | SNP | Proxy SNP | Effect Allele | SSOE beta | BF% beta | SSOE EAF | BF% EAF | BF% SE | BF% p value | SSOE SE | SSOE p value | Palindromic | PVE | F statistic |
|---|---|---|---|---|---|---|---|---|---|---|---|---|---|---|---|
| **1** | 7 | rs62472745 | rs10225928 | A | 0.007 | 0.0009 | 0.20 | 0.24 | 0.007 | 0.89 | 0.001 | 2.00E-06 | No | 0.0002 | 22.56 |
| **2** | 6 | rs10946808 | N/A | A | -0.008 | 0.018 | 0.73 | 0.67 | 0.005 | 0.0005 | 0.001 | 9.90E-10 | No | 0.0004 | 37.34 |
| **3** | 10 | rs11000785 | N/A | T | 0.006 | 0.013 | 0.28 | 0.30 | 0.030 | 0.67 | 0.001 | 2.60E-07 | No | 0.0003 | 26.54 |
| **4** | 1 | rs11210230 | N/A | T | -0.009 | 0.002 | 0.88 | 0.88 | 0.009 | 0.80 | 0.002 | 1.30E-07 | No | 0.0003 | 27.89 |
| **5** | 14 | rs11625806 | N/A | G | -0.006 | -0.001 | 0.59 | 0.62 | 0.006 | 0.80 | 0.001 | 6.80E-07 | No | 0.0002 | 24.68 |
| **6** | 5 | rs13180692 | rs11954477 | G | -0.006 | -0.0001 | 0.55 | 0.54 | 0.005 | 0.98 | 0.001 | 7.70E-07 | No | 0.0002 | 24.42 |
| **7** | 1 | rs1200154 | rs1200159 | T | -0.006 | 0.002 | 0.41 | 0.42 | 0.005 | 0.69 | 0.001 | 1.10E-07 | No | 0.0003 | 28.16 |
| **8** | 5 | rs35786746 | rs12187632 | G | 0.007 | 0.011 | 0.81 | 0.82 | 0.007 | 0.13 | 0.001 | 8.00E-07 | No | 0.0002 | 24.35 |
| **9** | 6 | rs1265178 | N/A | G | 0.007 | 0.001 | 0.76 | 0.80 | 0.007 | 0.93 | 0.001 | 3.20E-08 | No | 0.0003 | 30.57 |
| **10** | 12 | rs12821008 | N/A | C | -0.006 | -0.004 | 0.59 | 0.60 | 0.006 | 0.51 | 0.001 | 1.60E-07 | No | 0.0003 | 27.50 |
| **11** | 6 | rs13207819 | rs62422094 | G | -0.007 | 0.010 | 0.76 | 0.77 | 0.007 | 0.13 | 0.001 | 5.00E-07 | No | 0.0003 | 25.26 |
| **12** | 11 | rs551243 | rs1625595 | C | 0.006 | -0.002 | 0.53 | 0.50 | 0.006 | 0.79 | 0.001 | 1.30E-07 | No | 0.0003 | 27.80 |
| **13** | 5 | rs159544 | rs173946 | T | -0.007 | 0.003 | 0.61 | 0.60 | 0.006 | 0.58 | 0.001 | 1.40E-09 | No | 0.0004 | 36.64 |
| **14** | 14 | rs75930676 | rs17767761 | C | -0.02 | -0.02 | 0.95 | 0.96 | 0.017 | 0.26 | 0.003 | 5.10E-08 | No | 0.0003 | 29.69 |
| **15** | 11 | rs547118 | rs1786438 | T | 0.006 | -0.01 | 0.44 | 0.42 | 0.005 | 0.03 | 0.001 | 6.90E-07 | No | 0.0002 | 24.63 |
| **16** | 18 | rs1788826 | N/A | G | -0.006 | 0.02 | 0.35 | 0.34 | 0.006 | 0.003 | 0.001 | 7.60E-07 | No | 0.0002 | 24.45 |
| **17** | 9 | rs11143585 | rs1932202 | C | 0.007 | -0.005 | 0.75 | 0.76 | 0.007 | 0.43 | 0.001 | 3.70E-07 | No | 0.0003 | 25.85 |
| **18** | 20 | rs4811141 | rs2143625 | C | -0.008 | 0.02 | 0.13 | 0.13 | 0.008 | 0.03 | 0.002 | 1.70E-06 | No | 0.0002 | 22.95 |
| **19** | 10 | rs2148201 | N/A | G | -0.006 | -0.001 | 0.51 | 0.53 | 0.006 | 0.86 | 0.001 | 4.10E-07 | No | 0.0003 | 25.64 |
| **20** | 17 | rs216211 | N/A | A | 0.007 | 0.004 | 0.25 | 0.26 | 0.032 | 0.91 | 0.001 | 4.60E-07 | No | 0.0003 | 25.41 |
| **21** | 12 | rs2238046 | N/A | A | -0.006 | 0.007 | 0.61 | 0.62 | 0.006 | 0.21 | 0.001 | 7.40E-07 | No | 0.0002 | 24.51 |
| **22** | 7 | rs246743 | N/A | T | 0.008 | -0.008 | 0.15 | 0.16 | 0.009 | 0.35 | 0.002 | 5.50E-07 | No | 0.0002 | 25.07 |
| **23** | 15 | rs2470893 | N/A | C | 0.006 | -0.0001 | 0.67 | 0.69 | 0.006 | 0.98 | 0.001 | 5.50E-08 | No | 0.0003 | 29.54 |
| **24** | 3 | rs9881048 | rs2668196 | A | -0.007 | -0.01 | 0.19 | 0.20 | 0.008 | 0.14 | 0.001 | 9.00E-07 | No | 0.0002 | 24.12 |
| **25** | 8 | rs2725370 | rs2725371 | A | -0.007 | 0.001 | 0.30 | 0.28 | 0.02 | 0.95 | 0.001 | 1.10E-07 | No | 0.0003 | 28.22 |
| **26** | 2 | rs2871340 | N/A | G | -0.006 | 0.01 | 0.50 | 0.51 | 0.005 | 0.05 | 0.001 | 1.10E-07 | No | 0.0003 | 28.18 |
| **27** | 2 | rs288070 | N/A | G | -0.01 | -0.02 | 0.90 | 0.88 | 0.087 | 0.85 | 0.002 | 1.90E-08 | No | 0.0003 | 31.57 |
| **28** | 3 | rs336620 | rs400432 | A | 0.006 | -0.02 | 0.30 | 0.33 | 0.006 | 0.004 | 0.001 | 3.90E-07 | No | 0.0003 | 25.72 |
| **29** | 2 | rs4084828 | N/A | C | -0.006 | -0.006 | 0.46 | 0.47 | 0.007 | 0.44 | 0.001 | 4.50E-07 | No | 0.0003 | 25.45 |

| | | | | | | | | | | | | | | | |
|---|---|---|---|---|---|---|---|---|---|---|---|---|---|---|---|
| **30** | 13 | rs4411372 | N/A | T | -0.007 | 0.01 | 0.72 | 0.72 | 0.006 | 0.03 | 0.001 | 2.00E-08 | No | 0.0003 | 31.46 |
| **31** | 20 | rs118165004 | rs4809388 | C | 0.006 | -0.0003 | 0.83 | 0.83 | 0.008 | 0.97 | 0.001 | 1.40E-05 | No | 0.0002 | 18.81 |
| **32** | 5 | rs4865667 | N/A | C | 0.007 | -0.005 | 0.61 | 0.61 | 0.006 | 0.34 | 0.001 | 1.00E-08 | No | 0.0003 | 32.77 |
| **33** | 14 | rs11628052 | rs4900442 | C | -0.005 | -0.0008 | 0.54 | 0.52 | 0.006 | 0.89 | 0.001 | 1.20E-06 | No | 0.0002 | 23.55 |
| **34** | 10 | rs577525 | N/A | T | 0.006 | -0.006 | 0.44 | 0.47 | 0.005 | 0.25 | 0.001 | 2.70E-07 | No | 0.0003 | 26.43 |
| **35** | 4 | rs2749778 | rs6819202 | G | 0.006 | -0.006 | 0.40 | 0.39 | 0.006 | 0.26 | 0.001 | 1.40E-07 | No | 0.0003 | 27.79 |
| **36** | 8 | rs75276987 | rs7002919 | C | -0.010 | -0.003 | 0.92 | 0.92 | 0.010 | 0.75 | 0.002 | 2.40E-06 | No | 0.0002 | 22.24 |
| **37** | 9 | rs7031064 | N/A | A | -0.006 | 0.015 | 0.52 | 0.54 | 0.006 | 0.01 | 0.001 | 3.80E-07 | No | 0.0003 | 25.80 |
| **38** | 9 | rs7034243 | N/A | A | -0.006 | 0.004 | 0.66 | 0.65 | 0.006 | 0.48 | 0.001 | 1.00E-07 | No | 0.0003 | 28.34 |
| **39** | 1 | rs705692 | N/A | C | -0.009 | -0.001 | 0.11 | 0.11 | 0.009 | 0.87 | 0.002 | 6.30E-07 | No | 0.0002 | 24.81 |
| **40** | 3 | rs7627864 | N/A | C | 0.007 | -0.014 | 0.57 | 0.61 | 0.006 | 0.01 | 0.001 | 7.60E-09 | Yes | 0.0003 | 33.37 |
| **41** | 6 | rs763187 | N/A | A | -0.008 | 0.003 | 0.17 | 0.17 | 0.008 | 0.67 | 0.002 | 3.50E-07 | No | 0.0003 | 25.93 |
| **42** | 6 | rs7740107 | N/A | T | -0.006 | 0.006 | 0.26 | 0.26 | 0.006 | 0.33 | 0.001 | 6.10E-07 | No | 0.0002 | 24.88 |
| **43** | 4 | rs795973 | rs795980 | A | -0.006 | 0.007 | 0.67 | 0.67 | 0.005 | 0.17 | 0.001 | 1.00E-06 | No | 0.0002 | 23.86 |
| **44** | 19 | rs111901094 | rs8182472 | T | 0.006 | 0.004 | 0.88 | 0.88 | 0.007 | 0.57 | 0.002 | 7.80E-04 | No | 0.0001 | 11.29 |
| **45** | 6 | rs896302 | N/A | C | 0.007 | -0.01 | 0.29 | 0.29 | 0.006 | 0.11 | 0.001 | 1.70E-08 | No | 0.0003 | 31.76 |
| **46** | 7 | rs9177 | N/A | A | 0.006 | 0.0009 | 0.51 | 0.54 | 0.006 | 0.89 | 0.001 | 4.60E-07 | No | 0.0003 | 25.43 |
| **47** | 22 | rs9604803 | N/A | C | 0.006 | -0.002 | 0.67 | 0.66 | 0.006 | 0.69 | 0.001 | 8.00E-07 | No | 0.0002 | 24.36 |
| **48** | 17 | rs9903845 | N/A | C | 0.006 | 0.002 | 0.67 | 0.69 | 0.006 | 0.80 | 0.001 | 7.30E-07 | No | 0.0002 | 24.53 |
| **49** | 17 | rs166840 | rs9914273 | T | 0.007 | -0.003 | 0.60 | 0.61 | 0.005 | 0.50 | 0.001 | 2.40E-10 | No | 0.0004 | 40.12 |
| **50** | 18 | rs8093216 | rs9954933 | C | -0.006 | 0 | 0.65 | 0.65 | 0.006 | 1.00 | 0.001 | 3.10E-06 | No | 0.0002 | 21.76 |

**Table 4J: Cochran's Q test**

| **Method** | **Q** | **grees of freedo** | **p value** |
|---|---|---|---|
| MR Egger | 56.15 | 47 | 0.17 |
| rse variance weig | 56.94 | 48 | 0.18 |

**Table 4K: MR-Egger intercept**

| **Egger intercept** | **SE** | **p value** |
|---|---|---|
| -0.006 | 0.007 | 0.42 |

**Table 4L: I^2 statistic**

| | |
|---|---|
| **I^2** | 0.96 |

**Table 4M: F statistic for the whole instrument**

| | |
|---|---|
| **F statistic** | 93.46 |

**Table 4N: MR-PRESSO global test**

| Global test | p value | Outlier index | Outlier test | Distortion test p value |
|---|---|---|---|---|
| 59.65 | 0.18 | N/A | N/A | N/A |

**Table 4O: Causal estimates**

| Method | Total SNPs | Beta | SE | p value |
|---|---|---|---|---|
| IVW (fe) | 49 | -0.64 | 0.14 | 3.82E-06 |
| IVW (mre) | 49 | -0.64 | 0.15 | 2.21E-05 |
| MR Egger | 49 | 0.20 | 1.05 | 8.49E-01 |
| Weighted median | 49 | -0.44 | 0.21 | 3.20E-02 |

**Table 4P: Leave one out with fixed effects IVW**

| SNP removed | Beta | SE | p value |
|---|---|---|---|
| rs10225928 | -0.66 | 0.14 | 2.80E-06 |
| rs10946808 | -0.57 | 0.14 | 7.47E-05 |
| rs11000785 | -0.65 | 0.14 | 3.56E-06 |
| rs11210230 | -0.65 | 0.14 | 3.59E-06 |
| rs11625806 | -0.66 | 0.14 | 2.52E-06 |
| rs11954477 | -0.66 | 0.14 | 2.95E-06 |
| rs1200159 | -0.66 | 0.14 | 3.77E-06 |
| rs12187632 | -0.69 | 0.14 | 1.06E-06 |
| rs1265178 | -0.66 | 0.14 | 2.74E-06 |
| rs12821008 | -0.67 | 0.14 | 1.81E-06 |
| rs13207819 | -0.63 | 0.14 | 8.55E-06 |
| rs1625595 | -0.65 | 0.14 | 3.62E-06 |
| rs173946 | -0.65 | 0.14 | 4.34E-06 |

| | | | |
|---|---|---|---|
| rs17767761 | -0.67 | 0.14 | 1.63E-06 |
| rs1786438 | -0.61 | 0.14 | 1.36E-05 |
| rs1788826 | -0.60 | 0.14 | 2.28E-05 |
| rs1932202 | -0.64 | 0.14 | 5.24E-06 |
| rs2143625 | -0.61 | 0.14 | 1.30E-05 |
| rs2148201 | -0.66 | 0.14 | 2.66E-06 |
| rs216211 | -0.65 | 0.14 | 3.73E-06 |
| rs2238046 | -0.63 | 0.14 | 7.16E-06 |
| rs246743 | -0.64 | 0.14 | 5.66E-06 |
| rs2470893 | -0.66 | 0.14 | 2.84E-06 |
| rs2668196 | -0.68 | 0.14 | 1.27E-06 |
| rs2725371 | -0.65 | 0.14 | 3.78E-06 |
| rs2871340 | -0.62 | 0.14 | 1.28E-05 |
| rs288070 | -0.65 | 0.14 | 3.75E-06 |
| rs400432 | -0.60 | 0.14 | 2.25E-05 |
| rs4084828 | -0.67 | 0.14 | 2.14E-06 |
| rs4411372 | -0.61 | 0.14 | 1.52E-05 |
| rs4809388 | -0.65 | 0.14 | 3.40E-06 |
| rs4865667 | -0.64 | 0.14 | 6.00E-06 |
| rs4900442 | -0.66 | 0.14 | 2.84E-06 |
| rs577525 | -0.64 | 0.14 | 6.82E-06 |
| rs6819202 | -0.64 | 0.14 | 6.76E-06 |
| rs7002919 | -0.66 | 0.14 | 2.56E-06 |
| rs7031064 | -0.60 | 0.14 | 1.88E-05 |
| rs7034243 | -0.64 | 0.14 | 4.96E-06 |
| rs705692 | -0.66 | 0.14 | 2.68E-06 |
| rs763187 | -0.65 | 0.14 | 4.12E-06 |
| rs7740107 | -0.64 | 0.14 | 5.93E-06 |
| rs795980 | -0.63 | 0.14 | 8.39E-06 |
| rs8182472 | -0.66 | 0.14 | 2.40E-06 |
| rs896302 | -0.63 | 0.14 | 9.78E-06 |
| rs9177 | -0.66 | 0.14 | 2.91E-06 |

| | | | |
|---|---|---|---|
| rs9604803 | -0.65 | 0.14 | 4.00E-06 |
| rs9903845 | -0.66 | 0.14 | 2.56E-06 |
| rs9914273 | -0.65 | 0.14 | 4.61E-06 |
| rs9954933 | -0.66 | 0.14 | 3.10E-06 |

**Supplementary Table 5: Supplementary content relating to the MR analysis of the body fat percentage - ALS relationship.**

**Key**: SNP = single nucleotide polymorphism, PVE = proportion of variance explained, SE = standard error, EAF = effect allele frequency , ALS = amyotrophic lateral sclerosis , SSOE = strenuous sport or other exercise, VPA = vigorous physical activity, MVPA = moderate-vigorous physical activity, EA = educational attainment, BF% = body fat percentage, Chrom = chromosome, IVW = Inverse variance weighted, df = degrees of freedom, fe = fixed effects, mre = multiplicative random effects

**Conservative instrument, SNP-BF% p<5E-08**

**Table 7A: SNPs identified after clumping and inclusion of proxy variants**

| Index | Chrom | SNP | Proxy SNP | Effect Allele | BF% beta | ALS beta | BF% EAF | ALS EAF | ALS SE | ALS p value | BF% SE | BF% p value | Palindromic | PVE | F statistic |
|---|---|---|---|---|---|---|---|---|---|---|---|---|---|---|---|
| 1 | 16 | rs1558902 | N/A | A | 0.051 | -0.002308 | 0.40 | 0.41 | 0.0034948 | 0.508987 | 0.0047 | 3.79E-27 | No | 0.001 | 117.74 |
| 2 | 2 | rs2943652 | N/A | T | -0.0335 | -0.00812425 | 0.64 | 0.63 | 0.0035547 | 0.0222832 | 0.0047 | 1.53E-12 | No | 0.0005 | 50.80 |
| 3 | 16 | rs4788099 | N/A | A | -0.0269 | 0.000112037 | 0.62 | 0.62 | 0.0035324 | 0.974698 | 0.0047 | 1.21E-08 | No | 0.0003 | 32.76 |
| 4 | 18 | rs6567160 | N/A | T | -0.0341 | -0.00526367 | 0.75 | 0.76 | 0.0040042 | 0.188665 | 0.0053 | 1.33E-10 | No | 0.0004 | 41.40 |
| 5 | 2 | rs6738627 | N/A | A | 0.0304 | -0.00526836 | 0.37 | 0.37 | 0.0035462 | 0.137378 | 0.0052 | 5.68E-09 | No | 0.0003 | 34.18 |
| 6 | 2 | rs6755502 | N/A | T | -0.0392 | 0.00228258 | 0.17 | 0.18 | 0.0045073 | 0.612562 | 0.0061 | 1.41E-10 | No | 0.0004 | 41.30 |
| 7 | 19 | rs6857 | N/A | T | -0.048 | 0.00611514 | 0.16 | 0.16 | 0.004729 | 0.195974 | 0.0083 | 6.84E-09 | No | 0.0003 | 33.44 |
| 8 | 13 | rs693839 | N/A | T | -0.0283 | -0.000809125 | 0.68 | 0.69 | 0.0037231 | 0.827956 | 0.0049 | 6.56E-09 | No | 0.0003 | 33.36 |
| 9 | 17 | rs9906944 | N/A | T | -0.0333 | 0.00727855 | 0.33 | 0.33 | 0.003699 | 0.0491041 | 0.006 | 2.86E-08 | No | 0.0003 | 30.80 |

**Table 7B: Cochran's Q**

| Method | Q | df | p value |
|---|---|---|---|
| MR Egger | 14.26 | 7 | 0.05 |
| IVW | 14.77 | 8 | 0.06 |

**Table 7C: MR-Egger intercept**

| Egger intercept | SE | p value |
|---|---|---|
| 0.004 | 0.008 | 0.63 |

**Table 7D: I^2 statistic**

| | |
|---|---|
| **I^2** | 0.98 |

**Table 7E: F statistic for the whole instrument**

| | |
|---|---|
| **F statistic** | 44.94 |

**Table 7F: MR-PRESSO global test**

| Global test | p value | Outlier inde | Outlier test | Distortion test p value |
|---|---|---|---|---|
| 18.34 | 0.07 | N/A | N/A | N/A |

**Table 7G: Causal estimates**

| Method | Total SNPs | Beta | SE | p value |
|---|---|---|---|---|
| IVW (fe) | 9 | -0.03 | 0.03 | 0.41 |
| IVW (mre) | 9 | -0.03 | 0.05 | 0.55 |
| MR Egger | 9 | -0.14 | 0.23 | 0.56 |
| Weighted median | 9 | -0.05 | 0.05 | 0.33 |

**Table 7H: Leave one out with fixed effects IVW**

| SNP removed | Beta | SE | p value |
|---|---|---|---|
| rs1558902 | -0.02 | 0.04 | 0.58 |
| rs2943652 | -0.06 | 0.04 | 0.10 |
| rs4788099 | -0.03 | 0.04 | 0.40 |
| rs6567160 | -0.05 | 0.04 | 0.21 |
| rs6738627 | -0.01 | 0.04 | 0.69 |
| rs6755502 | -0.03 | 0.04 | 0.49 |
| rs6857 | -0.01 | 0.04 | 0.70 |
| rs693839 | -0.03 | 0.04 | 0.36 |
| rs9906944 | -0.01 | 0.04 | 0.83 |

**Liberal instrument, SNP-BF% p<1E-06**

**Table 7I: SNPs identified after clumping and inclusion of proxy variants**

| Index | Chrom | SNP | Proxy SNP | Effect Allele | BF% beta | ALS beta | BF% EAF | ALS EAF | ALS SE | ALS p value | BF% SE | BF% p value | Palindromic | PVE | F statistic |
|---|---|---|---|---|---|---|---|---|---|---|---|---|---|---|---|
| **1** | 4 | rs10938397 | N/A | A | -0.02 | -0.007 | 0.57 | 0.57 | 0.003 | 0.058 | 0.005 | 1.36E-07 | No | 0.0003 | 28.07 |
| **2** | 20 | rs1206773 | N/A | A | 0.03 | -0.004 | 0.25 | 0.26 | 0.004 | 0.271 | 0.005 | 1.36E-07 | No | 0.0003 | 28.11 |
| **3** | 1 | rs1278664 | N/A | T | -0.03 | -0.004 | 0.22 | 0.22 | 0.004 | 0.341 | 0.006 | 9.18E-07 | No | 0.0002 | 24.29 |

| | | | | | | | | | | | | | | | |
|---|---|---|---|---|---|---|---|---|---|---|---|---|---|---|---|
| **4** | 16 | rs1558902 | N/A | A | 0.05 | -0.002 | 0.40 | 0.41 | 0.003 | 0.509 | 0.005 | 3.79E-27 | No | 0.001 | 117.74 |
| **5** | 1 | rs1780050 | N/A | A | 0.03 | -0.003 | 0.59 | 0.58 | 0.003 | 0.400 | 0.005 | 1.49E-07 | No | 0.0003 | 27.34 |
| **6** | 2 | rs2943652 | N/A | T | -0.03 | -0.008 | 0.64 | 0.63 | 0.004 | 0.022 | 0.005 | 1.53E-12 | No | 0.0005 | 50.80 |
| **7** | 22 | rs3761445 | N/A | A | -0.02 | 0.007 | 0.59 | 0.59 | 0.003 | 0.035 | 0.005 | 1.75E-07 | No | 0.0003 | 26.95 |
| **8** | 16 | rs4788099 | N/A | A | -0.03 | 0.0001 | 0.62 | 0.62 | 0.004 | 0.975 | 0.005 | 1.21E-08 | No | 0.0003 | 32.76 |
| **9** | 18 | rs6567160 | N/A | T | -0.03 | -0.005 | 0.75 | 0.76 | 0.004 | 0.189 | 0.005 | 1.33E-10 | No | 0.0004 | 41.40 |
| **10** | 2 | rs6738627 | N/A | A | 0.03 | -0.005 | 0.37 | 0.37 | 0.004 | 0.137 | 0.005 | 5.68E-09 | No | 0.0003 | 34.18 |
| **11** | 2 | rs6755502 | N/A | T | -0.04 | 0.002 | 0.17 | 0.18 | 0.005 | 0.613 | 0.006 | 1.41E-10 | No | 0.0004 | 41.30 |
| **12** | 19 | rs6857 | N/A | T | -0.05 | 0.006 | 0.16 | 0.16 | 0.005 | 0.196 | 0.008 | 6.84E-09 | No | 0.0003 | 33.44 |
| **13** | 13 | rs693839 | N/A | T | -0.03 | -0.001 | 0.68 | 0.69 | 0.004 | 0.828 | 0.005 | 6.56E-09 | No | 0.0003 | 33.36 |
| **14** | 12 | rs7133378 | N/A | A | 0.03 | -0.001 | 0.32 | 0.33 | 0.004 | 0.750 | 0.005 | 2.31E-07 | No | 0.0003 | 26.39 |
| **15** | 17 | rs9906944 | N/A | T | -0.03 | 0.007 | 0.33 | 0.33 | 0.004 | 0.049 | 0.006 | 2.86E-08 | No | 0.0003 | 30.80 |
| **16** | 1 | rs990871 | N/A | T | 0.03 | -0.0003 | 0.62 | 0.61 | 0.004 | 0.930 | 0.005 | 5.87E-08 | No | 0.0003 | 29.34 |

**Table 7J: Cochran's Q test**

| **Method** | **Q** | **df** | **p value** |
|---|---|---|---|
| MR Egger | 25.22 | 14 | 0.03 |
| IVW | 25.37 | 15 | 0.05 |

**Table 5K: MR-Egger intercept test**

| **Egger intercept** | **SE** | **p value** |
|---|---|---|
| 0.002 | 0.01 | 0.78 |

**Table5L: I^2 statistic**

| | |
|---|---|
| **I^2** | 0.97 |

**Table 5M: F statistic for the whole instrument**

| | |
|---|---|
| **F statistic** | 37.79 |

**Table 5N: MR-PRESSO global test**

| **Global test** | **p value** | **Outlier inde** | **Outlier test** | **Distortion test p value** |
|---|---|---|---|---|

| | | | | |
|---|---|---|---|---|
| 28.68 | 0.04 | None | None | N/A |

**Table 5O: Causal estimates**

| Method | SNPs (n) | Beta | SE | p value |
|---|---|---|---|---|
| IVW (fe) | 16 | -0.03 | 0.03 | 0.3 |
| IVW (mre) | 16 | -0.03 | 0.04 | 0.43 |
| MR Egger | 16 | -0.08 | 0.16 | 0.65 |
| Weighted median | 16 | -0.05 | 0.04 | 0.31 |

**Table 5P: Leave one out with multiplicative random effects IVW**

| SNP removed | Beta | SE | p value |
|---|---|---|---|
| rs10938397 | -0.04 | 0.04 | 0.23 |
| rs1206773 | -0.02 | 0.04 | 0.54 |
| rs1278664 | -0.04 | 0.04 | 0.35 |
| rs1558902 | -0.03 | 0.04 | 0.54 |
| rs1780050 | -0.03 | 0.04 | 0.52 |
| rs2943652 | -0.05 | 0.03 | 0.13 |
| rs3761445 | -0.02 | 0.04 | 0.62 |
| rs4788099 | -0.03 | 0.04 | 0.44 |
| rs6567160 | -0.04 | 0.04 | 0.27 |
| rs6738627 | -0.02 | 0.04 | 0.60 |
| rs6755502 | -0.03 | 0.04 | 0.49 |
| rs6857 | -0.02 | 0.04 | 0.61 |
| rs693839 | -0.03 | 0.04 | 0.41 |
| rs7133378 | -0.03 | 0.04 | 0.47 |
| rs9906944 | -0.02 | 0.04 | 0.68 |
| rs990871 | -0.03 | 0.04 | 0.44 |

**Supplementary Table 6:** Supplementary content relating to the MR analysis of the educational attainment- ALS relationship.

**Key**: SNP = single nucleotide polymorphism, PVE = proportion of variance explained, SE = standard error, EAF = effect allele frequency , ALS = amyotrophic lateral sclerosis , SSOE = strenuous sport or other exercise, VPA = vigorous physical activity, MVPA = moderate-vigorous physical activity, EA = educational attainment, BF% = body fat percentage, Chrom = chromosome, IVW = Inverse variance weighted, df = degrees of freedom, fe = fixed effects, mre = multiplicative random effects

**Conservative instrument, SNP-EA p<5E-08. NOTE: Due to the large number of genome-wide significant SNPs and the power of the GWAS in question , a liberal instrument was not implemented in this aspect of the analysis.**

**Table 6A: SNPs identified after clumping and inclusion of proxy variants**

| Index | Chrom | SNP | Proxy SNP | Effect Allele | EA beta | ALS beta | EA EAF | ALS EAF | ALS SE | ALS p value | EA SE | EA p value | Palindromic | PVE | F statistic |
|---|---|---|---|---|---|---|---|---|---|---|---|---|---|---|---|
| 1 | 5 | rs10073890 | N/A | G | -0.01 | -0.002 | 0.74 | 0.74 | 0.004 | 0.59 | 0.002 | 1.11E-10 | No | 3.66E-05 | 41.46 |
| 2 | 1 | rs1008078 | N/A | T | -0.02 | 0.004 | 0.41 | 0.40 | 0.004 | 0.21 | 0.002 | 1.20E-23 | No | 8.92E-05 | 100.93 |
| 3 | 2 | rs10189857 | N/A | G | -0.02 | 0.004 | 0.42 | 0.43 | 0.003 | 0.26 | 0.002 | 6.70E-24 | No | 8.99E-05 | 101.76 |
| 4 | 2 | rs10205801 | N/A | A | -0.01 | -0.0004 | 0.51 | 0.55 | 0.003 | 0.90 | 0.002 | 7.17E-10 | No | 3.35E-05 | 37.92 |
| 5 | 7 | rs10215082 | N/A | A | -0.01 | 0.0004 | 0.44 | 0.46 | 0.004 | 0.91 | 0.002 | 3.33E-14 | No | 5.07E-05 | 57.39 |
| 6 | 7 | rs10240905 | N/A | T | -0.01 | -0.002 | 0.33 | 0.36 | 0.004 | 0.54 | 0.002 | 3.79E-11 | No | 3.84E-05 | 43.47 |
| 7 | 6 | rs10456918 | N/A | A | -0.01 | 0.005 | 0.82 | 0.83 | 0.005 | 0.29 | 0.002 | 3.67E-11 | No | 3.88E-05 | 43.95 |
| 8 | 18 | rs10460095 | N/A | G | 0.01 | -0.002 | 0.41 | 0.42 | 0.003 | 0.48 | 0.002 | 4.87E-10 | No | 3.43E-05 | 38.86 |
| 9 | 9 | rs1051474 | N/A | T | -0.01 | 0.003 | 0.73 | 0.72 | 0.004 | 0.39 | 0.002 | 4.86E-12 | No | 4.23E-05 | 47.89 |
| 10 | 9 | rs10760023 | N/A | C | -0.01 | 0.001 | 0.67 | 0.66 | 0.004 | 0.87 | 0.002 | 2.12E-09 | No | 3.16E-05 | 35.80 |
| 11 | 11 | rs10765775 | N/A | A | 0.01 | 0.003 | 0.40 | 0.39 | 0.004 | 0.45 | 0.002 | 2.62E-17 | No | 6.31E-05 | 71.48 |
| 12 | 12 | rs10772644 | N/A | G | -0.02 | -0.003 | 0.11 | 0.11 | 0.006 | 0.62 | 0.003 | 1.50E-09 | No | 3.23E-05 | 36.54 |
| 13 | 12 | rs10773002 | N/A | T | -0.02 | 0.001 | 0.72 | 0.75 | 0.004 | 0.90 | 0.002 | 8.68E-29 | No | 0.0001 | 123.69 |
| 14 | 2 | rs10856785 | N/A | T | -0.01 | 0.001 | 0.73 | 0.73 | 0.004 | 0.79 | 0.002 | 3.83E-09 | No | 3.07E-05 | 34.76 |
| 15 | 12 | rs10862376 | N/A | T | -0.02 | -0.007 | 0.86 | 0.85 | 0.005 | 0.13 | 0.002 | 1.40E-11 | No | 4.04E-05 | 45.72 |
| 16 | 1 | rs10875121 | N/A | G | -0.02 | -0.004 | 0.14 | 0.16 | 0.005 | 0.44 | 0.002 | 5.53E-16 | No | 5.82E-05 | 65.85 |
| 17 | 10 | rs10887801 | N/A | G | -0.01 | -0.002 | 0.56 | 0.57 | 0.003 | 0.62 | 0.002 | 2.27E-10 | No | 3.57E-05 | 40.41 |
| 18 | 5 | rs10940921 | N/A | G | -0.01 | -0.009 | 0.57 | 0.59 | 0.003 | 0.01 | 0.002 | 7.00E-10 | No | 3.34E-05 | 37.85 |
| 19 | 9 | rs10963297 | N/A | G | 0.02 | -0.004 | 0.25 | 0.23 | 0.004 | 0.36 | 0.002 | 7.36E-22 | No | 8.17E-05 | 92.47 |
| 20 | 10 | rs10994777 | N/A | G | -0.01 | 0.003 | 0.86 | 0.84 | 0.005 | 0.50 | 0.002 | 3.36E-10 | No | 3.50E-05 | 39.60 |
| 21 | 11 | rs11023749 | N/A | A | 0.01 | 0.001 | 0.67 | 0.67 | 0.004 | 0.87 | 0.002 | 2.96E-10 | No | 3.49E-05 | 39.55 |
| 22 | 9 | rs1105307 | N/A | A | -0.01 | 0.000 | 0.24 | 0.27 | 0.004 | 1.00 | 0.002 | 1.67E-09 | No | 3.20E-05 | 36.18 |
| 23 | 2 | rs1106090 | N/A | A | 0.01 | -0.002 | 0.63 | 0.62 | 0.004 | 0.55 | 0.002 | 2.09E-11 | No | 3.97E-05 | 44.93 |

| | | | | | | | | | | | | | | | |
|---|---|---|---|---|---|---|---|---|---|---|---|---|---|---|---|
| 24 | 18 | rs11081529 | N/A | T | 0.01 | 0.007 | 0.74 | 0.73 | 0.004 | 0.06 | 0.002 | 1.82E-12 | No | 4.39E-05 | 49.68 |
| 25 | 2 | rs11123818 | N/A | G | -0.02 | 0.001 | 0.61 | 0.63 | 0.004 | 0.82 | 0.002 | 1.72E-32 | No | 0.0001 | 141.41 |
| 26 | 9 | rs111821073 | N/A | C | -0.01 | 0.002 | 0.84 | 0.84 | 0.005 | 0.74 | 0.002 | 4.85E-09 | No | 3.02E-05 | 34.15 |
| 27 | 11 | rs11222609 | N/A | G | -0.01 | -0.001 | 0.34 | 0.36 | 0.004 | 0.77 | 0.002 | 1.05E-09 | No | 3.28E-05 | 37.09 |
| 28 | 4 | rs112687095 | N/A | G | -0.01 | 0.009 | 0.84 | 0.83 | 0.005 | 0.05 | 0.002 | 2.42E-08 | No | 2.74E-05 | 30.99 |
| 29 | 2 | rs112806496 | N/A | G | 0.02 | 0.009 | 0.09 | 0.09 | 0.006 | 0.14 | 0.003 | 8.28E-10 | No | 3.32E-05 | 37.59 |
| 30 | 7 | rs113520408 | N/A | A | 0.01 | -0.008 | 0.29 | 0.27 | 0.004 | 0.03 | 0.002 | 1.02E-11 | No | 4.08E-05 | 46.13 |
| 31 | 7 | rs113615161 | N/A | T | -0.01 | 0.002 | 0.14 | 0.13 | 0.005 | 0.76 | 0.003 | 3.97E-09 | No | 3.06E-05 | 34.67 |
| 32 | 11 | rs1143770 | N/A | C | -0.01 | 0.002 | 0.41 | 0.42 | 0.003 | 0.48 | 0.002 | 4.31E-11 | No | 3.85E-05 | 43.62 |
| 33 | 3 | rs115000530 | N/A | T | 0.03 | 0.01 | 0.06 | 0.06 | 0.007 | 0.11 | 0.004 | 3.30E-14 | No | 5.09E-05 | 57.62 |
| 34 | 4 | rs115454970 | N/A | T | -0.01 | -0.01 | 0.30 | 0.26 | 0.004 | 0.001 | 0.002 | 2.76E-09 | No | 3.13E-05 | 35.46 |
| 35 | 11 | rs11601122 | N/A | G | -0.02 | -0.003 | 0.15 | 0.16 | 0.005 | 0.59 | 0.002 | 2.24E-17 | No | 6.33E-05 | 71.66 |
| 36 | 13 | rs11620355 | N/A | G | -0.02 | 0.005 | 0.88 | 0.91 | 0.006 | 0.42 | 0.003 | 4.77E-09 | No | 3.03E-05 | 34.26 |
| 37 | 14 | rs11627087 | N/A | A | 0.02 | -0.002 | 0.91 | 0.93 | 0.007 | 0.73 | 0.003 | 3.71E-08 | No | 2.67E-05 | 30.27 |
| 38 | 15 | rs11635092 | N/A | A | -0.01 | -0.003 | 0.36 | 0.36 | 0.004 | 0.36 | 0.002 | 3.89E-12 | No | 4.27E-05 | 48.37 |
| 39 | 17 | rs11657342 | N/A | A | 0.01 | 0.003 | 0.36 | 0.38 | 0.004 | 0.37 | 0.002 | 1.94E-13 | No | 4.77E-05 | 54.03 |
| 40 | 18 | rs11663602 | N/A | A | -0.01 | -0.008 | 0.26 | 0.26 | 0.004 | 0.05 | 0.002 | 1.64E-10 | No | 3.60E-05 | 40.76 |
| 41 | 2 | rs11678980 | N/A | G | 0.02 | 0.008 | 0.55 | 0.55 | 0.004 | 0.03 | 0.002 | 4.29E-24 | No | 9.08E-05 | 102.81 |
| 42 | 2 | rs11681861 | N/A | G | -0.01 | -0.007 | 0.16 | 0.14 | 0.005 | 0.19 | 0.003 | 2.88E-08 | No | 2.71E-05 | 30.70 |
| 43 | 2 | rs11694904 | N/A | T | 0.01 | -0.005 | 0.34 | 0.31 | 0.004 | 0.15 | 0.002 | 4.78E-11 | No | 3.81E-05 | 43.13 |
| 44 | 4 | rs11732657 | N/A | A | -0.01 | 0.004 | 0.70 | 0.75 | 0.004 | 0.31 | 0.002 | 9.54E-11 | No | 3.69E-05 | 41.82 |
| 45 | 6 | rs11752914 | N/A | T | 0.01 | 0.003 | 0.80 | 0.81 | 0.004 | 0.54 | 0.002 | 2.13E-08 | No | 2.76E-05 | 31.28 |
| 46 | 7 | rs11772580 | N/A | G | 0.01 | 0.009 | 0.75 | 0.75 | 0.004 | 0.03 | 0.002 | 2.57E-09 | No | 3.14E-05 | 35.58 |
| 47 | 15 | rs117799466 | N/A | G | -0.01 | -0.001 | 0.62 | 0.61 | 0.004 | 0.83 | 0.002 | 2.91E-09 | No | 3.10E-05 | 35.10 |
| 48 | 17 | rs11871429 | N/A | A | 0.01 | -0.003 | 0.80 | 0.77 | 0.004 | 0.44 | 0.002 | 1.92E-12 | No | 4.40E-05 | 49.77 |
| 49 | 1 | rs12028010 | N/A | C | -0.02 | 0.003 | 0.22 | 0.23 | 0.004 | 0.50 | 0.002 | 4.51E-17 | No | 6.23E-05 | 70.49 |
| 50 | 1 | rs12134151 | N/A | C | -0.01 | 0.002 | 0.52 | 0.51 | 0.003 | 0.61 | 0.002 | 2.42E-13 | Yes | 4.74E-05 | 53.63 |
| 51 | 9 | rs12375949 | N/A | T | -0.01 | 0.002 | 0.43 | 0.43 | 0.003 | 0.52 | 0.002 | 3.31E-17 | No | 6.25E-05 | 70.77 |
| 52 | 2 | rs12468040 | N/A | G | -0.01 | 0.005 | 0.60 | 0.62 | 0.004 | 0.20 | 0.002 | 2.46E-16 | No | 5.92E-05 | 66.96 |
| 53 | 4 | rs12503522 | N/A | C | 0.01 | 0.004 | 0.75 | 0.72 | 0.004 | 0.36 | 0.002 | 2.24E-09 | No | 3.16E-05 | 35.81 |
| 54 | 5 | rs12519073 | N/A | C | 0.01 | -0.006 | 0.76 | 0.76 | 0.004 | 0.12 | 0.002 | 1.60E-09 | No | 3.23E-05 | 36.54 |
| 55 | 11 | rs12574281 | N/A | A | -0.01 | 0.000 | 0.60 | 0.64 | 0.004 | 0.89 | 0.002 | 8.85E-10 | No | 3.31E-05 | 37.45 |

| | | | | | | | | | | | | | | | |
|---|---|---|---|---|---|---|---|---|---|---|---|---|---|---|---|
| 56 | 17 | rs12602286 | N/A | T | 0.02 | -0.001 | 0.89 | 0.87 | 0.005 | 0.85 | 0.003 | 2.37E-11 | No | 3.93E-05 | 44.50 |
| 57 | 4 | rs12643771 | N/A | C | -0.02 | 0.001 | 0.69 | 0.68 | 0.004 | 0.72 | 0.002 | 1.61E-16 | No | 6.01E-05 | 68.06 |
| 58 | 4 | rs12647336 | N/A | A | -0.01 | -0.004 | 0.86 | 0.85 | 0.005 | 0.36 | 0.002 | 4.82E-09 | No | 3.02E-05 | 34.23 |
| 59 | 5 | rs12655753 | N/A | A | -0.03 | -0.013 | 0.02 | 0.02 | 0.011 | 0.23 | 0.005 | 1.18E-08 | No | 2.87E-05 | 32.53 |
| 60 | 9 | rs12682775 | N/A | T | -0.01 | -0.001 | 0.79 | 0.78 | 0.004 | 0.86 | 0.002 | 5.99E-09 | No | 2.99E-05 | 33.86 |
| 61 | 11 | rs12804787 | N/A | A | 0.02 | -0.003 | 0.93 | 0.93 | 0.007 | 0.67 | 0.003 | 2.96E-08 | No | 2.72E-05 | 30.77 |
| 62 | 10 | rs1291818 | N/A | C | -0.01 | 0.002 | 0.52 | 0.48 | 0.003 | 0.51 | 0.002 | 1.78E-10 | No | 3.60E-05 | 40.73 |
| 63 | 17 | rs12940014 | N/A | C | 0.01 | 0.005 | 0.52 | 0.51 | 0.003 | 0.14 | 0.002 | 3.73E-08 | No | 2.68E-05 | 30.31 |
| 64 | 2 | rs13010566 | N/A | A | -0.01 | 0.002 | 0.44 | 0.47 | 0.003 | 0.49 | 0.002 | 4.59E-10 | No | 3.43E-05 | 38.88 |
| 65 | 2 | rs13029509 | N/A | A | -0.01 | -0.003 | 0.47 | 0.48 | 0.003 | 0.34 | 0.002 | 7.17E-10 | No | 3.36E-05 | 38.08 |
| 66 | 3 | rs13090388 | N/A | C | -0.03 | 0.000 | 0.69 | 0.69 | 0.004 | 0.97 | 0.002 | 4.29E-54 | No | 0.000212 | 240.25 |
| 67 | 4 | rs13130765 | N/A | C | -0.01 | 0.007 | 0.46 | 0.48 | 0.004 | 0.06 | 0.002 | 4.69E-09 | Yes | 3.04E-05 | 34.35 |
| 68 | 4 | rs13141210 | N/A | C | -0.01 | -0.002 | 0.49 | 0.49 | 0.003 | 0.62 | 0.002 | 2.26E-15 | No | 5.53E-05 | 62.61 |
| 69 | 4 | rs13145650 | N/A | C | 0.02 | 0.001 | 0.09 | 0.08 | 0.006 | 0.85 | 0.003 | 3.80E-10 | No | 3.47E-05 | 39.29 |
| 70 | 13 | rs1334297 | N/A | A | 0.02 | 0.002 | 0.78 | 0.74 | 0.004 | 0.54 | 0.002 | 3.06E-37 | No | 0.000144 | 162.70 |
| 71 | 2 | rs13422673 | N/A | C | 0.01 | -0.004 | 0.52 | 0.53 | 0.003 | 0.24 | 0.002 | 1.74E-12 | No | 4.41E-05 | 49.91 |
| 72 | 2 | rs13428598 | N/A | T | 0.02 | -0.005 | 0.38 | 0.39 | 0.004 | 0.13 | 0.002 | 3.90E-21 | No | 7.84E-05 | 88.79 |
| 73 | 5 | rs1363862 | N/A | G | 0.01 | -0.001 | 0.74 | 0.73 | 0.004 | 0.81 | 0.002 | 1.02E-09 | No | 3.29E-05 | 37.20 |
| 74 | 17 | rs1381247 | N/A | C | -0.01 | -0.005 | 0.29 | 0.33 | 0.004 | 0.18 | 0.002 | 2.46E-08 | No | 2.74E-05 | 30.98 |
| 75 | 4 | rs1391438 | N/A | T | 0.02 | 0.004 | 0.31 | 0.33 | 0.004 | 0.33 | 0.002 | 5.79E-20 | No | 7.36E-05 | 83.28 |
| 76 | 2 | rs1427298 | N/A | C | -0.01 | 0.004 | 0.59 | 0.59 | 0.004 | 0.22 | 0.002 | 3.28E-09 | No | 3.11E-05 | 35.17 |
| 77 | 4 | rs1450782 | N/A | G | -0.01 | -0.002 | 0.60 | 0.58 | 0.003 | 0.65 | 0.002 | 4.93E-08 | No | 2.64E-05 | 29.84 |
| 78 | 2 | rs1455350 | N/A | T | 0.02 | -0.002 | 0.53 | 0.51 | 0.003 | 0.57 | 0.002 | 2.61E-21 | Yes | 7.96E-05 | 90.14 |
| 79 | 5 | rs152603 | N/A | G | 0.01 | -0.005 | 0.39 | 0.36 | 0.004 | 0.17 | 0.002 | 9.47E-09 | No | 2.93E-05 | 33.14 |
| 80 | 12 | rs1558727 | N/A | C | 0.01 | -0.001 | 0.52 | 0.52 | 0.003 | 0.81 | 0.002 | 3.09E-10 | No | 3.49E-05 | 39.54 |
| 81 | 8 | rs1566085 | N/A | T | 0.02 | -0.0003 | 0.57 | 0.55 | 0.003 | 0.92 | 0.002 | 6.90E-22 | No | 8.18E-05 | 92.54 |
| 82 | 1 | rs1569092 | N/A | A | 0.02 | -0.002 | 0.18 | 0.14 | 0.005 | 0.72 | 0.002 | 1.16E-14 | No | 5.27E-05 | 59.63 |
| 83 | 13 | rs1582173 | N/A | C | -0.01 | -0.005 | 0.26 | 0.25 | 0.004 | 0.22 | 0.002 | 1.33E-09 | No | 3.25E-05 | 36.80 |
| 84 | 5 | rs1584469 | N/A | T | -0.01 | -0.001 | 0.31 | 0.30 | 0.004 | 0.85 | 0.002 | 2.10E-12 | No | 4.38E-05 | 49.61 |
| 85 | 5 | rs1592757 | N/A | C | -0.01 | 0.000 | 0.38 | 0.36 | 0.004 | 0.96 | 0.002 | 5.89E-09 | No | 2.98E-05 | 33.70 |
| 86 | 4 | rs1595973 | N/A | C | 0.01 | -0.001 | 0.44 | 0.42 | 0.003 | 0.83 | 0.002 | 6.56E-09 | No | 2.96E-05 | 33.55 |
| 87 | 18 | rs1618725 | N/A | T | 0.01 | 0.002 | 0.52 | 0.47 | 0.003 | 0.62 | 0.002 | 2.22E-17 | No | 6.37E-05 | 72.05 |

| | | | | | | | | | | | | | | | |
|---|---|---|---|---|---|---|---|---|---|---|---|---|---|---|---|
| 88 | 1 | rs1620977 | N/A | G | -0.02 | 0.005 | 0.69 | 0.73 | 0.004 | 0.21 | 0.002 | 1.14E-25 | No | 9.73E-05 | 110.09 |
| 89 | 12 | rs1671770 | N/A | C | -0.01 | -0.005 | 0.81 | 0.82 | 0.004 | 0.27 | 0.002 | 1.91E-09 | No | 3.20E-05 | 36.22 |
| 90 | 2 | rs16846463 | N/A | A | 0.02 | 0.004 | 0.88 | 0.90 | 0.006 | 0.48 | 0.003 | 1.38E-15 | No | 5.61E-05 | 63.55 |
| 91 | 1 | rs16854920 | N/A | T | -0.01 | 0.005 | 0.64 | 0.66 | 0.004 | 0.16 | 0.002 | 2.51E-08 | No | 2.73E-05 | 30.95 |
| 92 | 12 | rs1689510 | N/A | G | -0.02 | -0.001 | 0.66 | 0.67 | 0.004 | 0.76 | 0.002 | 1.40E-22 | No | 8.46E-05 | 95.71 |
| 93 | 20 | rs16995054 | N/A | C | 0.01 | 0.012 | 0.80 | 0.80 | 0.004 | 0.01 | 0.002 | 2.52E-11 | No | 3.95E-05 | 44.66 |
| 94 | 3 | rs17048855 | N/A | A | 0.01 | 0.005 | 0.32 | 0.33 | 0.004 | 0.14 | 0.002 | 3.27E-11 | No | 3.87E-05 | 43.75 |
| 95 | 12 | rs17110109 | N/A | C | 0.01 | -0.003 | 0.38 | 0.38 | 0.004 | 0.35 | 0.002 | 4.71E-09 | No | 3.02E-05 | 34.17 |
| 96 | 10 | rs17126938 | N/A | T | -0.02 | -0.008 | 0.88 | 0.86 | 0.005 | 0.08 | 0.003 | 8.14E-10 | No | 3.33E-05 | 37.75 |
| 97 | 9 | rs17425572 | N/A | G | -0.01 | 0.001 | 0.54 | 0.55 | 0.003 | 0.73 | 0.002 | 6.89E-13 | No | 4.58E-05 | 51.84 |
| 98 | 5 | rs17489649 | N/A | A | 0.01 | -0.001 | 0.67 | 0.67 | 0.004 | 0.78 | 0.002 | 1.57E-14 | No | 5.21E-05 | 58.98 |
| 99 | 20 | rs175325 | N/A | A | -0.01 | 0.001 | 0.58 | 0.60 | 0.004 | 0.73 | 0.002 | 1.11E-11 | No | 4.06E-05 | 45.91 |
| 100 | 7 | rs17551064 | N/A | G | -0.01 | 0.001 | 0.16 | 0.17 | 0.005 | 0.90 | 0.002 | 8.62E-11 | No | 3.72E-05 | 42.14 |
| 101 | 5 | rs17563464 | N/A | A | -0.01 | 0.001 | 0.20 | 0.22 | 0.004 | 0.75 | 0.002 | 2.89E-12 | No | 4.29E-05 | 48.54 |
| 102 | 11 | rs17565975 | N/A | A | -0.01 | -0.003 | 0.53 | 0.55 | 0.003 | 0.32 | 0.002 | 2.56E-11 | No | 3.94E-05 | 44.60 |
| 103 | 4 | rs17598675 | N/A | C | 0.01 | 0.000 | 0.52 | 0.48 | 0.003 | 0.98 | 0.002 | 1.75E-12 | No | 4.39E-05 | 49.74 |
| 104 | 14 | rs176218 | N/A | G | -0.02 | 0.010 | 0.80 | 0.81 | 0.004 | 0.03 | 0.002 | 1.85E-18 | No | 6.78E-05 | 76.70 |
| 105 | 5 | rs1827540 | N/A | G | -0.01 | 0.008 | 0.47 | 0.48 | 0.003 | 0.02 | 0.002 | 4.50E-10 | No | 3.43E-05 | 38.88 |
| 106 | 8 | rs1866823 | N/A | A | 0.01 | -0.002 | 0.55 | 0.55 | 0.003 | 0.53 | 0.002 | 3.81E-09 | No | 3.08E-05 | 34.82 |
| 107 | 2 | rs1882273 | N/A | G | 0.01 | -0.002 | 0.65 | 0.65 | 0.004 | 0.65 | 0.002 | 1.09E-11 | No | 4.09E-05 | 46.25 |
| 108 | 19 | rs192436652 | N/A | C | 0.03 | -0.007 | 0.98 | 0.97 | 0.012 | 0.57 | 0.005 | 1.35E-10 | No | 3.64E-05 | 41.17 |
| 109 | 10 | rs1925576 | N/A | A | -0.01 | -0.004 | 0.56 | 0.54 | 0.003 | 0.20 | 0.002 | 4.94E-09 | No | 3.00E-05 | 33.99 |
| 110 | 2 | rs1947114 | N/A | G | 0.01 | -0.001 | 0.26 | 0.25 | 0.004 | 0.71 | 0.002 | 2.64E-08 | No | 2.75E-05 | 31.12 |
| 111 | 21 | rs1964927 | N/A | G | -0.01 | 0.006 | 0.64 | 0.64 | 0.004 | 0.11 | 0.002 | 9.90E-16 | No | 5.71E-05 | 64.63 |
| 112 | 14 | rs2016392 | N/A | A | -0.01 | 0.004 | 0.23 | 0.21 | 0.004 | 0.30 | 0.002 | 4.35E-09 | No | 3.04E-05 | 34.45 |
| 113 | 16 | rs2052285 | N/A | A | 0.01 | -0.003 | 0.58 | 0.59 | 0.004 | 0.46 | 0.002 | 1.34E-10 | No | 3.64E-05 | 41.18 |
| 114 | 14 | rs2067854 | N/A | A | 0.01 | -0.005 | 0.18 | 0.20 | 0.004 | 0.25 | 0.002 | 1.38E-12 | No | 4.41E-05 | 49.94 |
| 115 | 6 | rs2179152 | N/A | C | 0.01 | 0.007 | 0.64 | 0.63 | 0.004 | 0.04 | 0.002 | 1.21E-16 | No | 6.04E-05 | 68.34 |
| 116 | 6 | rs2182505 | N/A | T | 0.01 | 0.003 | 0.26 | 0.28 | 0.004 | 0.51 | 0.002 | 1.64E-08 | No | 2.83E-05 | 31.99 |
| 117 | 17 | rs225291 | N/A | G | -0.01 | 0.002 | 0.80 | 0.79 | 0.004 | 0.64 | 0.002 | 1.84E-08 | No | 2.80E-05 | 31.71 |
| 118 | 6 | rs2256965 | N/A | G | -0.01 | -0.003 | 0.54 | 0.58 | 0.003 | 0.36 | 0.002 | 1.59E-10 | No | 3.63E-05 | 41.08 |
| 119 | 7 | rs2283076 | N/A | A | 0.01 | 0.005 | 0.79 | 0.78 | 0.004 | 0.24 | 0.002 | 2.07E-08 | No | 2.77E-05 | 31.39 |

| | | | | | | | | | | | | | | | |
|---|---|---|---|---|---|---|---|---|---|---|---|---|---|---|---|
| 120 | 19 | rs2287838 | N/A | A | -0.01 | 0.004 | 0.53 | 0.54 | 0.003 | 0.28 | 0.002 | 1.53E-11 | No | 4.01E-05 | 45.38 |
| 121 | 17 | rs2302761 | N/A | T | 0.01 | -0.0003 | 0.19 | 0.21 | 0.004 | 0.94 | 0.002 | 1.00E-10 | No | 3.71E-05 | 41.97 |
| 122 | 4 | rs2347526 | N/A | T | -0.01 | -0.00002 | 0.36 | 0.34 | 0.004 | 1.00 | 0.002 | 6.84E-15 | No | 5.37E-05 | 60.74 |
| 123 | 15 | rs2414072 | N/A | A | -0.01 | -0.004 | 0.49 | 0.46 | 0.003 | 0.21 | 0.002 | 4.35E-09 | Yes | 3.05E-05 | 34.54 |
| 124 | 14 | rs242093 | N/A | A | -0.01 | 0.001 | 0.55 | 0.58 | 0.003 | 0.83 | 0.002 | 2.07E-09 | No | 3.17E-05 | 35.93 |
| 125 | 8 | rs2447535 | N/A | A | -0.01 | -0.001 | 0.28 | 0.29 | 0.004 | 0.88 | 0.002 | 1.69E-10 | No | 3.60E-05 | 40.75 |
| 126 | 13 | rs2478208 | N/A | C | -0.01 | -0.001 | 0.50 | 0.52 | 0.003 | 0.67 | 0.002 | 4.82E-10 | Yes | 3.43E-05 | 38.88 |
| 127 | 5 | rs2545798 | N/A | T | 0.01 | 0.002 | 0.51 | 0.52 | 0.003 | 0.66 | 0.002 | 3.11E-15 | Yes | 5.47E-05 | 61.96 |
| 128 | 18 | rs2554835 | N/A | G | -0.01 | 0.003 | 0.60 | 0.61 | 0.004 | 0.38 | 0.002 | 2.69E-08 | No | 2.74E-05 | 30.98 |
| 129 | 2 | rs2570497 | N/A | C | 0.01 | 0.006 | 0.33 | 0.36 | 0.004 | 0.11 | 0.002 | 3.03E-12 | No | 4.29E-05 | 48.53 |
| 130 | 8 | rs2725370 | N/A | T | -0.02 | -0.004 | 0.29 | 0.30 | 0.004 | 0.32 | 0.002 | 1.97E-16 | No | 5.96E-05 | 67.47 |
| 131 | 13 | rs277828 | N/A | C | 0.01 | 0.000 | 0.74 | 0.75 | 0.004 | 0.99 | 0.002 | 2.71E-08 | No | 2.74E-05 | 30.98 |
| 132 | 1 | rs2819336 | N/A | T | 0.02 | -0.001 | 0.34 | 0.37 | 0.004 | 0.82 | 0.002 | 5.46E-25 | No | 9.42E-05 | 106.66 |
| 133 | 1 | rs2820314 | rs2820312 | G | 0.01 | -0.001 | 0.69 | 0.68 | 0.004 | 0.83 | 0.002 | 1.26E-09 | No | 3.25E-05 | 36.74 |
| 134 | 4 | rs28373063 | N/A | G | -0.01 | 0.002 | 0.79 | 0.82 | 0.005 | 0.70 | 0.002 | 1.23E-09 | No | 3.25E-05 | 36.79 |
| 135 | 15 | rs28513670 | N/A | G | 0.01 | -0.002 | 0.15 | 0.17 | 0.005 | 0.72 | 0.002 | 5.06E-11 | No | 3.81E-05 | 43.09 |
| 136 | 3 | rs2885198 | N/A | A | 0.01 | -0.001 | 0.50 | 0.52 | 0.003 | 0.86 | 0.002 | 1.81E-09 | No | 3.21E-05 | 36.35 |
| 137 | 1 | rs2901616 | N/A | A | 0.01 | 0.003 | 0.52 | 0.48 | 0.003 | 0.36 | 0.002 | 3.77E-08 | No | 2.68E-05 | 30.28 |
| 138 | 19 | rs2905426 | N/A | G | -0.01 | -0.007 | 0.36 | 0.35 | 0.004 | 0.06 | 0.002 | 9.26E-09 | No | 2.90E-05 | 32.82 |
| 139 | 8 | rs2923431 | N/A | C | 0.01 | 0.006 | 0.64 | 0.63 | 0.004 | 0.07 | 0.002 | 9.84E-11 | No | 3.71E-05 | 41.95 |
| 140 | 7 | rs2971970 | N/A | T | -0.02 | 0.014 | 0.23 | 0.22 | 0.004 | 0.00 | 0.002 | 1.25E-15 | No | 5.64E-05 | 63.85 |
| 141 | 14 | rs2998315 | N/A | G | 0.01 | 0.000 | 0.58 | 0.54 | 0.003 | 0.98 | 0.002 | 1.12E-13 | No | 4.87E-05 | 55.07 |
| 142 | 10 | rs3013014 | N/A | G | 0.01 | 0.001 | 0.39 | 0.42 | 0.003 | 0.72 | 0.002 | 2.92E-09 | No | 3.13E-05 | 35.44 |
| 143 | 1 | rs301800 | N/A | T | 0.02 | 0.007 | 0.18 | 0.19 | 0.004 | 0.13 | 0.002 | 1.33E-11 | No | 4.05E-05 | 45.80 |
| 144 | 1 | rs3026996 | N/A | A | 0.02 | 0.008 | 0.72 | 0.76 | 0.004 | 0.06 | 0.002 | 1.05E-14 | No | 5.27E-05 | 59.65 |
| 145 | 5 | rs31940 | N/A | G | -0.02 | -0.005 | 0.87 | 0.87 | 0.005 | 0.36 | 0.002 | 3.24E-10 | No | 3.50E-05 | 39.60 |
| 146 | 7 | rs320693 | N/A | C | 0.01 | -0.002 | 0.47 | 0.46 | 0.003 | 0.62 | 0.002 | 1.58E-12 | Yes | 4.43E-05 | 50.16 |
| 147 | 4 | rs337637 | N/A | G | -0.01 | 0.003 | 0.66 | 0.65 | 0.004 | 0.42 | 0.002 | 2.11E-10 | No | 3.56E-05 | 40.25 |
| 148 | 5 | rs34316 | N/A | C | -0.02 | 0.005 | 0.58 | 0.58 | 0.003 | 0.18 | 0.002 | 3.35E-30 | No | 0.0001 | 129.73 |
| 149 | 1 | rs34394051 | N/A | G | 0.01 | -0.008 | 0.16 | 0.16 | 0.005 | 0.09 | 0.002 | 6.20E-09 | No | 2.97E-05 | 33.64 |
| 150 | 16 | rs34485537 | N/A | T | 0.01 | -0.005 | 0.39 | 0.40 | 0.004 | 0.18 | 0.002 | 5.67E-10 | No | 3.41E-05 | 38.61 |
| 151 | 7 | rs34853711 | N/A | C | -0.02 | -0.007 | 0.25 | 0.21 | 0.004 | 0.10 | 0.002 | 9.88E-15 | No | 5.30E-05 | 60.02 |

| | | | | | | | | | | | | | | | |
|---|---|---|---|---|---|---|---|---|---|---|---|---|---|---|---|
| 152 | 1 | rs35039375 | N/A | G | -0.02 | 0.006 | 0.09 | 0.09 | 0.006 | 0.28 | 0.003 | 1.22E-11 | No | 4.05E-05 | 45.80 |
| 153 | 12 | rs35309068 | N/A | G | 0.01 | -0.002 | 0.47 | 0.44 | 0.003 | 0.63 | 0.002 | 1.15E-14 | No | 5.27E-05 | 59.68 |
| 154 | 16 | rs35316276 | N/A | C | -0.01 | -0.010 | 0.71 | 0.71 | 0.004 | 0.01 | 0.002 | 1.52E-09 | No | 3.23E-05 | 36.56 |
| 155 | 7 | rs35417702 | N/A | C | 0.01 | 0.003 | 0.42 | 0.47 | 0.003 | 0.41 | 0.002 | 1.93E-17 | No | 6.38E-05 | 72.25 |
| 156 | 3 | rs35475880 | N/A | G | 0.02 | -0.001 | 0.82 | 0.81 | 0.004 | 0.74 | 0.002 | 3.80E-13 | No | 4.66E-05 | 52.77 |
| 157 | 22 | rs35532491 | N/A | T | 0.02 | 0.005 | 0.11 | 0.10 | 0.006 | 0.42 | 0.003 | 2.42E-12 | No | 4.35E-05 | 49.24 |
| 158 | 4 | rs36083520 | N/A | C | 0.02 | 0.000 | 0.17 | 0.17 | 0.005 | 0.94 | 0.002 | 2.60E-13 | No | 4.71E-05 | 53.36 |
| 159 | 7 | rs36119825 | N/A | G | -0.01 | -0.001 | 0.53 | 0.56 | 0.003 | 0.78 | 0.002 | 4.82E-10 | No | 3.41E-05 | 38.64 |
| 160 | 4 | rs363096 | N/A | C | 0.01 | -0.004 | 0.57 | 0.58 | 0.003 | 0.23 | 0.002 | 2.04E-15 | No | 5.55E-05 | 62.80 |
| 161 | 1 | rs3747631 | N/A | G | -0.02 | 0.001 | 0.77 | 0.79 | 0.004 | 0.78 | 0.002 | 2.97E-26 | No | 9.95E-05 | 112.58 |
| 162 | 22 | rs3788556 | N/A | T | 0.01 | -0.004 | 0.46 | 0.49 | 0.004 | 0.29 | 0.002 | 2.78E-11 | No | 3.91E-05 | 44.29 |
| 163 | 6 | rs3800546 | N/A | G | -0.01 | -0.001 | 0.27 | 0.26 | 0.004 | 0.74 | 0.002 | 9.73E-10 | No | 3.29E-05 | 37.18 |
| 164 | 16 | rs3809634 | N/A | A | -0.01 | 0.001 | 0.67 | 0.68 | 0.004 | 0.89 | 0.002 | 1.09E-08 | No | 2.89E-05 | 32.71 |
| 165 | 5 | rs3890802 | N/A | A | -0.01 | -0.004 | 0.27 | 0.28 | 0.004 | 0.28 | 0.002 | 2.74E-09 | No | 3.11E-05 | 35.19 |
| 166 | 1 | rs3897821 | N/A | A | 0.02 | -0.006 | 0.65 | 0.66 | 0.004 | 0.10 | 0.002 | 8.25E-17 | No | 6.15E-05 | 69.63 |
| 167 | 12 | rs401687 | N/A | G | -0.01 | 0.002 | 0.54 | 0.54 | 0.003 | 0.47 | 0.002 | 1.86E-11 | Yes | 4.00E-05 | 45.28 |
| 168 | 5 | rs406413 | N/A | A | 0.02 | -0.006 | 0.79 | 0.78 | 0.004 | 0.17 | 0.002 | 4.84E-16 | No | 5.81E-05 | 65.77 |
| 169 | 7 | rs4073894 | N/A | A | 0.02 | -0.004 | 0.18 | 0.21 | 0.004 | 0.38 | 0.002 | 5.40E-13 | No | 4.61E-05 | 52.17 |
| 170 | 3 | rs4328757 | N/A | T | 0.01 | -0.005 | 0.65 | 0.61 | 0.004 | 0.16 | 0.002 | 9.39E-10 | No | 3.32E-05 | 37.60 |
| 171 | 6 | rs4352658 | N/A | T | -0.02 | -0.013 | 0.09 | 0.08 | 0.006 | 0.04 | 0.003 | 5.55E-12 | No | 4.19E-05 | 47.38 |
| 172 | 20 | rs4369924 | N/A | A | 0.01 | -0.001 | 0.17 | 0.16 | 0.005 | 0.82 | 0.002 | 5.82E-09 | No | 2.99E-05 | 33.88 |
| 173 | 9 | rs4382592 | N/A | T | -0.02 | -0.004 | 0.30 | 0.31 | 0.004 | 0.34 | 0.002 | 1.01E-18 | No | 6.91E-05 | 78.20 |
| 174 | 10 | rs4384309 | N/A | G | -0.01 | 0.001 | 0.52 | 0.53 | 0.004 | 0.77 | 0.002 | 2.52E-10 | No | 3.55E-05 | 40.16 |
| 175 | 14 | rs4442732 | N/A | G | -0.01 | 0.006 | 0.60 | 0.60 | 0.004 | 0.11 | 0.002 | 1.49E-09 | No | 3.22E-05 | 36.48 |
| 176 | 5 | rs466047 | N/A | A | 0.01 | -0.005 | 0.50 | 0.48 | 0.003 | 0.13 | 0.002 | 1.80E-10 | Yes | 3.60E-05 | 40.73 |
| 177 | 2 | rs4667029 | N/A | G | 0.01 | -0.003 | 0.39 | 0.40 | 0.004 | 0.43 | 0.002 | 3.44E-08 | No | 2.69E-05 | 30.50 |
| 178 | 5 | rs4700393 | N/A | G | 0.02 | -0.006 | 0.53 | 0.53 | 0.003 | 0.10 | 0.002 | 1.51E-34 | No | 0.0001 | 150.57 |
| 179 | 7 | rs4726070 | N/A | G | -0.01 | -0.002 | 0.38 | 0.40 | 0.004 | 0.50 | 0.002 | 5.95E-13 | No | 4.57E-05 | 51.69 |
| 180 | 8 | rs4733264 | N/A | C | -0.01 | 0.005 | 0.63 | 0.61 | 0.004 | 0.14 | 0.002 | 4.53E-08 | No | 2.66E-05 | 30.06 |
| 181 | 11 | rs4757957 | N/A | C | 0.01 | 0.0001 | 0.65 | 0.69 | 0.004 | 0.98 | 0.002 | 1.81E-14 | No | 5.19E-05 | 58.72 |
| 182 | 12 | rs4766424 | N/A | G | -0.01 | 0.003 | 0.91 | 0.87 | 0.005 | 0.52 | 0.003 | 4.43E-08 | No | 2.64E-05 | 29.87 |
| 183 | 15 | rs4778058 | N/A | C | 0.01 | -0.002 | 0.52 | 0.51 | 0.003 | 0.55 | 0.002 | 2.40E-09 | No | 3.16E-05 | 35.79 |

| | | | | | | | | | | | | | | | |
|---|---|---|---|---|---|---|---|---|---|---|---|---|---|---|---|
| 184 | 16 | rs4787457 | N/A | G | -0.02 | 0.002 | 0.31 | 0.35 | 0.004 | 0.49 | 0.002 | 3.73E-23 | No | 8.64E-05 | 97.85 |
| 185 | 20 | rs4810227 | N/A | A | 0.01 | -0.0003 | 0.63 | 0.61 | 0.004 | 0.93 | 0.002 | 3.57E-13 | No | 4.67E-05 | 52.83 |
| 186 | 1 | rs4839155 | N/A | T | 0.01 | 0.003 | 0.75 | 0.77 | 0.004 | 0.52 | 0.002 | 3.94E-10 | No | 3.46E-05 | 39.12 |
| 187 | 1 | rs4846724 | N/A | G | -0.01 | 0.000 | 0.51 | 0.46 | 0.003 | 0.94 | 0.002 | 2.26E-09 | No | 3.17E-05 | 35.86 |
| 188 | 6 | rs4870482 | N/A | G | -0.01 | 0.007 | 0.26 | 0.28 | 0.004 | 0.06 | 0.002 | 1.27E-08 | No | 2.87E-05 | 32.49 |
| 189 | 16 | rs4888746 | N/A | G | -0.01 | -0.003 | 0.38 | 0.39 | 0.004 | 0.34 | 0.002 | 4.15E-08 | No | 2.64E-05 | 29.93 |
| 190 | 6 | rs4895650 | N/A | T | 0.01 | -0.005 | 0.54 | 0.53 | 0.003 | 0.12 | 0.002 | 2.16E-08 | No | 2.78E-05 | 31.48 |
| 191 | 11 | rs4945424 | N/A | A | -0.01 | 0.000 | 0.42 | 0.44 | 0.003 | 0.90 | 0.002 | 6.94E-09 | No | 2.97E-05 | 33.65 |
| 192 | 12 | rs4964046 | N/A | G | 0.01 | 0.002 | 0.34 | 0.36 | 0.004 | 0.65 | 0.002 | 3.36E-09 | No | 3.09E-05 | 35.00 |
| 193 | 2 | rs4972400 | N/A | G | -0.01 | 0.000 | 0.65 | 0.65 | 0.004 | 0.93 | 0.002 | 1.70E-10 | No | 3.60E-05 | 40.79 |
| 194 | 15 | rs4984541 | N/A | A | -0.01 | 0.004 | 0.76 | 0.76 | 0.004 | 0.39 | 0.002 | 2.77E-09 | No | 3.13E-05 | 35.48 |
| 195 | 11 | rs510706 | N/A | G | -0.01 | -0.001 | 0.38 | 0.37 | 0.004 | 0.84 | 0.002 | 2.73E-09 | No | 3.13E-05 | 35.40 |
| 196 | 1 | rs532799 | N/A | G | 0.01 | -0.003 | 0.67 | 0.69 | 0.004 | 0.45 | 0.002 | 1.08E-08 | No | 2.87E-05 | 32.51 |
| 197 | 7 | rs535307 | N/A | A | 0.01 | -0.0002 | 0.32 | 0.32 | 0.004 | 0.95 | 0.002 | 4.73E-08 | No | 2.63E-05 | 29.77 |
| 198 | 3 | rs55736314 | N/A | C | -0.01 | 0.001 | 0.58 | 0.59 | 0.004 | 0.73 | 0.002 | 1.63E-16 | No | 5.98E-05 | 67.64 |
| 199 | 10 | rs55771711 | N/A | C | 0.02 | -0.009 | 0.23 | 0.25 | 0.004 | 0.02 | 0.002 | 5.41E-15 | No | 5.39E-05 | 61.06 |
| 200 | 15 | rs56391344 | N/A | A | 0.02 | 0.003 | 0.24 | 0.25 | 0.004 | 0.40 | 0.002 | 1.34E-15 | No | 5.62E-05 | 63.59 |
| 201 | 1 | rs575113 | N/A | A | 0.01 | -0.003 | 0.28 | 0.29 | 0.004 | 0.37 | 0.002 | 5.30E-12 | No | 4.22E-05 | 47.73 |
| 202 | 1 | rs59123361 | N/A | A | -0.02 | 0.004 | 0.11 | 0.10 | 0.006 | 0.55 | 0.003 | 5.87E-13 | No | 4.57E-05 | 51.78 |
| 203 | 8 | rs59480703 | N/A | G | 0.01 | -0.002 | 0.84 | 0.80 | 0.004 | 0.68 | 0.002 | 8.32E-09 | No | 2.92E-05 | 33.10 |
| 204 | 16 | rs60483752 | N/A | G | -0.01 | 0.010 | 0.42 | 0.44 | 0.004 | 0.00 | 0.002 | 3.89E-10 | Yes | 3.47E-05 | 39.28 |
| 205 | 14 | rs60904894 | N/A | A | -0.01 | 0.002 | 0.50 | 0.49 | 0.003 | 0.51 | 0.002 | 1.62E-09 | No | 3.21E-05 | 36.35 |
| 206 | 20 | rs6122735 | N/A | T | 0.01 | 0.000 | 0.41 | 0.39 | 0.004 | 0.94 | 0.002 | 1.49E-09 | No | 3.22E-05 | 36.41 |
| 207 | 20 | rs6123924 | N/A | A | 0.02 | -0.005 | 0.84 | 0.84 | 0.005 | 0.34 | 0.002 | 7.55E-11 | No | 3.74E-05 | 42.28 |
| 208 | 18 | rs613872 | N/A | G | 0.02 | 0.003 | 0.17 | 0.18 | 0.004 | 0.51 | 0.002 | 1.20E-14 | No | 5.25E-05 | 59.43 |
| 209 | 18 | rs62097985 | N/A | C | 0.01 | 0.004 | 0.59 | 0.58 | 0.003 | 0.23 | 0.002 | 6.06E-14 | No | 4.95E-05 | 56.08 |
| 210 | 18 | rs62103084 | N/A | T | 0.02 | 0.006 | 0.14 | 0.11 | 0.006 | 0.28 | 0.003 | 1.42E-09 | No | 3.23E-05 | 36.56 |
| 211 | 2 | rs62157915 | N/A | T | -0.02 | 0.007 | 0.94 | 0.94 | 0.007 | 0.32 | 0.003 | 1.96E-09 | No | 3.19E-05 | 36.10 |
| 212 | 2 | rs62183776 | N/A | T | -0.01 | 0.001 | 0.19 | 0.19 | 0.004 | 0.89 | 0.002 | 1.65E-09 | No | 3.21E-05 | 36.33 |
| 213 | 2 | rs62184480 | N/A | T | -0.02 | -0.002 | 0.24 | 0.27 | 0.004 | 0.63 | 0.002 | 1.28E-15 | No | 5.65E-05 | 64.00 |
| 214 | 1 | rs622169 | N/A | C | -0.01 | -0.002 | 0.53 | 0.55 | 0.004 | 0.57 | 0.002 | 1.89E-08 | No | 2.78E-05 | 31.50 |
| 215 | 7 | rs62439690 | N/A | G | 0.01 | -0.008 | 0.73 | 0.74 | 0.004 | 0.04 | 0.002 | 2.18E-08 | No | 2.77E-05 | 31.39 |

| | | | | | | | | | | | | | | | |
|---|---|---|---|---|---|---|---|---|---|---|---|---|---|---|---|
| 216 | 7 | rs62444881 | N/A | C | -0.02 | -0.010 | 0.81 | 0.80 | 0.004 | 0.02 | 0.002 | 5.79E-17 | No | 6.18E-05 | 69.96 |
| 217 | 15 | rs6493265 | N/A | C | 0.01 | -0.006 | 0.61 | 0.60 | 0.003 | 0.06 | 0.002 | 1.70E-15 | No | 5.60E-05 | 63.36 |
| 218 | 20 | rs6513959 | N/A | A | 0.01 | -0.001 | 0.72 | 0.69 | 0.004 | 0.89 | 0.002 | 1.88E-10 | No | 3.58E-05 | 40.48 |
| 219 | 6 | rs6557171 | N/A | C | 0.02 | 0.002 | 0.72 | 0.68 | 0.004 | 0.59 | 0.002 | 4.15E-18 | No | 6.62E-05 | 74.95 |
| 220 | 1 | rs663234 | N/A | G | -0.01 | 0.001 | 0.61 | 0.60 | 0.004 | 0.76 | 0.002 | 7.39E-09 | No | 2.95E-05 | 33.36 |
| 221 | 3 | rs66568921 | N/A | G | 0.02 | -0.004 | 0.36 | 0.35 | 0.004 | 0.32 | 0.002 | 7.49E-18 | No | 6.53E-05 | 73.94 |
| 222 | 2 | rs6731373 | N/A | G | 0.01 | 0.005 | 0.66 | 0.66 | 0.004 | 0.22 | 0.002 | 3.47E-12 | No | 4.25E-05 | 48.15 |
| 223 | 2 | rs6731967 | N/A | C | -0.01 | 0.002 | 0.21 | 0.24 | 0.004 | 0.65 | 0.002 | 2.36E-09 | No | 3.14E-05 | 35.52 |
| 224 | 8 | rs67885444 | N/A | T | 0.01 | -0.004 | 0.17 | 0.15 | 0.005 | 0.40 | 0.002 | 1.48E-09 | No | 3.24E-05 | 36.73 |
| 225 | 2 | rs67890737 | N/A | C | 0.01 | 0.000 | 0.67 | 0.65 | 0.004 | 0.95 | 0.002 | 2.01E-10 | No | 3.59E-05 | 40.63 |
| 226 | 3 | rs6803651 | N/A | G | -0.01 | -0.003 | 0.59 | 0.57 | 0.004 | 0.43 | 0.002 | 4.36E-11 | No | 3.82E-05 | 43.24 |
| 227 | 3 | rs6805241 | N/A | C | -0.01 | -0.001 | 0.20 | 0.23 | 0.004 | 0.81 | 0.002 | 3.09E-12 | No | 4.28E-05 | 48.45 |
| 228 | 5 | rs6867851 | N/A | C | -0.01 | 0.007 | 0.42 | 0.41 | 0.004 | 0.06 | 0.002 | 3.97E-12 | Yes | 4.25E-05 | 48.11 |
| 229 | 6 | rs6938002 | N/A | G | 0.01 | 0.003 | 0.60 | 0.60 | 0.004 | 0.42 | 0.002 | 5.41E-09 | No | 3.00E-05 | 33.95 |
| 230 | 7 | rs6959891 | N/A | G | -0.01 | -0.001 | 0.30 | 0.29 | 0.004 | 0.81 | 0.002 | 1.74E-09 | No | 3.19E-05 | 36.13 |
| 231 | 8 | rs7012546 | N/A | C | -0.01 | -0.001 | 0.58 | 0.59 | 0.003 | 0.73 | 0.002 | 4.93E-09 | No | 3.04E-05 | 34.41 |
| 232 | 8 | rs7016302 | N/A | G | 0.01 | -0.004 | 0.18 | 0.17 | 0.005 | 0.41 | 0.002 | 4.98E-08 | No | 2.63E-05 | 29.72 |
| 233 | 5 | rs702606 | N/A | T | 0.01 | -0.003 | 0.83 | 0.86 | 0.005 | 0.56 | 0.003 | 1.12E-08 | No | 2.88E-05 | 32.58 |
| 234 | 9 | rs7029718 | N/A | G | -0.02 | 0.002 | 0.56 | 0.60 | 0.004 | 0.50 | 0.002 | 1.85E-44 | No | 0.0002 | 196.48 |
| 235 | 9 | rs7031698 | N/A | C | 0.01 | -0.004 | 0.78 | 0.77 | 0.004 | 0.31 | 0.002 | 1.26E-09 | No | 3.24E-05 | 36.70 |
| 236 | 12 | rs710629 | N/A | A | 0.01 | 0.005 | 0.66 | 0.65 | 0.004 | 0.17 | 0.002 | 2.96E-09 | No | 3.13E-05 | 35.39 |
| 237 | 1 | rs71646142 | N/A | C | -0.01 | -0.002 | 0.83 | 0.81 | 0.004 | 0.58 | 0.002 | 3.11E-09 | No | 3.10E-05 | 35.12 |
| 238 | 18 | rs7233920 | N/A | A | -0.01 | 0.001 | 0.22 | 0.23 | 0.004 | 0.90 | 0.002 | 7.13E-11 | No | 3.74E-05 | 42.38 |
| 239 | 11 | rs72486027 | N/A | C | 0.01 | 0.003 | 0.72 | 0.75 | 0.004 | 0.41 | 0.002 | 1.25E-08 | No | 2.87E-05 | 32.50 |
| 240 | 19 | rs7257460 | N/A | T | 0.01 | 0.000 | 0.73 | 0.70 | 0.004 | 0.93 | 0.002 | 1.25E-09 | No | 3.24E-05 | 36.70 |
| 241 | 21 | rs7278859 | N/A | A | -0.01 | -0.004 | 0.69 | 0.68 | 0.004 | 0.29 | 0.002 | 4.15E-08 | No | 2.65E-05 | 29.98 |
| 242 | 2 | rs72807818 | N/A | G | -0.02 | 0.004 | 0.88 | 0.86 | 0.005 | 0.47 | 0.003 | 2.79E-14 | No | 5.10E-05 | 57.75 |
| 243 | 6 | rs72828517 | N/A | C | 0.02 | -0.007 | 0.14 | 0.17 | 0.005 | 0.13 | 0.002 | 2.83E-16 | No | 5.94E-05 | 67.18 |
| 244 | 10 | rs72840994 | N/A | G | 0.01 | 0.000 | 0.18 | 0.19 | 0.004 | 1.00 | 0.002 | 7.77E-09 | No | 2.94E-05 | 33.33 |
| 245 | 14 | rs730384 | N/A | G | -0.01 | 0.002 | 0.54 | 0.56 | 0.003 | 0.51 | 0.002 | 3.01E-09 | No | 3.12E-05 | 35.30 |
| 246 | 12 | rs7315713 | N/A | A | 0.01 | -0.001 | 0.34 | 0.31 | 0.004 | 0.78 | 0.002 | 4.34E-08 | No | 2.64E-05 | 29.87 |
| 247 | 12 | rs73301698 | N/A | A | -0.01 | 0.002 | 0.23 | 0.22 | 0.004 | 0.66 | 0.002 | 5.81E-10 | No | 3.40E-05 | 38.52 |

| | | | | | | | | | | | | | | | |
|---|---|---|---|---|---|---|---|---|---|---|---|---|---|---|---|
| 248 | 13 | rs7332724 | N/A | T | -0.01 | 0.002 | 0.26 | 0.27 | 0.004 | 0.60 | 0.002 | 1.26E-09 | No | 3.27E-05 | 36.96 |
| 249 | 10 | rs73344830 | N/A | G | -0.02 | -0.004 | 0.60 | 0.59 | 0.003 | 0.22 | 0.002 | 1.95E-23 | No | 8.83E-05 | 100.00 |
| 250 | 14 | rs736282 | N/A | C | -0.01 | -0.006 | 0.52 | 0.54 | 0.003 | 0.08 | 0.002 | 2.07E-10 | No | 3.58E-05 | 40.51 |
| 251 | 3 | rs73874335 | N/A | C | 0.020 | -0.001 | 0.94 | 0.94 | 0.007 | 0.85 | 0.004 | 3.40E-08 | No | 2.68E-05 | 30.39 |
| 252 | 21 | rs743316 | N/A | T | 0.01 | -0.007 | 0.82 | 0.79 | 0.004 | 0.12 | 0.002 | 1.20E-08 | No | 2.87E-05 | 32.46 |
| 253 | 5 | rs74643044 | N/A | T | -0.02 | 0.02 | 0.97 | 0.95 | 0.008 | 0.05 | 0.004 | 1.80E-09 | No | 3.20E-05 | 36.22 |
| 254 | 1 | rs74701752 | N/A | G | -0.02 | -0.006 | 0.90 | 0.91 | 0.006 | 0.28 | 0.003 | 2.38E-08 | No | 2.75E-05 | 31.16 |
| 255 | 17 | rs74998289 | N/A | T | 0.02 | 0.015 | 0.76 | 0.78 | 0.004 | 0.00 | 0.002 | 1.31E-17 | No | 6.46E-05 | 73.09 |
| 256 | 1 | rs7543462 | N/A | A | -0.01 | 0.000 | 0.42 | 0.46 | 0.003 | 0.96 | 0.002 | 1.25E-08 | No | 2.86E-05 | 32.38 |
| 257 | 2 | rs7594904 | N/A | C | 0.01 | -0.001 | 0.42 | 0.41 | 0.003 | 0.72 | 0.002 | 2.05E-08 | No | 2.77E-05 | 31.37 |
| 258 | 2 | rs7603132 | N/A | G | -0.01 | 0.002 | 0.85 | 0.81 | 0.004 | 0.66 | 0.002 | 9.17E-10 | No | 3.31E-05 | 37.52 |
| 259 | 2 | rs76076331 | N/A | C | -0.02 | -0.004 | 0.87 | 0.87 | 0.005 | 0.48 | 0.002 | 4.40E-14 | No | 5.04E-05 | 57.04 |
| 260 | 19 | rs76608582 | N/A | C | -0.03 | 0.007 | 0.96 | 0.94 | 0.009 | 0.40 | 0.004 | 3.11E-10 | No | 3.49E-05 | 39.53 |
| 261 | 11 | rs76878669 | N/A | C | 0.01 | -0.002 | 0.75 | 0.76 | 0.004 | 0.67 | 0.002 | 8.67E-12 | No | 4.11E-05 | 46.57 |
| 262 | 3 | rs77025239 | N/A | A | -0.01 | -0.003 | 0.11 | 0.15 | 0.005 | 0.60 | 0.002 | 1.33E-09 | No | 3.26E-05 | 36.93 |
| 263 | 11 | rs77128898 | N/A | T | -0.03 | 0.002 | 0.02 | 0.03 | 0.010 | 0.82 | 0.005 | 9.47E-09 | No | 2.92E-05 | 33.00 |
| 264 | 8 | rs77702622 | N/A | A | -0.02 | -0.007 | 0.08 | 0.06 | 0.007 | 0.32 | 0.004 | 2.99E-12 | No | 4.29E-05 | 48.60 |
| 265 | 6 | rs7773815 | N/A | A | -0.01 | 0.007 | 0.48 | 0.49 | 0.003 | 0.03 | 0.002 | 2.08E-08 | No | 2.78E-05 | 31.43 |
| 266 | 12 | rs77835879 | N/A | A | 0.02 | 0.006 | 0.91 | 0.90 | 0.006 | 0.28 | 0.003 | 2.68E-08 | No | 2.73E-05 | 30.90 |
| 267 | 7 | rs7796203 | N/A | G | 0.01 | 0.000 | 0.47 | 0.44 | 0.003 | 0.99 | 0.002 | 3.60E-10 | No | 3.48E-05 | 39.45 |
| 268 | 7 | rs7803932 | N/A | A | 0.01 | 0.004 | 0.16 | 0.16 | 0.005 | 0.43 | 0.002 | 2.44E-10 | No | 3.54E-05 | 40.04 |
| 269 | 7 | rs7808399 | N/A | A | -0.01 | -0.007 | 0.45 | 0.44 | 0.003 | 0.06 | 0.002 | 3.78E-10 | No | 3.46E-05 | 39.15 |
| 270 | 8 | rs7833201 | N/A | C | -0.02 | -0.004 | 0.13 | 0.12 | 0.005 | 0.50 | 0.003 | 5.05E-09 | No | 3.02E-05 | 34.19 |
| 271 | 9 | rs7863447 | N/A | G | -0.02 | 0.003 | 0.17 | 0.16 | 0.005 | 0.59 | 0.002 | 5.91E-13 | No | 4.58E-05 | 51.86 |
| 272 | 9 | rs7864396 | N/A | C | 0.01 | 0.002 | 0.37 | 0.36 | 0.004 | 0.50 | 0.002 | 2.95E-08 | No | 2.70E-05 | 30.59 |
| 273 | 5 | rs78721320 | N/A | G | -0.01 | -0.003 | 0.80 | 0.79 | 0.005 | 0.44 | 0.002 | 2.28E-09 | No | 3.15E-05 | 35.62 |
| 274 | 10 | rs790647 | N/A | C | 0.01 | -0.003 | 0.77 | 0.77 | 0.004 | 0.46 | 0.002 | 2.17E-13 | No | 4.76E-05 | 53.83 |
| 275 | 10 | rs7920624 | N/A | T | -0.01 | -0.004 | 0.49 | 0.48 | 0.003 | 0.29 | 0.002 | 3.97E-12 | Yes | 4.26E-05 | 48.26 |
| 276 | 10 | rs7924036 | N/A | T | 0.02 | -0.004 | 0.54 | 0.51 | 0.003 | 0.29 | 0.002 | 1.07E-18 | No | 6.89E-05 | 77.96 |
| 277 | 3 | rs79269403 | N/A | G | -0.01 | -0.002 | 0.78 | 0.78 | 0.004 | 0.68 | 0.002 | 1.17E-12 | No | 4.44E-05 | 50.31 |
| 278 | 11 | rs7928622 | N/A | T | 0.01 | -0.001 | 0.31 | 0.31 | 0.004 | 0.81 | 0.002 | 2.52E-08 | No | 2.76E-05 | 31.20 |
| 279 | 11 | rs795230 | N/A | T | 0.01 | -0.006 | 0.42 | 0.42 | 0.003 | 0.09 | 0.002 | 2.97E-08 | No | 2.71E-05 | 30.63 |

| | | | | | | | | | | | | | | | |
|---|---|---|---|---|---|---|---|---|---|---|---|---|---|---|---|
| 280 | 1 | rs79523955 | N/A | A | 0.02 | -0.001 | 0.90 | 0.90 | 0.006 | 0.92 | 0.003 | 1.87E-10 | No | 3.58E-05 | 40.54 |
| 281 | 12 | rs7977614 | N/A | G | 0.01 | -0.003 | 0.31 | 0.28 | 0.004 | 0.48 | 0.002 | 2.09E-11 | No | 3.96E-05 | 44.78 |
| 282 | 13 | rs7993663 | N/A | T | -0.01 | -0.003 | 0.64 | 0.64 | 0.004 | 0.48 | 0.002 | 3.25E-11 | No | 3.88E-05 | 43.95 |
| 283 | 14 | rs8008382 | N/A | T | -0.01 | -0.001 | 0.31 | 0.30 | 0.004 | 0.87 | 0.002 | 6.12E-11 | No | 3.77E-05 | 42.64 |
| 284 | 11 | rs80171383 | N/A | A | 0.01 | -0.009 | 0.12 | 0.14 | 0.005 | 0.07 | 0.002 | 1.83E-09 | No | 3.20E-05 | 36.20 |
| 285 | 14 | rs8020034 | N/A | G | -0.02 | 0.004 | 0.79 | 0.82 | 0.004 | 0.35 | 0.002 | 1.17E-15 | No | 5.64E-05 | 63.86 |
| 286 | 16 | rs818415 | N/A | T | -0.01 | -0.005 | 0.82 | 0.81 | 0.004 | 0.22 | 0.002 | 1.72E-08 | No | 2.81E-05 | 31.80 |
| 287 | 8 | rs837080 | N/A | C | 0.01 | 0.005 | 0.49 | 0.48 | 0.003 | 0.12 | 0.002 | 1.43E-10 | No | 3.65E-05 | 41.26 |
| 288 | 5 | rs892612 | N/A | C | 0.01 | -0.004 | 0.84 | 0.85 | 0.005 | 0.43 | 0.002 | 6.63E-10 | No | 3.37E-05 | 38.16 |
| 289 | 11 | rs894067 | N/A | G | -0.01 | 0.000 | 0.61 | 0.61 | 0.004 | 0.97 | 0.002 | 2.74E-09 | No | 3.13E-05 | 35.39 |
| 290 | 3 | rs9289300 | N/A | T | -0.02 | -0.006 | 0.82 | 0.84 | 0.005 | 0.22 | 0.002 | 1.10E-10 | No | 3.69E-05 | 41.75 |
| 291 | 6 | rs9320493 | N/A | G | -0.01 | -0.002 | 0.86 | 0.85 | 0.005 | 0.73 | 0.002 | 6.13E-09 | No | 2.98E-05 | 33.74 |
| 292 | 6 | rs9342482 | N/A | T | 0.01 | 0.004 | 0.29 | 0.25 | 0.004 | 0.27 | 0.002 | 1.36E-10 | No | 3.64E-05 | 41.17 |
| 293 | 6 | rs9349956 | N/A | A | -0.02 | -0.002 | 0.76 | 0.81 | 0.005 | 0.58 | 0.002 | 6.28E-17 | No | 6.17E-05 | 69.89 |
| 294 | 6 | rs9372625 | N/A | A | 0.02 | -0.001 | 0.41 | 0.38 | 0.004 | 0.87 | 0.002 | 6.76E-42 | No | 0.0002 | 183.33 |
| 295 | 6 | rs9384679 | N/A | T | -0.01 | 0.006 | 0.41 | 0.38 | 0.004 | 0.10 | 0.002 | 4.88E-08 | No | 2.62E-05 | 29.69 |
| 296 | 6 | rs9386319 | N/A | G | 0.01 | 0.001 | 0.43 | 0.39 | 0.004 | 0.73 | 0.002 | 1.27E-08 | No | 2.87E-05 | 32.44 |
| 297 | 1 | rs9436866 | N/A | C | 0.02 | -0.006 | 0.10 | 0.10 | 0.006 | 0.31 | 0.003 | 7.45E-11 | No | 3.75E-05 | 42.41 |
| 298 | 6 | rs9503598 | N/A | A | 0.01 | -0.007 | 0.44 | 0.44 | 0.004 | 0.05 | 0.002 | 3.12E-10 | No | 3.52E-05 | 39.82 |
| 299 | 13 | rs9529119 | N/A | C | 0.01 | -0.003 | 0.20 | 0.21 | 0.004 | 0.45 | 0.002 | 2.13E-10 | No | 3.56E-05 | 40.30 |
| 300 | 13 | rs9556958 | N/A | C | 0.01 | 0.008 | 0.47 | 0.48 | 0.003 | 0.02 | 0.002 | 2.38E-10 | No | 3.57E-05 | 40.36 |
| 301 | 22 | rs9616906 | N/A | G | -0.01 | 0.001 | 0.58 | 0.55 | 0.003 | 0.74 | 0.002 | 2.92E-18 | No | 6.69E-05 | 75.75 |
| 302 | 2 | rs9679654 | N/A | T | -0.01 | -0.002 | 0.52 | 0.57 | 0.003 | 0.55 | 0.002 | 1.29E-09 | No | 3.24E-05 | 36.70 |
| 303 | 4 | rs969512 | N/A | T | 0.01 | -0.005 | 0.30 | 0.34 | 0.004 | 0.18 | 0.002 | 3.22E-12 | No | 4.30E-05 | 48.69 |
| 304 | 11 | rs9704097 | N/A | A | -0.01 | 0.002 | 0.47 | 0.51 | 0.003 | 0.63 | 0.002 | 1.61E-09 | No | 3.21E-05 | 36.28 |
| 305 | 3 | rs9882532 | N/A | C | -0.01 | 0.002 | 0.36 | 0.35 | 0.004 | 0.55 | 0.002 | 8.17E-12 | No | 4.11E-05 | 46.58 |
| 306 | 17 | rs9914918 | N/A | G | -0.01 | -0.001 | 0.72 | 0.71 | 0.004 | 0.76 | 0.002 | 8.90E-10 | No | 3.30E-05 | 37.35 |
| 307 | 16 | rs9933256 | N/A | A | 0.01 | 0.002 | 0.59 | 0.57 | 0.004 | 0.62 | 0.002 | 4.57E-11 | No | 3.84E-05 | 43.47 |
| 308 | 16 | rs9936270 | N/A | T | -0.01 | 0.002 | 0.31 | 0.25 | 0.004 | 0.63 | 0.002 | 6.43E-12 | No | 4.17E-05 | 47.18 |
| 309 | 18 | rs9964724 | N/A | C | -0.02 | -0.004 | 0.34 | 0.32 | 0.004 | 0.24 | 0.002 | 2.66E-27 | No | 0.0001 | 116.83 |
| 310 | 4 | rs9995567 | N/A | G | -0.01 | 0.002 | 0.62 | 0.65 | 0.004 | 0.66 | 0.002 | 1.93E-08 | No | 2.78E-05 | 31.44 |

**Table 6B: Cochran's Q test**

| Method | Q | df | p value |
|---|---|---|---|
| MR Egger | 345.78 | 296 | 0.02 |
| IVW | 345.78 | 297 | 0.03 |

**Table 6C: MR-Egger intercept test**

| Egger intercept | SE | p value |
|---|---|---|
| -1.71E-06 | 0.001 | 1.00 |

**Table 6D: I^2 statistic**

| | |
|---|---|
| **I^2** | 0.98 |

**Table 6E: F statistic for the whole instrument**

| | |
|---|---|
| **F statistic** | 51.96 |

**Table 6F: MR-PRESSO global test**

| Global test | p value | Outlier index | Outlier test | Distortion test p value |
|---|---|---|---|---|
| 348.17 | 0.03 | 34 and 244 | do not alter the | 0.78 |

**Table 6G: Causal estimates**

| Method | SNPs (n) | Beta | SE | p value |
|---|---|---|---|---|
| IVW (fe) | 298 | -0.01 | 0.02 | 0.62 |
| IVW (mre) | 298 | -0.01 | 0.02 | 0.64 |
| MR Egger | 298 | -0.01 | 0.07 | 0.91 |
| Weighted median | 298 | -0.03 | 0.03 | 0.17 |

**Supplementary Table 7: Supplementary content relating to the MR analysis of the sedentary behaviour - ALS relationship.**

**Key**: SNP = single nucleotide polymorphism, PVE = proportion of variance explained, SE = standard error, EAF = effect allele frequency , ALS = amyotrophic lateral sclerosis , SSOE = strenuous sport or other exercise, VPA = vigorous physical activity, MVPA = moderate-vigorous physical activity, EA = educational attainment, BF% = body fat percentage, Chrom = chromosome, IVW = Inverse variance weighted, df = degrees of freedom, fe = fixed effects, mre = multiplicative random effects

**Conservative instrument, SNP-sedentary behaviour p<5E-08.**

**Table 7A: SNPs identified after clumping and inclusion of proxy variants**

| Index | Chrom | SNP | Proxy SNP | Effect Allele | Sedentary beta | ALS beta | Sedentary EAF | ALS EAF | ALS SE | ALS p value | Sedentary SE | Sedentary p value | Palindromic | PVE | F statistic |
|---|---|---|---|---|---|---|---|---|---|---|---|---|---|---|---|
| **1** | 3 | rs1858242 | N/A | A | 0.03 | -0.003 | 0.26 | 0.26 | 0.004 | 0.52 | 0.005 | 3.80E-09 | No | 0.0004 | 34.75 |
| **2** | 5 | rs25981 | N/A | G | 0.03 | 0.004 | 0.53 | 0.53 | 0.003 | 0.21 | 0.005 | 2.70E-09 | Yes | 0.0004 | 35.37 |
| **3** | 5 | rs26579 | N/A | G | 0.03 | -0.002 | 0.42 | 0.42 | 0.003 | 0.57 | 0.005 | 2.60E-09 | No | 0.0004 | 35.45 |
| **4** | 7 | rs34858520 | N/A | A | 0.03 | 0.002 | 0.56 | 0.56 | 0.003 | 0.47 | 0.005 | 4.50E-09 | No | 0.0004 | 34.40 |
| **5** | 1 | rs61776614 | N/A | C | 0.05 | 0.01 | 0.92 | 0.92 | 0.007 | 0.1 | 0.009 | 3.90E-08 | No | 0.0003 | 30.18 |
| **6** | 5 | rs6870096 | N/A | C | -0.03 | 0.002 | 0.32 | 0.32 | 0.004 | 0.63 | 0.005 | 2.40E-08 | No | 0.0003 | 31.15 |

**Table 7B: Cochran's Q test**

| Method | Q | df | p value |
|---|---|---|---|
| MR Egger | 1.97 | 3 | 0.58 |
| IVW | 4.07 | 4 | 0.40 |

**Table 7C: MR-Egger intercept**

| MR-Egger intercept | SE | p value |
|---|---|---|
| -0.01 | 0.01 | 0.24 |

**Table 7D: I^2 statistic**

| | |
|---|---|
| **I^2** | 0.96 |

**Table 7E: F statistic for the whole instrument**

| | |
|---|---|
| **F statistic** | 40.17 |

**Table 7F:MR-PRESSO global test**

| Global test | p value | Outlier index | Outlier test | Distortion test p value |
|---|---|---|---|---|
| 6.19 | 0.44 | N/A | N/A | N/A |

**Table 7G: Causal estimates**

| Method | SNPs (n) | Beta | SE | p value |
|---|---|---|---|---|
| IVW (fe) | 5 | 0.01 | 0.06 | 0.81 |
| IVW (mre) | 5 | 0.01 | 0.06 | 0.81 |
| MR Egger | 5 | 0.48 | 0.33 | 0.24 |
| Weighted median | 5 | -0.06 | 0.08 | 0.40 |

**Table 7H: Leave one out with fixed effects IVW**

| SNP removed | Beta | SE | p value |
|---|---|---|---|
| rs1858242 | 0.04 | 0.06 | 0.55 |
| rs26579 | 0.04 | 0.07 | 0.57 |
| rs34858520 | -0.01 | 0.06 | 0.92 |
| rs61776614 | -0.03 | 0.06 | 0.63 |
| rs6870096 | 0.03 | 0.06 | 0.62 |

**Liberal instrument, SNP- sedentary behaviour p<1E-06**

**Table 7I: SNPs identified after clumping and inclusion of proxy variants**

| Index | Chrom | SNP | Proxy SNP | Effect Allele | Sedentary beta | ALS beta | Sedentary EAF | ALS EAF | ALS SE | ALS p value | Sedentary SE | Sedentary p value | Palindromic | PVE | F statistic |
|---|---|---|---|---|---|---|---|---|---|---|---|---|---|---|---|
| **1** | 1 | rs10916119 | N/A | G | 0.03 | -0.001 | 0.82 | 0.68 | 0.004 | 0.78 | 0.007 | 3.20E-07 | No | 0.0003 | 26.13 |
| **2** | 18 | rs1187244 | N/A | T | 0.03 | -0.001 | 0.70 | 0.70 | 0.004 | 0.86 | 0.005 | 5.50E-07 | No | 0.0003 | 25.09 |
| **3** | 1 | rs17379561 | N/A | A | -0.04 | 0.005 | 0.86 | 0.86 | 0.005 | 0.31 | 0.007 | 1.20E-07 | No | 0.0003 | 27.95 |
| **4** | 2 | rs17408272 | N/A | G | 0.03 | 0.001 | 0.74 | 0.74 | 0.004 | 0.87 | 0.005 | 9.00E-07 | No | 0.0003 | 24.14 |
| **5** | 3 | rs17790917 | N/A | A | -0.03 | -0.007 | 0.79 | 0.79 | 0.004 | 0.12 | 0.006 | 1.50E-07 | No | 0.0003 | 27.57 |
| **6** | 3 | rs1858242 | N/A | A | 0.03 | -0.003 | 0.26 | 0.26 | 0.004 | 0.52 | 0.005 | 3.80E-09 | No | 0.0004 | 34.75 |
| **7** | 5 | rs25981 | N/A | G | 0.03 | 0.004 | 0.53 | 0.53 | 0.003 | 0.21 | 0.005 | 2.70E-09 | Yes | 0.0004 | 35.37 |
| **8** | 5 | rs26579 | N/A | G | 0.03 | -0.002 | 0.42 | 0.42 | 0.003 | 0.57 | 0.005 | 2.60E-09 | No | 0.0004 | 35.45 |

| | | | | | | | | | | | | | | | |
|---|---|---|---|---|---|---|---|---|---|---|---|---|---|---|---|
| **9** | 7 | rs34858520 | N/A | A | 0.03 | 0.002 | 0.56 | 0.56 | 0.003 | 0.47 | 0.005 | 4.50E-09 | No | 0.0004 | 34.40 |
| **10** | 3 | rs34910 | N/A | T | 0.03 | -0.004 | 0.72 | 0.72 | 0.004 | 0.25 | 0.005 | 3.00E-07 | No | 0.0003 | 26.27 |
| **11** | 2 | rs4953413 | N/A | T | 0.03 | -0.003 | 0.39 | 0.40 | 0.004 | 0.32 | 0.005 | 1.80E-07 | No | 0.0003 | 27.26 |
| **12** | 17 | rs57521363 | N/A | A | 0.02 | -0.006 | 0.63 | 0.62 | 0.004 | 0.08 | 0.005 | 6.90E-07 | No | 0.0003 | 24.65 |
| **13** | 1 | rs61776614 | N/A | C | 0.05 | 0.011 | 0.92 | 0.92 | 0.007 | 0.10 | 0.009 | 3.90E-08 | No | 0.0003 | 30.18 |
| **14** | 5 | rs6870096 | N/A | C | -0.03 | 0.002 | 0.32 | 0.32 | 0.004 | 0.63 | 0.005 | 2.40E-08 | No | 0.0003 | 31.15 |
| **15** | 21 | rs714167 | N/A | T | 0.07 | 0.008 | 0.97 | 0.97 | 0.009 | 0.41 | 0.01 | 4.70E-07 | No | 0.0003 | 25.36 |
| **16** | 4 | rs731069 | N/A | A | -0.02 | 0.003 | 0.54 | 0.53 | 0.003 | 0.45 | 0.005 | 2.20E-07 | No | 0.0003 | 26.87 |
| **17** | 16 | rs75779711 | N/A | C | -0.04 | 0.007 | 0.85 | 0.82 | 0.005 | 0.16 | 0.007 | 2.50E-07 | No | 0.0003 | 26.62 |

**Table 7J: Cochran's Q test**

| Method | Q | df | p value |
|---|---|---|---|
| MR Egger | 11.48 | 14 | 0.65 |
| IVW | 15.06 | 15 | 0.45 |

**Table 7K: MR-Egger intercept test**

| Egger intercept | SE | p value |
|---|---|---|
| -0.009 | 0.005 | 0.08 |

**Table 7L: I^2 statistic**

| | |
|---|---|
| **I^2** | 0.96 |

**Table 7M: F statistic for the whole instrument**

| | |
|---|---|
| **F statistic** | 30.9 |

**Table 7N: MR-PRESSO global test**

| Global test | p value | Outlier index | Outlier test | Distortion test p value |
|---|---|---|---|---|
| 16.99 | 0.49 | N/A | N/A | N/A |

**Table 7O: Causal estimates**

| Method | SNPs (n) | Beta | SE | p value |
|---|---|---|---|---|

| | | | | |
|---|---|---|---|---|
| IVW (fe) | 16 | -0.04 | 0.03 | 0.29 |
| IVW (mre) | 16 | -0.04 | 0.03 | 0.29 |
| MR Egger | 16 | 0.27 | 0.17 | 0.12 |
| Weighted median | 16 | -0.07 | 0.05 | 0.17 |

**Table 7P: Leave one out with fixed effects IVW**

| SNP removed | Beta | SE | p value |
|---|---|---|---|
| rs10916119 | -0.03655663 | 0.03560383 | 0.3045328 |
| rs1187244 | -0.03657028 | 0.03481913 | 0.2935835 |
| rs17379561 | -0.02940193 | 0.03490209 | 0.3995575 |
| rs17408272 | -0.03927275 | 0.03478176 | 0.2588477 |
| rs17790917 | -0.05146789 | 0.03489181 | 0.1401935 |
| rs1858242 | -0.03246569 | 0.03516911 | 0.355939 |
| rs26579 | -0.03315084 | 0.0352251 | 0.3466462 |
| rs34858520 | -0.04585502 | 0.03516657 | 0.1922548 |
| rs34910 | -0.02839437 | 0.03486538 | 0.4154158 |
| rs4953413 | -0.02959515 | 0.03489904 | 0.3964258 |
| rs57521363 | -0.02354414 | 0.03480632 | 0.4987658 |
| rs61776614 | -0.05253169 | 0.03492052 | 0.1324986 |
| rs6870096 | -0.03407343 | 0.03502104 | 0.3305823 |
| rs714167 | -0.04485179 | 0.03482012 | 0.1977112 |
| rs731069 | -0.03160979 | 0.03487316 | 0.3647127 |
| rs75779711 | -0.02607637 | 0.03489485 | 0.454892 |

**Supplementary Table 8: This table shows all pathways associated with acute exercise at two minutes post activity. The proportion of genes in each of these pathways which are statistically significant for missense, disease and damaging mutations in ALS patients (MAF < 1%) is presented. The significance of this proportion is then expressed both as an uncorrected p value and a corrected false discovery rate (FDR). Pathways with a significance under 0.0006 after comparing against 5000 iterations were then compared against 30,000 iterations in order to ensure accuracy.**

| Pathway | Genes (n) | Enriched ALS risk (%) | P value (5000 permutations) | P value (30000 permutations) | FDR |
|---|---|---|---|---|---|
| B Cell Receptor Signaling | 90 | 19 | 2.00E-04 | 5.30E-04 | 0.02 |
| iCOS-iCOSL Signaling in T Helper Cells | 61 | 18 | 5.20E-03 | N/A | 0.03 |
| IL-4 Signaling | 50 | 22 | 0.001 | N/A | 0.02 |
| T Cell Receptor Signaling | 57 | 21 | 2.00E-04 | 6.00E-04 | 0.02 |
| Sphingosine-1-phosphate Signaling | 60 | 20 | 0.0018 | N/A | 0.02 |
| CD28 Signaling in T Helper Cells | 62 | 21 | 8.00E-04 | N/A | 0.02 |
| Leptin Signaling in Obesity | 45 | 22 | 0.002 | N/A | 0.02 |
| P2Y Purigenic Receptor Signaling Pathway | 63 | 19 | 0.002 | N/A | 0.02 |
| Virus Entry via Endocytic Pathways | 55 | 20 | 0.002 | N/A | 0.03 |
| NF-KB Activation by Viruses | 47 | 19 | 0.006 | N/A | 0.04 |
| PDGF Signaling | 48 | 21 | 0.001 | N/A | 0.02 |
| Reelin Signaling in Neurons | 46 | 22 | 1.80E-03 | N/A | 0.02 |
| CTLA4 Signaling in Cytotoxic T Lymphocytes | 47 | 19 | 0.005 | N/A | 0.03 |
| Role of Macrophages, Fibroblasts and Endothelial Cells in Rheumatoid Arthritis | 114 | 15 | 0.006 | N/A | 0.04 |
| Cardiac Hypertrophy Signaling | 90 | 16 | 0.01 | N/A | 0.05 |
| Production of Nitric Oxide and Reactive Oxygen Species in Macrophages | 76 | 16 | 0.009 | N/A | 0.05 |
| Macropinocytosis Signaling | 42 | 19 | 6.20E-03 | N/A | 0.04 |
| Renin-Angiotensin Signaling | 55 | 18 | 0.007 | N/A | 0.04 |
| Endocannabinoid Cancer Inhibition Pathway | 64 | 19 | 3.00E-03 | N/A | 0.03 |
| HGF Signaling | 52 | 17 | 0.01 | N/A | 0.05 |
| Dendritic Cell Maturation | 76 | 18 | 2.00E-03 | N/A | 0.02 |
| Endocannabinoid Developing Neuron Pathway | 54 | 19 | 0.005 | N/A | 0.03 |
| PTEN Signaling | 52 | 17 | 0.01 | N/A | 0.05 |
| Melanocyte Development and Pigmentation Signaling | 45 | 20 | 0.004 | N/A | 0.03 |
| Neuroinflammation Signaling Pathway | 106 | 17 | 0.001 | N/A | 0.02 |

| | | | | | |
|---|---|---|---|---|---|
| Glioblastoma Multiforme Signaling | 65 | 18 | 0.001 | N/A | 0.02 |
| PKCθ Signaling in T Lymphocytes | 64 | 17 | 0.008 | N/A | 0.04 |
| IL-3 Signaling | 40 | 20 | 0.005 | N/A | 0.03 |
| Rac Signaling | 50 | 18 | 0.010 | N/A | 0.05 |
| IL-15 Signaling | 37 | 19 | 0.01 | N/A | 0.05 |
| IL-2 Signaling | 33 | 21 | 0.007 | N/A | 0.04 |
| Gα12/13 Signaling | 55 | 18 | 0.004 | N/A | 0.03 |
| Angiopoietin Signaling | 37 | 24 | 0.001 | N/A | 0.02 |
| Pancreatic Adenocarcinoma Signaling | 48 | 21 | 0.001 | N/A | 0.02 |
| RANK Signaling in Osteoclasts | 42 | 24 | 4.00E-04 | 4.70E-04 | 0.02 |
| EGF Signaling | 31 | 23 | 0.004 | N/A | 0.03 |
| Prostate Cancer Signaling | 42 | 19 | 0.006 | N/A | 0.04 |
| Mouse Embryonic Stem Cell Pluripotency | 44 | 18 | 0.01 | N/A | 0.05 |
| IL-17A Signaling in Airway Cells | 33 | 24 | 0.001 | N/A | 0.02 |
| Ephrin A Signaling | 27 | 26 | 0.001 | N/A | 0.02 |
| CD40 Signaling | 33 | 24 | 8.00E-04 | N/A | 0.02 |
| GM-CSF Signaling | 33 | 21 | 0.007 | N/A | 0.04 |
| LPS-stimulated MAPK Signaling | 37 | 19 | 0.01 | N/A | 0.05 |
| PEDF Signaling | 37 | 22 | 0.0014 | N/A | 0.02 |
| NGF Signaling | 46 | 24 | 2.00E-04 | 2.70E-04 | 0.02 |
| p53 Signaling | 42 | 21 | 0.0024 | N/A | 0.03 |
| IL-17 Signaling | 36 | 19 | 0.009 | N/A | 0.05 |
| Role of IL-17A in Arthritis | 29 | 21 | 0.01 | N/A | 0.05 |
| FLT3 Signaling in Hematopoietic Progenitor Cells | 36 | 22 | 0.003 | N/A | 0.03 |
| IL-9 Signaling | 21 | 29 | 0.001 | N/A | 0.02 |
| Melanoma Signaling | 26 | 23 | 0.004 | N/A | 0.03 |
| Docosahexaenoic Acid (DHA) Signaling | 23 | 30 | 2.00E-04 | 4.00E-04 | 0.02 |
| Small Cell Lung Cancer Signaling | 33 | 21 | 0.003 | N/A | 0.03 |
| HIF1α Signaling | 44 | 20 | 0.003 | N/A | 0.03 |
| Neurotrophin/TRK Signaling | 32 | 22 | 0.003 | N/A | 0.03 |
| Aldosterone Signaling in Epithelial Cells | 56 | 18 | 0.007 | N/A | 0.04 |
| Chronic Myeloid Leukemia Signaling | 40 | 20 | 0.004 | N/A | 0.03 |

| | | | | | |
|---|---|---|---|---|---|
| Renal Cell Carcinoma Signaling | 33 | 21 | 0.005 | N/A | 0.03 |
| Ovarian Cancer Signaling | 50 | 18 | 0.007 | N/A | 0.04 |
| SPINK1 General Cancer Pathway | 30 | 23 | 0.003 | N/A | 0.03 |
| Lymphotoxin β Receptor Signaling | 26 | 27 | 0.001 | N/A | 0.02 |
| Role of PI3K/AKT Signaling in the Pathogenesis of Influenza | 29 | 21 | 0.008 | N/A | 0.04 |
| Role of p14/p19ARF in Tumor Suppression | 18 | 28 | 0.003 | N/A | 0.03 |
| TR/RXR Activation | 35 | 23 | 0.002 | N/A | 0.03 |
| FGF Signaling | 32 | 31 | 2.00E-04 | 1.00E-04 | 0.02 |
| Human Embryonic Stem Cell Pluripotency | 45 | 20 | 0.003 | N/A | 0.03 |
| Cell Cycle Control of Chromosomal Replication | 21 | 24 | 0.004 | N/A | 0.03 |
| Wnt/Ca+ pathway | 23 | 26 | 0.002 | N/A | 0.02 |
| Amyotrophic Lateral Sclerosis Signaling | 36 | 25 | 4.00E-04 | 7.00E-04 | 0.02 |
| Estrogen-Dependent Breast Cancer Signaling | 28 | 29 | 6.00E-04 | 1.60E-03 | 0.02 |
| Regulation of the Epithelial-Mesenchymal Transition Pathway | 55 | 20 | 0.002 | N/A | 0.02 |
| Formaldehyde Oxidation II (Glutathione-dependent) | 2 | 50 | 0.008 | N/A | 0.04 |

**Supplementary Table 9: ALS-related genes which are differentially expressed following exercise.** The data presented includes the false discovery rate (FDR) and fold change (FC) with respect to the response to acute exercise measured at four time points post exercise.

| Gene name | All TP (FDR) | 2 min (FDR) | 15 min (FDR) | 30 min (FDR) | 1 hour (FDR) | 2 min (log2FC) | 15 min (log2FC) | 30 min (log2FC) | 1 hour (log2FC) |
|---|---|---|---|---|---|---|---|---|---|
| **ARHGEF28** | 5.40E-28 | 1.50E-12 | 5.00E-11 | 5.40E-03 | 1.00E+00 | 0.5 | 0.4 | 0.3 | 0 |
| **OPTN** | 5.70E-15 | 1.50E-09 | 2.00E-04 | 9.90E-03 | 1.00E+00 | 0.2 | 0.1 | 0.1 | 0 |
| **PFN1** | 9.90E-14 | 9.50E-10 | 7.20E-01 | 1.00E+00 | 1.00E+00 | 0.3 | 0 | 0 | -0.1 |
| **TBK1** | 1.60E-13 | 2.10E-12 | 1.40E-04 | 1.60E-02 | 1.00E+00 | 0.3 | 0.2 | 0.2 | 0.1 |
| **VCP** | 9.60E-13 | 8.10E-10 | 6.80E-06 | 4.60E-02 | 1.00E+00 | 0.2 | 0.1 | 0.1 | 0 |
| **TUBA4A** | 3.70E-12 | 7.40E-04 | 6.30E-01 | 2.00E-01 | 1.50E-01 | -0.2 | -0.1 | 0.2 | 0.2 |
| **CYP27A1** | 1.00E-11 | 8.00E-04 | 2.40E-01 | 1.00E+00 | 2.10E-02 | -0.2 | -0.2 | 0 | 0.3 |
| **VAPB** | 1.80E-11 | 1.20E-05 | 1.10E-01 | 1.00E+00 | 3.60E-01 | -0.1 | -0.1 | 0 | 0.1 |
| **GBA2** | 2.40E-10 | 1.90E-07 | 1.20E-02 | 5.50E-01 | 1.00E+00 | 0.2 | 0.1 | 0.1 | 0 |
| **EWSR1** | 8.90E-08 | 5.00E-02 | 4.70E-01 | 1.00E+00 | 1.50E-01 | 0.1 | 0.1 | -0.1 | -0.1 |
| **GRN** | 1.30E-06 | 5.60E-05 | 5.50E-03 | 5.20E-01 | 1.00E+00 | -0.5 | -0.2 | -0.2 | 0 |
| **SETX** | 3.00E-06 | 8.00E-07 | 3.00E-01 | 5.10E-01 | 1.00E+00 | 0.2 | 0.1 | 0.1 | 0 |
| **ALS2** | 4.50E-05 | 5.30E-03 | 2.40E-02 | 1.00E+00 | 1.00E+00 | 0.1 | 0.1 | 0 | 0 |
| **SPG20** | 8.30E-05 | 9.80E-01 | 6.80E-01 | 1.00E+00 | 1.70E-01 | 0 | -0.1 | 0.1 | 0.1 |
| **HNRNPA1** | 1.70E-04 | 2.10E-02 | 7.90E-02 | 1.30E-02 | 4.00E-03 | -0.1 | -0.1 | -0.1 | -0.1 |
| **C9orf72** | 2.10E-04 | 9.20E-04 | 5.80E-01 | 1.00E+00 | 1.00E+00 | -0.2 | 0 | 0 | 0 |
| **UBQLN2** | 2.20E-04 | 2.20E-02 | 2.70E-04 | 7.00E-02 | 1.90E-01 | -0.1 | -0.1 | -0.1 | -0.1 |
| **SPG11** | 1.30E-03 | 4.40E-03 | 9.30E-03 | 1.20E-01 | 1.10E-01 | 0.1 | 0.1 | 0.1 | 0.1 |
| **ATXN2** | 1.70E-03 | 1.10E-01 | 1.00E+00 | 1.00E+00 | 9.60E-01 | -0.1 | 0 | 0 | 0.1 |
| **DCTN1** | 3.10E-03 | 2.60E-02 | 1.00E+00 | 1.00E+00 | 1.00E+00 | 0.1 | 0 | 0 | 0 |
| **CHCHD10** | 4.40E-03 | 1.90E-02 | 9.90E-02 | 1.00E+00 | 1.00E+00 | -0.2 | -0.1 | 0 | 0 |
| **ANG** | 1.30E-02 | 4.40E-01 | 4.80E-01 | 1.00E+00 | 1.00E+00 | 0 | -0.1 | 0 | 0 |
| **FUS** | 3.00E-02 | 1.00E+00 | 6.20E-01 | 1.00E+00 | 1.00E+00 | -0.1 | 0.1 | -0.1 | -0.1 |

**Table 9B: The full clinically validated ALS-related gene panel assessed for differential expression during acute exercise**

| Gene name |
|---|
| ALS2 |

| |
|---|
| ANG |
| ANXA11 |
| ARHGEF28 |
| ATXN2 |
| C9orf72 |
| CHCHD10 |
| CHMP2B |
| CYP27A1 |
| DAO |
| DCTN1 |
| ERBB4 |
| EWSR1 |
| FIG4 |
| FUS |
| GBA2 |
| GRN |
| HNRNPA1 |
| HNRNPA2B1 |
| KIF5A |
| MAPT |
| MATR3 |
| NEFH |
| NEK1 |
| OPTN |
| PFN1 |
| PRPH |
| SETX |
| SIGMAR1 |
| SOD1 |
| SPAST |
| SPG20 |
| SPG11 |

| |
|---|
| SQSTM1 |
| SS18L1 |
| TAF15 |
| TARDBP |
| TBK1 |
| TUBA4A |
| UBQLN2 |
| VAPB |
| VCP |
| VPS54 |
| VRK1 |

### METHODOLOGICAL LIMITATIONS

#### Two-sample mendelian randomisation

The effect of SSOE upon ALS has been modelled through the dichotomisation of a continuous variable, no particular significance can be appointed to the point of dichotomisation and a causal effect parameter cannot be meaningfully assigned.[1] We are therefore able to provide a positive direction of effect, but unable to assign an odds ratio to the exposure. Although the ten SNP conservative instruments for SSOE is positively correlated with ALS, it does not reach significance. This appears to be related to power and does not nullify our findings, but does increase the risk of pleiotropy having impacted upon our results, which we have minimised through a number of robust measures. Because individual SNPs explain a small fraction of complex traits such as those addressed in this study, a further potential source of MR error is insufficient power which may increase the risk of type 1 error in our low-intensity PA results. In order to address power limitations, we implemented a liberal exposure instrument in our MR. The risk of type 1 error was reduced by calculation with a range of robust measures and assessments of instrument strength. Self-report questionnaire and interview data in this study are vulnerable to well recognised limitations including recall bias and social desirability bias.

#### Case-control study of historical PA

The HAPAQ is designed to optimise accurate data collection.[2] All questions are closed so as to minimize ambiguity.[3] The separation of PA into domains matches memory encoding of PA.[4] To aid recall of duration for each activity the data were collected in a disaggregated manner: participants were asked first about activity type, and then frequency and finally duration. A life calendar was constructed and repeatedly referred to during the interview to facilitate recall using temporal memory cues.[5] The HAPAQ was validated using a mixed gender population of comparable average age to the participants in the present study;[2] but these individuals did not suffer neurological disease which may alter recall of PA.

### R CODE UTILISED FOR MR AND BURDEN TESTING:

**# MENDELIAN RANDOMISATION CODE UTILISED.**
**#This code was modified for each calculation by inputting the relevant exposure GWAS. The basic format is presented here.**

**# 1. Load required packages and load in exposure data locally**

```
library(readr)
library(TwoSampleMR)
library(dplyr)

exposuregwas  <-  read_table2([GWAS EXPOSURE DATA LOCATION HERE,READR
FUNCTION MAY DIFFER ACCORDING TO THE GWAS FORMATTING])

renamedexposuregwas <- rename(exposuregwas , c(  [INSERT COLUMN NAME CHANGES IF
REQUIRED TO COMPLY WITH TwoSampleMR FORMAT]  ) )

dfEx <- as.data.frame(renamedexposuregwas)

sig_threshold<- dfEx %>% filter(pval < [INSERT NUMERICAL  SIGNIFICANCE LEVEL HERE] )

exposure <- format_data(sig_threshold , type = "exposure" )

exposure$exposure  <- gsub( "exposure" , “[INSERT EXPOSURE PHENOTYPE NAME HERE]”
, exposure$exposure )
```

**# 2. Clump exposure data**

```
clumpedexposure <- clump_data(exposure)
```

**# 3. If you require a proxy SNP, insert now with this code (skip step if no proxy)**

```
proxysnp <- dfEx %>%
  filter (SNP %in%  c(“[LIST OF PROXY SNPS GO HERE]”))

proxyexp <- format_data(proxysnp , "exposure")

proxyexp$exposure  <- gsub( "exposure" , “[INSERT EXPOSURE PHENOTYPE NAME HERE]”
, proxyexp$exposure )

clumpedexposure <- rbind(clumpedexposure, proxyexp)
```

**# 4. Load outcome data locally**

```
outcome_dat <- read_outcome_data(
  snps = clumpedexposure$SNP,
  [INSERT DETAILS OF OUTCOME DATA LOCATION AND COLUMN NAMES HERE AS PER TwoSampleMR FORMAT]
)

outcome_dat$outcome  <- gsub( "outcome" , “[INSERT OUTCOME PHENOTYPE NAME HERE]” , outcome_dat$outcome )
```

**# 5. If you are uncertain of requirement for proxy SNPs , check if any SNPs are not present in both data sets (skip step if you’re certain no proxy is required)**

```
missing <- clumpedexposure %>%
  filter ( !SNP %in% outcome_dat$SNP)

View(missing)
```

**# 6. If an SNP was identified to be missing in step 5, you need to locate an appropriate proxy. If no proxy is required skip this step. Note the first column in the data frame must be the SNP column.**

```
library(LDlinkR)
for (i in 1:nrow(missing))  {
  x <- LDproxy(missing [i , 1], pop = "EUR", r2d = "r2", token = “[INSERT TOKEN ID HERE]”)

  eligible <- x[x$R2 >= 0.9 , ]

  A <- dfEx %>%
    filter ( SNP %in% eligible$RS_Number)

  Common <- [DATA FRAME CONTAINING THE ENTIRE OUTCOME GWAS HERE] %>%
    filter ( SNP %in% A$SNP)

  R2vals <- eligible %>%
    filter( RS_Number %in% Common$SNP)
  missing$proxySNP[i] <- R2vals[1 , 1]
}

View(missing) #missing now contains a column of proxy SNPs to use at step 3
```

**# 7. If you have used a proxy SNP, there will be two different 'id.exposure' names, they must be changed to the same value for harmonisation (if not proxy was inserted then skip this step)**

```
clumpedexposure$id.exposure  <- gsub( "[ID 1]" , "[ID 2]" , clumpedexposure$id.exposure )
```

**# 8 Harmonise the data, the default setting is that palindromic alleles with intermediate effect allele frequency are removed**

```
dat <- harmonise_data(
  exposure_dat = clumpedexposure,
  outcome_dat = outcome_dat ,
)
```

**# 9. Add a column of PVE and F statistics. During the above processes only EAF have been retained and therefore MAF needs to be calculated during this process.**

**# A) Calculate a per-SNP F statistic**

```
dat$EAF2 <- (1 - dat$eaf.exposure)
dat$MAF <- pmin(dat$eaf.exposure, dat$EAF2)
PVEfx <- function(BETA, MAF, SE, N){
  pve <- (2*(BETA^2)*MAF*(1 - MAF))/
    ((2*(BETA^2)*MAF*(1 - MAF)) + ((SE^2)*2*N*MAF*(1 - MAF)))
  return(pve)
}
dat$PVE <- mapply(PVEfx,
                  dat$beta.exposure,
                  dat$MAF,
                  dat$se.exposure,
                  N = [INSERT EXPOSURE GWAS POPULATION SIZE)
dat$FSTAT <- (([INSERT EXPOSURE GWAS POPULATION SIZE] - 1 - 1)/1)*(dat$PVE/(1 - dat$PVE))
```

**# B) Calculate a total instrument F statistic**

```
(([INSERT EXPOSURE GWAS POPULATION SIZE] - [INSERT TOTAL NUMBER OF SNPS IN INSTRUMENT] - 1)/[INSERT TOTAL NUMBER OF SNPS IN INSTRUMENT])*([INSERT TOTAL PVE]/(1 - [INSERT TOTAL PVE]))
```

**#10. Calculate the cochran's Q, egger intercept, causal estimates, I^2, MR-PRESSO , leave one out and develop graphs**

**# Cochran's Q**

```
View(mr_heterogeneity(dat))
```

**#Egger intercept**

```
View(mr_pleiotropy_test(dat))
```

**#I^2**

```
Isq( dat$beta.exposure , dat$se.exposure)
```

**#MR-PRESSO**

```
run_mr_presso(dat)
```

**#MR causal estimates and odds ratios (OR not used in this paper due to binary exposure)**

```
View(generate_odds_ratios(  mr(dat , method_list =  c("mr_ivw_fe" , "mr_ivw_mre" ,
"mr_egger_regression" , "mr_weighted_median"))))
```

**#Leave one out analysis**

```
View(mr_leaveoneout(dat, method = [INSERT MOST APPROPRIATE MR METHOD AS PER
TwoSampleMR FORMAT]))
```

**#Scatter plot**

```
mr_scatter_plot(mr(dat, method_list = c( "mr_ivw_fe" , "mr_ivw_mre" , "mr_egger_regression" ,
"mr_weighted_median")), dat)
```

**#Funnel plot**

```
mr_funnel_plot( mr_singlesnp(
  dat,
  parameters = default_parameters(),
  single_method = "mr_wald_ratio",
  all_method = c("mr_ivw_fe" , "mr_ivw_mre" , "mr_egger_regression" , "mr_weighted_median")
))
```

**#BURDEN TESTING CODE UTILISED FOR EXERCISE-RELATED GENES IN ALS PATIENTS.**
**#This has not been made generic, as only one set of data was analysed with this code (i.e. the exercise transcript and ALS transcript data). The data utilised is available in supplementary table 2.5 in the study of origin. We only imported pathways associated at 2 minutes post-exercise into R.**

**# Pre analysis, load in relevant packages**

**library(dplyr)**

**# 1. Load in the transcripts which are differentially expressed in response to exercise from Contrepois et al. 2020.**
**# split each pathway into its constituent genes.**
**# name according to the pathway.**

transcript_upregulated_exercise <- read_csv("transcript_upregulated_exercise.csv")

two_mins <- transcript_upregulated_exercise [ , c("Pathway" , "sig_genes_2 min" ) ]

split <- str_split_fixed(two_mins$`sig_genes_2 min` , "[,/]" , 180 )

row.names (split) <- two_mins$`Pathway`

**# 2. Load in the Project MinE transcript data (MAF1%)**

ProjectMinE_Transcripts_0_01_results <-
read_table2("burden/ProjectMinE.Transcripts.0.01.results.txt")

**# 3. identify which genes enriched in exercise are significantly related to MND.**

project_mine_enriched <- subset(ProjectMinE_Transcripts_0_01_results , GeneName %in% split ,)

```
p_missense <- project_mine_enriched %>%
  group_by(GeneName) %>%
  slice(which.min(p.firth.dis.dam.miss))
```

sigmiss <- p_missense[p_missense$p.firth.dis.dam.miss <0.05, ]

**# 4. produce a vector of significant gene names**

genes_sig <- sigmiss$GeneName[!duplicated(sigmiss$GeneName)]

**# 5. Create a loop which calculates the proportion of exercise-related genes in each pathway which are significant in MND.**
**# Note, 1 is subtracted from the length as a blank box is counted at the end of each row.**
**result <- rep(0,length (split[,1]))**

names(result) <- row.names (split)

```
for (i in 1:nrow(split))
{
  x1 <- unique (split[i,])
  x2 <- which (genes_sig %in% x1)
  result[i] <- (length (x2) / (length (x1)-1))
}
```

**# 6. Create a loop which calculates the total number of genes in each pathway. 1 is subtracted from the total genes to remove empty cells as a count.**

```
Total_Genes <- rep(0 , nrow(split))

for (i in 1:nrow(split))
{
  x1 <- unique (split[i,])

  Total_Genes[i] <- (length (x1)-1)
}
```

**# 7. Create a data frame which contains the pathway and gives information about the total number of significant genes.**

```
result.df <- data.frame( result , two_mins , Total_Genes)

View(result.df)
```

**# 8. Compare the results to 5000 random iterations**

```
genes_only <- ProjectMinE_Transcripts_0_01_results %>%  distinct(GeneName)

gene_sig <- sigmiss %>% distinct(GeneName)

results <-matrix (nrow = 323 , ncol=5000 )

for ( x in 1:5000) {
  for (i in 1:323)
  {
    a <- data.frame(i , result.df[ i , 2] , c(sample_n(genes_only , result.df[i , 4], replace = FALSE)))
    b <- which(a$GeneName %in% gene_sig$GeneName)
    c <- length(b) / result.df [i,4]
    d <- c >  result.df[i , 1]
    results [i , x ] <- d
  }
}
```

```
pvals <- matrix(nrow = 323 , ncol =1)
for ( m in 1:323)
{
  A <- sum(results[ m , ])
  B <- A / 5000
  pvals [m , 1] <- B
}

table <- cbind(result.df[ 1:nrow(results), 2] , result.df[1:nrow(results), 4], pvals)
```

**# 9. Calculate the false discovery rate . Note, prior to this stage we calculated p values for very significant pathways by operating 30,000 random iterations as per the methods section. Therefore, the table was edited with new p values for very significant pathways before this stage.**

```
FDR <- p.adjust(table[ , 3] , method = "BH", n = length(table))

fulltable <- cbind(table , FDR )

write.csv(fulltable,file = file.choose(new = T))
```

### REFERENCES


1 Burgess S, Labrecque JA. Mendelian randomization with a binary exposure variable: interpretation and presentation of causal estimates. European Journal of Epidemiology. 2018; **33**: 947–52.

2 Besson H, Harwood CA, Ekelund U, *et al.* Validation of the historical adulthood physical activity questionnaire (HAPAQ) against objective measurements of physical activity. *Int J Behav Nutr Phys Act* 2010; **7**: 54.

3 Durante R, Ainsworth BE. The recall of physical activity: using a cognitive model of the question-answering process. *Med Sci Sports Exerc* 1996; **28**: 1282–91.

4 Baranowski T, Domel SB. A cognitive model of children's reporting of food intake. *Am J Clin Nutr* 1994; **59**: 212S – 217S.

5 Friedenreich CM. IMPROVING LONGTERM RECALL IN EPIDEMIOLOGIC STUDIES. Epidemiology. 1994; **5**: 1–3.
